# Fetal Gene Regulatory Gene Deletions are Associated with Poor Cognition in Schizophrenia and Community-Based Samples

**DOI:** 10.1101/2024.08.02.24311302

**Authors:** Jennifer K. Forsyth, Jinhan Zhu, Ariana S. Chavannes, Zachary H. Trevorrow, Mahnoor Hyat, Sam A. Sievertsen, Sophie Ferreira-Ianone, Matthew P. Conomos, Keith H. Nuechterlein, Robert F. Asarnow, Michael F. Green, Katherine H. Karlsgodt, Diana O. Perkins, Tyrone D. Cannon, Jean M. Addington, Kristen S. Cadenhead, Barbara A. Cornblatt, Matcheri S. Keshavan, Daniel H. Mathalon, William S. Stone, Ming T. Tsuang, Elaine F. Walker, Scott W. Woods, Katherine L. Narr, Sarah C. McEwen, Charles H. Schleifer, Cindy M. Yee, Caroline K. Diehl, Anika Guha, Gregory A. Miller, Aaron F. Alexander-Bloch, Sha Zhiqiang, Glessner Joseph, Jakob Seidlitz, Richard A. I. Bethlehem, Roel A. Ophoff, Carrie E. Bearden

**Affiliations:** Department of Psychology, University of Washington, WA, USA; Institute for Public Health Genetics, University of Washington, WA, USA; Department of Biostatistics, University of Washington, WA, USA; Department of Psychiatry and Biobehavioral Sciences, University of California, Los Angeles, CA, USA; VA Greater Los Angeles Healthcare System, Los Angeles, CA, USA; Department of Psychology, University of California, Los Angeles, CA, USA; Department of Psychiatry, University of North Carolina, Chapel Hill, NC, USA; Department of Psychology, Yale University, New Haven, CT, USA; Department of Psychiatry, University of Calgary, Calgary, Alberta, Canada; Department of Psychiatry, University of California, San Diego, CA, USA; Department of Psychiatry, Donald and Barbara Zucker School of Medicine at Hofstra/Northwell, Hempstead, NY, USA; Department of Psychiatry, Harvard Medical School at Beth Israel Deaconess Medical Center, Boston, Massachusetts, USA; Department of Psychiatry, University of California, and San Francisco Veterans Affairs Medical Center, San Francisco, CA, USA; Department of Psychology, Emory University, Atlanta, GA, USA; Department of Psychiatry, Yale University, New Haven, CT, USA; Department of Neurology, University of California, Los Angeles, CA, USA; Department of Psychiatry, University of Pennsylvania, Philadelphia, PA, USA; Department of Psychology, University of Cambridge, Cambridge, UK; Department of Human Genetics, University of California, Los Angeles, CA, USA

## Abstract

**Objective:** Schizophrenia is a neurodevelopmental disorder involving clinical and genetic heterogeneity. Multiple recurrent copy number variants (CNVs) increase risk for schizophrenia spectrum disorders (SSD). However, how known risk CNVs and broader genome-wide CNVs influence clinical variability is unclear. Furthermore, whether biological annotation of CNV scores can improve power for patient stratification is unknown.

**Methods:** This study examined associations between severe phenotypes in 617 SSD individuals, namely, child-onset psychosis or borderline intellectual functioning (IQ), and: 1) known risk CNVs; 2) genome-wide deletion burden scores; and 3) novel scores capturing deletion burden in 18 previously validated and mutually exclusive gene-sets, representing distinct aspects of neurodevelopment. Associations with borderline IQ were assessed for replicability in 233 SSD-relatives and 581 controls, and 9,930 youth from the Adolescent Brain Cognitive Development (ABCD) Study.

**Results:** Known SSD- (odds ratios (OR)=7.07, 95%CI[1.60,31.32]) and neurodevelopmental disorder (NDD)-risk CNVs (OR=4.56, 95%CI[1.48,14.10]) were associated with borderline IQ in SSD. Furthermore, beyond effects of known NDD-risk CNVs, deletion of genes involved in regulating gene expression during fetal brain development was associated with borderline IQ across SSD cases and non-cases (OR=2.57, 95%CI[1.44,4.60]), and in the ABCD cohort (OR=1.33, 95%CI[1.00,1.76]). Exploratory structural MRI-based analyses showed associations between fetal gene regulatory gene deletions and altered gray matter volume (*b*=0.09, 95%CI[0.004,0.17]) and cortical thickness (*b*=0.14, 95%CI[0.05,0.24]) across SSD cases and non-cases.

**Conclusions:** Results confirm contributions of known risk CNVs to severe phenotypes in SSD, implicate disrupted fetal brain development in poor cognition, and demonstrate the utility of a neurodevelopmental framework for identifying mechanisms underlying severe SSD-relevant phenotypes.

## INTRODUCTION

One of the greatest barriers to understanding the core biological processes that underlie schizophrenia spectrum disorders (SSD; i.e., schizophrenia and other psychotic disorders) and improving patient outcomes is the clinical and genetic heterogeneity of these disorders. For example, while psychosis onset is most common in late adolescence or early adulthood, a subset of patients experience psychosis as early as childhood(1). Similarly, while cognitive functioning in SSD is, on average, 1 to 2 standard deviations below healthy individuals(2), some patients function in the above average range, while others function in the borderline or intellectual disability (ID) range. Importantly, early psychosis onset and poor cognitive function are associated with poor long-term outcomes, including a more chronic course of illness(3–5). Identifying etiologic factors underlying severe clinical phenotypes is crucial for personalized medicine approaches to care.

One class of genetic variants that may contribute to severe phenotypes in SSD are rare copy number variants (CNVs), in which large stretches of DNA are deleted or duplicated. CNVs occur across the genome; however, regions flanked by low-copy repeats (LCRs) are particularly vulnerable to non-allelic homologous recombination and therefore recurrent CNVs(6). The largest study of CNVs in SSD by the Psychiatric Genomics Consortium (PGC) identified 13 recurrent loci as SSD-associated(7).

Although only a small portion of cases carry these CNVs, most increase risk substantially when present(7). Notably, the majority of SSD-associated CNVs also increase risk for early-onset neurodevelopmental disorders (NDD), including ID and autism spectrum disorder (ASD)(8–12), indicating pleiotropic effects. SSD- and broader NDD-risk CNVs were recently associated with lower cognitive ability in general population cohorts(13–15) and one SSD cohort(16). Additionally, NDD-risk CNVs were associated with childhood-onset SSD in one prior study(17), although this has yet to be replicated. Further studies are needed to clarify associations between known risk CNVs and severe phenotypes in SSD.

Beyond recurrent risk CNVs, broader genome-wide deletions may also contribute to severe phenotypes in SSD. In the Genome Aggregation Database (gnomAD) study of 14,891 individuals, rare structural variants, including deletions, were estimated to yield loss-of-function (LOF) effects on ∼5.5 genes per genome, accounting for ∼25-29% of rare, protein-truncating events per person(6). Global deletion burden scores (e.g., total number of deleted genes) have been weakly associated with cognitive functioning in some community samples(18,19). However, refining deletion scores with genomic annotations may clarify relationships. For example, recent studies in community samples(20–22) and one SSD cohort(16) found stronger associations with cognitive functioning when deletion burden was weighted by metrics capturing intolerance of affected genes to LOF, such as the “loss-of-function observed/expected upper bound fraction” (LOEUF) metric, which quantified the number of LOF variants per gene in gnomAD compared to the number expected given local sequence context and CpG methylation levels. For example, deletion of a LOF-intolerant gene (i.e., with an extreme LOEUF score) was associated with a decrease in general intelligence by 2.6 points(21). Identifying other annotations that refine CNV scores may improve power for patient stratification.

One potential strategy is to annotate CNVs by the neurodevelopmental pathway affected genes participate in. In particular, we previously derived and validated a set of 18 mutually-exclusive neurodevelopmental modules (i.e., gene-sets), spanning 17,216 genes, by applying weighted-gene co-expression network analysis to BrainSpan transcriptomic data from human brain samples spanning fetal development through early adulthood(23). Identifying these gene-sets leveraged the fact that genes are co-expressed in specific patterns to give cells their stable identity and drive biological processes, with expression patterns in the brain changing most dynamically during early development to drive the progression from cell proliferation to neuronal differentiation, synapse formation, and circuit refinement(24–26). These gene-sets were useful for identifying neurodevelopmental processes associated with SSD versus ASD. Specifically, genetic risk variants for SSD were enriched for gene-sets involved in modulating neuronal excitability and postnatal synaptic signaling and plasticity, as well as transcriptional regulation across development. ASD risk variants showed partial enrichment for these gene-sets but strong additional enrichment for gene-sets involved in early neuronal differentiation and in regulating the fetal gene expression changes that drive this early developmental process. The utility of annotating CNV burden scores by neurodevelopmental pathway for explaining severe phenotypes in SSD is unknown.

The present study therefore investigated whether severe phenotypes in SSD (i.e., borderline intellectual functioning and child-onset psychosis) are associated with known SSD- or NDD-risk CNVs, genome-wide deletion burden scores, or novel scores capturing deletion burden in each of our 18, previously validated, neurodevelopmental gene-sets. While global duplication burden weighted by LOF-intolerance was previously found to have a subtle effect on cognition(21), deletion burden weighted by LOF-intolerance had an over 3-fold greater effect(21), in line with LOF effects of exonic deletions.

Global and neurodevelopmental gene-set burden scores analyzed in the current study therefore focused on deletion burden scores. Primary analyses focused on a within-SSD case, phenotypic variability design. However, analyses for borderline intellectual functioning were extended to a sample of SSD relatives and controls to assess generalizability and replicability across healthy individuals and those at elevated risk for SSD but without a SSD diagnosis. Following associations between borderline intellectual functioning and fetal gene regulatory gene deletions, relationships between this gene-set and cortical morphology were explored in subjects with available structural magnetic resonance imaging (MRI) data. Replicability of primary findings was further assessed in the Adolescent Brain Cognitive Development (ABCD) Study^®^ cohort of over 11,000 youth.

## METHODS

### SSD-Focused Cohort

Data from 1,514 individuals, including 645 with a SSD, 253 relatives of a SSD patient, and 616 healthy controls, were harmonized from SSD-focused research clinics at UCLA, Feinstein Institute for Medical Research, and 9 sites of the North American Prodrome Longitudinal Study (NAPLS). Genetic ancestry of participants was diverse (Fig. S1). Studies were included if the protocol included DNA sample collection, structured clinical interviews for diagnosis, and standard cognitive measures.

SSD diagnoses and age of psychosis onset were determined using structured clinical interview. Child-onset SSD was defined as psychosis onset before age 13(17). Borderline intellectual functioning was defined as IQ estimate ≤85 (hereafter “borderline IQ”)(27). SSD-relative status was determined via the Family Interview for Genetic Studies (FIGS)(28), or because a family member with a SSD had been directly assessed. Specific inclusion/exclusion criteria differed between studies, but SSD-relatives and controls had no history of a psychotic disorder.

T1-weighted MRI data was available for a subset of participants. Reduced gray matter volume (GMV), surface area (SA), and cortical thickness (CT) are well-established in SSD(40); however, SSD patients show greater heterogeneity than controls(41), which may reflect variability in genetic risk profiles. Exploratory analyses therefore examined associations between these metrics and neurodevelopmental gene-set metrics of interest. MRI data was processed using standard Freesurfer pipelines. Age- and sex-normalized cortical morphology metrics (i.e., centile scores) were derived from population reference models(29), herein referred to as “BrainCharts”. See Supplementary Methods and Tables S1-2 for details.

### CNV Processing

CNVs were called from Illumina Global Screening Array data using PennCNV(30) and QuantiSNP(31) and processed via standard quality control (QC) procedures(7). CNVs identified by both algorithms, a minimum of 10 SNPs and 20 kilobases (kb) in length, and observed in <1% of the sample or gnomAD(6) were retained for analysis. CNVs identified as having ≥40% overlap with SSD-associated CNVs(7) were analyzed as known SSD-risk CNVs, and CNVs overlapping SSD-, ASD-(32), developmental disorder (DD-;33), or NDD-associated CNVs(9) were analyzed as broader NDD-risk CNVs (see Extended Table 1). CNVs were also annotated for any overlap with canonical RefSeq protein-coding gene exons(34). Total number of deleted genes and number of deleted genes within each of the 18 neurodevelopmental gene-sets (M1-M18) described above were calculated per subject. The LOEUF metric from gnomAD was used to calculate total deletion score per subject weighted by affected genes’ LOF-intolerance (i.e., “total deletion LOEUF sum”). A metric of connectivity for each gene within its assigned gene-set, “kWithin” score, was obtained using the intramodularConnectivity function from the WGCNA package in R, and was additionally used to calculate total deletion score per subject weighted by affected genes’ within-gene-set connectivity (i.e., “total deletion kWithin sum”).

Exploratory analyses were conducted for deletion LOEUF and kWithin sum per neurodevelopmental gene-set. See Supplementary Methods for details and Extended Table 2 for summary of neurodevelopmental gene-sets.

### Statistical Analyses

#### Demographic and Phenotypic Characteristics

Chi-square tests assessed differences between SSD, SSD-relative, and control groups in binary demographic characteristics. ANOVA controlling for sex and age of testing assessed group differences in continuous phenotypes (i.e., IQ estimate and MRI centiles), followed by Tukey pairwise comparisons as needed.

#### CNV Scores versus Phenotypic Characteristics

Logistic mixed models compared rates of rare deletions and duplications and presence of known SSD- or broader NDD-risk CNVs between SSD cases versus all non-cases (i.e., controls and SSD-relatives).

Logistic mixed models were then run within SSD cases to examine associations between child-onset psychosis or borderline IQ and 3 types of CNV scores: 1) presence of known SSD- or broader NDD-risk CNVs; 2) number of genes deleted per neurodevelopmental gene-set; and 3) global deletion burden scores, namely, total number of deleted genes, total deletion LOEUF sum, and total deletion kWithin sum. To minimize model convergence failures and low-confidence coefficient estimates, only gene-sets with ≥3 subjects with deletions affecting the gene-set were analyzed. All models included covariates for sex, 10 ancestry principal components (PCs), and a genetic relatedness matrix (GRM). Age at testing was an additional covariate for borderline IQ analyses. Analyses were run using the GENESIS package in R(35). Ancestry PCs and the GRM were generated using a state-of-the-art pipeline with PC-AiR(36) and PC-Relate(37), wherein PC-Relate calculates close genetic relatedness while accounting for ancestry PCs (i.e., to isolate relatedness due to family structure), and ancestry PCs are refined accounting for the GRM (i.e., to account for family-level genetic relatedness). *P*-values reported are two-sided and false discovery rate (FDR) correction was applied across genetic scores tested that achieved model convergence per severe SSD-relevant phenotype.

Following initial gene-set associations for borderline IQ in SSD cases, parallel analyses were run: 1) excluding SSD cases with known NDD-risk CNVs; 2) in non-cases only (i.e., SSD-relatives and controls); and 3) across SSD cases and non-cases without NDD-risk CNVs. Analyses incorporating non-cases included covariates for diagnostic group.

### ABCD Study Cohort

ABCD is a study of >11,000 diverse youth in the United States with clinical, cognitive, and neuroimaging data, and genotyping via Affymetrix Smokescreen array(38). Cognitive functioning estimates were available via the National Institutes of Health (NIH) Cognitive Toolbox at baseline, when youth were ∼9-10 years old. CNV calling and annotation procedures were similar to the primary SSD-focused cohort (see Supplementary Methods). A schematic of the main CNV scores analyzed and overall study analysis flow is shown in Fig. S2.

## RESULTS

### CNV Rates in SSD Cases vs. Non-Cases

Data from up to 617 SSD cases, 233 SSD-relatives, and 581 controls were analyzed following QC. See Table 1 for demographic, clinical, and CNV characteristics in the cohort. Rare, large CNVs (i.e., deletions or duplications >20 kb and 10 SNPs, with population frequency <1%) were identified in 536 SSD (86.9%), 202 SSD-relative (86.7%), and 489 control (84.2%) subjects.

**Table 1.** Demographic, clinical, and CNV burden characteristics across groups for included subjects.

|  | SSD Subjects |  | SSD-Relatives |  | Controls |  | 3-Group | 3-Group |
| --- | --- | --- | --- | --- | --- | --- | --- | --- |
|  | *N* or<br>*Mean* | % or<br>SD | *N* or<br>*Mean* | % or<br>SD | *N* or<br>*Mean* | % or<br>SD | Test Statistic | Test<br>p-value |
| *N* Subjects (% Female)* | 617 | 30.1%<sup>+</sup> | 233 | 55.8%<sup>^</sup> | 581 | 50.8%<sup>^</sup> | $\chi^2(df=2)=71.56$ | <0.001 |
| Self-Identified Race* | | | | | | | $\chi^2(df=10)=101.05$ | <0.001 |
| White | 280 | 45.3%<sup>+</sup> | 128 | 54.9%<sup>^</sup> | 336 | 57.8%<sup>^</sup> |  |  |
| Black/African American | 123 | 19.9%<sup>+</sup> | 16 | 6.9%<sup>□</sup> | 91 | 15.7% |  |  |
| Asian | 60 | 9.7% | 19 | 8.2% | 73 | 12.6% |  |  |
| American Indian / Alaska Native | 15 | 2.4%<sup>+</sup> | 27 | 11.6%<sup>^</sup> | 16 | 2.8%<sup>+</sup> |  |  |
| Native Hawaiian | 3 | 0.5% | 0 | 0% | 1 | 0.2% |  |  |
| >1/Other/Unknown | 139 | 22.5%<sup>□</sup> | 43 | 18.5% | 65 | 11.2%<sup>^</sup> |  |  |
| Hispanic/Latino* | 142 | 23.0%<sup>+</sup> | 77 | 33.0%<sup>^□</sup> | 118 | 20.3%<sup>+</sup> | $\chi^2(df=2)=18.19$ | <0.001 |
| *N* with Rare Deletions or Duplications | 536 | 86.9% | 202 | 86.7% | 489 | 84.2% | $W2=1.30$ | 0.523 |
| *N* with Rare Deletions<sup>&</sup> | 388 | 62.9% | 137 | 58.8% | 328 | 56.5% | $W2=5.28$ | 0.071 |
| *N* with Rare Genic Deletions*<sup>&</sup> | 197 | 31.9%<sup>+</sup> | 53 | 22.7%<sup>^</sup> | 147 | 25.3%<sup>^</sup> | $W2=9.93$ | 0.007 |
| *N* with Rare Duplications | 409 | 66.3% | 148 | 63.5% | 384 | 66.1% | $W2=0.58$ | 0.747 |
| *N* with Rare Genic Duplications | 323 | 52.4% | 105 | 45.1% | 289 | 49.7% | $W2=0.83$ | 0.659 |
| Mean IQ* | 99.1 | 16.0<sup>+□</sup> | 105.1 | 16.9<sup>^□</sup> | 110.9 | 13.6<sup>^+</sup> | $F(2,1391)=91.90$ | <0.001 |
| *N* Borderline IQ | 120 | 20.0% | 31 | 14.0% | 26 | 4.5% |  |  |
| Mean Age Cognitive Testing* | 23.82 | 10.28<sup>+□</sup> | 35.94 | 18.37<sup>^□</sup> | 24.54 | 12.27<sup>^+</sup> | $F(2,1395)=80.79$ | <0.001 |
| Mean Age Psychosis Onset | 19.93 | 5.54 | NA |  | NA |  |  |  |
| *N* Child-Onset | 39 | 6.4% |  |  |  |  |  |  |

\*Overall group difference $p < .05$ .
<sup>^</sup>Pairwise difference from SSD $p < .05$ .
<sup>+</sup>Pairwise difference from SSD Relative $p < .05$ .
<sup>□</sup> Pairwise difference from Controls $p < .05$
<sup>&</sup>Overall case versus non-case difference $p < .05$

The rates of having any rare CNV did not differ significantly between SSD cases compared to all non-cases (i.e., SSD-relatives and controls combined; OR=1.09, 95%CI[0.79,1.52], *p*=.60); however, compared to non-cases, SSD cases were more likely to have a rare deletion (OR=1.30, 95%CI[1.02,1.65], *p*=.030). SSD cases were also more likely to have rare deletions spanning genes compared to non-cases (OR=1.49, 95%CI[1.15,1.93], *p*=.002). Conversely, likelihood of having any rare duplication or any rare duplication spanning genes did not differ between SSD cases versus non-cases (OR=0.92, 95%CI [0.73,1.17], *p*=.50; OR=1.04, 95%CI[0.84,1.31], *p*=.69, respectively).

Known SSD- and broader NDD-risk CNVs were identified in 9 (1.5%) and 16 (2.6%) SSD individuals, respectively, compared to 0 (0.0%) and 1 (0.4%) SSD relatives, respectively, and 2 (0.3%) and 6 (1.0%) controls, respectively (see Extended Table 3). Compared to non-cases, SSD cases showed a significantly increased rate of SSD-risk CNVs (OR=7.04, 95%CI[1.47,33.80], *p*=.015), as well as broader NDD-risk CNVs (OR=3.98, 95%CI[1.55,10.20], *p*=.004).

Exploratory analyses comparing all three groups for overall CNV rates showed a similar pattern of results with no differences between controls and SSD-relatives (see Supplementary Results for details).

### Known Risk CNVs and Severe SSD-Relevant Phenotypes

Among the 39 SSD cases with child-onset psychosis, 2 (5.1%) had SSD-risk CNVs and 3 (7.7%) had NDD-risk CNVs compared to 7 (1.2%) and 13 (2.3%) of 568 later-onset cases, respectively. Although elevated in child-onset psychosis, these rates were not significantly different (OR =3.74, 95%CI[0.69,20.22], *p*=.125, *q*=.667; OR=2.97, 95%CI[0.74,11.86], *p*=.123, *q*=.667, respectively; Fig. 1A). However, exploratory analyses of known risk deletions versus duplications, separately, revealed a nominal association between NDD-risk deletions and child-onset psychosis (OR=4.33, 95%CI[1.03,18.19], *p*=.045; Fig. S4A). SSD-risk deletions showed a similar effect that was not significant, given reduced sample size (OR=4.34, 95%CI[0.77,24.31], *p*=.095). The model for NDD-risk duplications did not converge and only one SSD subject had a SSD-risk duplication, precluding analysis.

**Figure 1.**
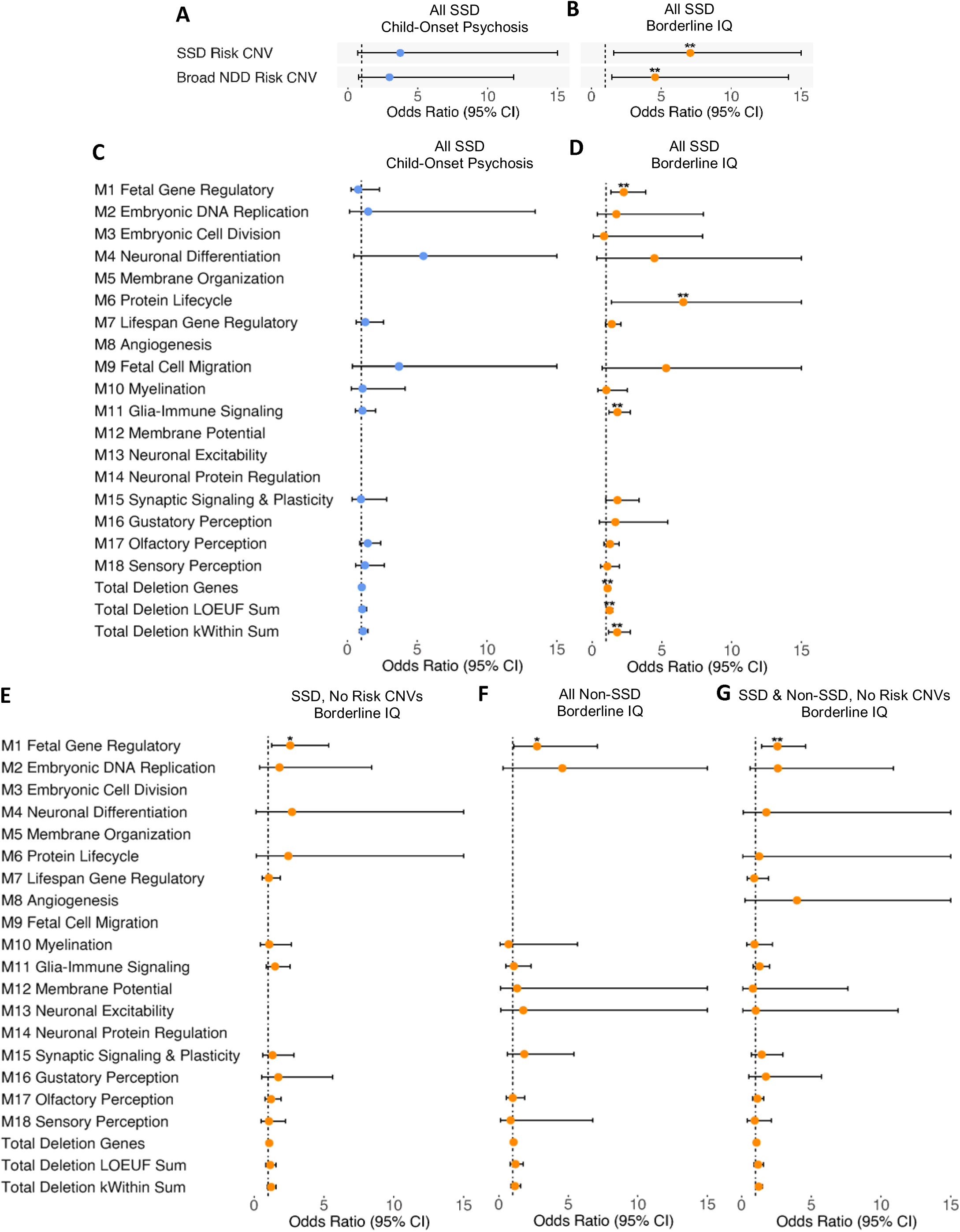
Odds ratio (OR) associated with presence of known risk CNVs for likelihood of (A) child-onset psychosis (*n* with child-onset psychosis = 39, *n* with later-onset psychosis = 568), and (B) borderline IQ (*n* with borderline IQ = 120, *n* without borderline IQ = 478) across all SSD cases, and for (C) child-onset psychosis and (D) borderline IQ associated with number of genes deleted per neurodevelopmental gene-set and global deletion burden scores. OR for number of genes deleted per neurodevelopmental gene-set and global deletion burden scores for (E) for SSD cases excluding those with known risk CNVs (*n* with borderline IQ = 113, *n* without borderline IQ = 469), (F) non-SSD subjects (*n* with borderline IQ = 57, *n* without borderline IQ = 741), and (F) SSD cases, SSD-relatives, and controls, excluding subjects with known risk CNVs (*n* with borderline IQ = 168, *n* without borderline IQ = 1210). \**p*<0.05; \*\**q*<0.05, corrected for number of CNV scores tested per analysis group. Max OR confidence interval shown=15; see Extended Table 4 for full statistics.

SSD- and NDD-risk CNVs were found in 5 (4.2%) and 7 (5.8%) of 120 SSD subjects with cognitive functioning in the borderline IQ or lower range versus 4 (0.8%) and 9 (1.9%) of 478 subjects with higher cognitive functioning. Both SSD-(OR=7.07, 95%CI[1.60,31.32], *p*=.01, *q*=.030) and NDD-risk CNVs (OR=4.56, 95%CI[1.48,14.10], *p*=.008, *q*=.030) were associated with borderline IQ (Fig. 1B). Exploratory analyses of known risk deletions and duplications separately suggested stronger effects of SSD-(OR=10.26, 95%CI[2.02,52.03], *p*=.005) and NDD-risk deletions (OR=4.88, 95%CI[1.40,17.05], *p*=.013) than NDD-risk duplications (OR=3.09, 95%CI[0.26,37.32], *p*=.38; Fig. S4B) for borderline IQ. Analyses in non-cases (i.e., SSD-relative and controls) suggested elevated rates of borderline IQ in individuals with NDD-risk CNVs, but with reduced power, this was not significant (OR=7.40, 95%CI[0.99,55.60], *p*=.052, *q*=.336). There were too few SSD-risk CNVs in non-cases for analysis.

Exploratory analyses using a more conservative definition of SSD-associated CNVs (i.e. surviving Bonferroni versus FDR correction in (7)), showed a similar pattern of association with borderline IQ and not childhood-onset SSD. An exploratory analysis of NDD-risk CNVs, excluding those also associated with SSD, showed a weaker, non-significant relationship with borderline IQ, reflecting both attenuated effect and reduced power, although associations were in the same direction. See Supplementary Results for detail.

### Neurodevelopmental and Global Deletion Burden Scores and Severe SSD-Relevant Phenotypes

Examination of relationships for childhood-onset SSD and global and neurodevelopmental gene-set deletion burden scores showed no significant associations, *p*s=*ns* (Fig. 1C).

However, borderline IQ in SSD cases was associated with deletion burden in the M1 fetal gene regulatory (OR=2.27, 95%CI[1.34,3.84], *p*=.002, *q*=.030; Fig. 1D), M6 protein lifecycle (OR=6.54, 95%CI[1.39,30.75], *p*=.017, *q*=.039), and M11 glia and immune signaling gene-sets (OR=1.82, 95%CI[1.21,2.74], *p*=.004, *q*=.030), as was total number of deleted genes (OR=1.11, 95%CI[1.03,1.20], *p*=.007, *q*=.030), total deletion LOEUF sum (OR=1.25, 95%CI[1.05,1.50], *p*=.013, *q*=.035), and total deletion kWithin sum (OR=1.30, 95%CI[1.07,1.58], *p*=.007, *q*=.030).

Importantly, SSD- and NDD-risk CNVs are generally large, multi-gene CNVs(7,32,39), and often affect M1, M6, and M11 genes, as well as LOF-intolerant genes (see Extended Table 3). To determine whether associations exist beyond effects of known risk CNVs, analyses were re-run excluding SSD subjects with known NDD-risk CNVs. Only M1 fetal gene regulatory deletions remained associated with borderline IQ (OR=2.59, 95%CI[1.25,5.34], *p*=.010, *q*=.143; Fig 1E), with 9 (8.0%) of the remaining 113 SSD cases with borderline IQ carrying a M1 deletion, compared to 12 (2.6%) of 469 cases without borderline IQ. Although this did not survive multiple-testing correction, the effect was similar, suggesting this was due to reduced power rather than reduced effect. Extending analyses to non-SSD cases also revealed a nominal association between M1 deletions and borderline IQ (OR=2.75, 95%CI[1.07,7.10], *p*=.036, *q*=.336; Fig. 1F). Maximizing statistical power across SSD cases and non-cases without known risk CNVs revealed a significant association for M1 deletions and borderline IQ (OR=2.57, 95%CI[1.44,4.60], *p*=.002, *q*=.025; Fig. 1G).

Exploratory analyses for neurodevelopmental gene-sets weighted by LOEUF score (Extended Table 5, Fig. S6) or kWithin connectivity score (Extended Table 6, Fig. S7) showed a similar pattern of results, with slightly more robust associations between borderline IQ and M1 kWithin sum compared to M1 LOEUF sum. Furthermore, restricting analyses to 438 SSD subjects with narrow schizophrenia or schizophreniform diagnoses revealed highly similar associations for borderline IQ and child-onset psychosis, indicating that this pattern exists across the psychosis spectrum (Extended Table 7, Fig. S8). Controlling for differences in cognitive assessment type, DNA tissue source, or genotyping batch also showed similar results (Extended Tables 8-10). Furthermore, controlling for the number of genes deleted in all gene-sets other than the primary gene-set of interest showed highly similar results (Extended Table 11, Fig. S9), with the exception that most individual gene-sets were not associated with borderline IQ when subjects with NDD-risk CNVs were included in the analysis. However, M1 deletion burden remained associated with borderline IQ, controlling for non-M1 deletion burden, when subjects with NDD-risk CNVs were excluded from the analysis. Using a Bonferroni-correction for the number of independent neurodevelopment gene-sets annotated (i.e., .05/18 = .0278) to interpret gene-set associations yields similar conclusions. See Supplementary Methods and Results for detail.

Thus, beyond known NDD-risk CNVs, deletion of M1 fetal gene regulatory genes increased likelihood of borderline IQ across SSD cases and non-cases.

### M1 Fetal Gene Regulatory Deletions and Cortical Morphology

Approximately half the cohort had QC-passing T1 structural MRI data (SSD *n=*284, SSD-relative *n*=105, Control *n*=317). Groups differed significantly across GMV (*F*(2,682)=11.50, *p*=.00001), CT (*F*(2,682)=10.92, *p*=.00002), and SA centiles (*F*(2,682)=4.14, *p*=.02; Fig. 2A-C). SSD cases showed reduced centiles across metrics compared to controls (*p*s<.027). Relatives were intermediate, differing from controls for GMV (*p*=.008), but not CT (*p*=.49) or SA centile (*p*=.12), nor from SSD subjects for any metric (*p*s>.10).

**Figure 2.**
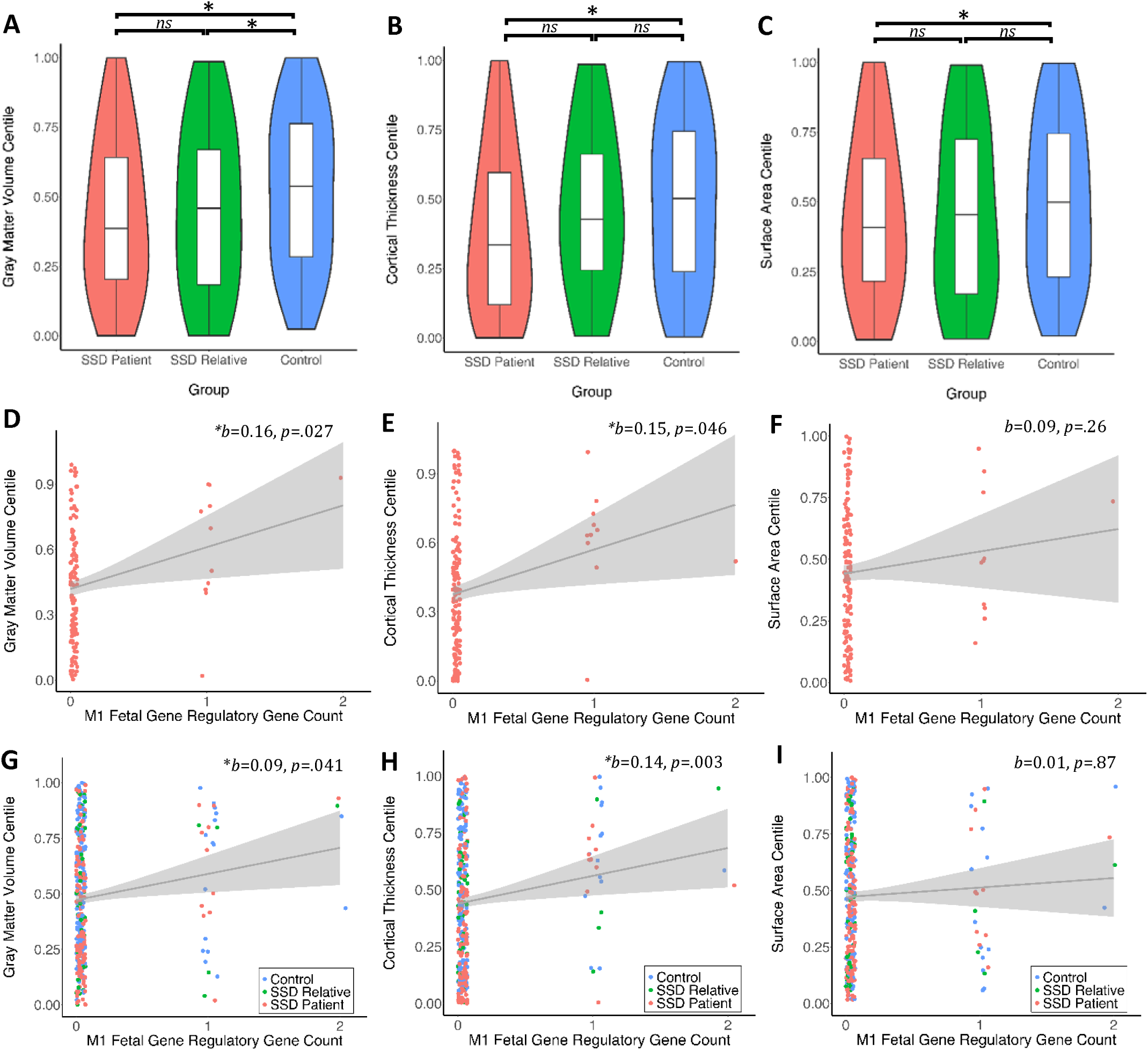
Group differences in gray matter volume (A), cortical thickness (B), and surface area (C) centiles. Excluding subjects with known risk CNVs, relationships between M1 deletions and cortical centile scores for (D-F) SSD subjects (*n* = 277) and (G-I) across SSD cases and non-cases (*n* SSD-relatives = 105, *n* Controls = 313). To facilitate interpretability, relationships are plotted using raw centile and gene count values; reported effect sizes and p-values are from the mixed models. \**p*<0.05.

Excluding subjects with known NDD-risk CNVs, which often result in macrocephaly or microcephaly(42), M1 deletions were associated with significantly increased GMV (*b*=0.16, 95%CI[0.02,0.29], *p*=.027) and CT in SSD cases (*b*=0.15, 95%CI[0.003,0.31], *p*=.046), but not SA centile (*b*=0.09, 95%CI[-0.06,0.23], *p*=.26; Fig. 2D-F). Extending analyses to all cases and non-cases without known risk CNVs showed associations for GMV (*b*=0.09, 95%CI[0.004,0.17], *p*=.041) and CT (*b*=0.14, 95%CI[0.05,0.24], *p*=.003), but not SA centile (*b*=0.01, 95%CI[-0.08,0.10], *p*=.87; Fig. 2G-I).

Analyses including tissue type, genotyping batch, or non-M1 deletion burden as covariates in models were largely similar, although some associations were no longer significant controlling for batch. See Supplementary Results for details.

Exploratory analyses of M1 deletions weighted by LOEUF score were not significantly associated with brain centiles in this half of the sample (*p*s>.05, Fig. S10); however, M1 deletions weighted by kWithin score were associated with increased CT centile across SSD cases and non-cases (*b*=0.14, 95%CI[0.001,0.29], *p*=.048; Fig. S11).

### Replication in ABCD

In ABCD, 9,930 youth had QC-passing CNV calls. Detection of rare, large CNVs was lower in the ABCD study cohort (i.e., 28.54% with deletions and 35.52% with duplications; see Table S4) than in the primary SSD-focused cohort (i.e., 59.61% with deletions and 65.76% with duplications), likely reflecting noisier Affymetrix array data.

Rates of borderline IQ were significantly elevated in carriers of NDD-risk CNVs (OR=1.82, 95%CI[1.20,2.74], *p*=.005, *q*=.018) and non-significantly elevated for SSD-risk CNVs (OR=1.56, 95%CI[0.87,2.79], *p*=.14, *q*=.16; Fig. 3). Borderline IQ was not significantly associated with M1 gene deletions when subjects with NDD-risk CNVs were included (OR=1.22, 95%CI[0.98,1.51], *p*=.072, *q*=.12), partially reflecting effects of one subject with a 22q11.2 deletion with an extreme number of deleted M1 genes who was just above the borderline IQ range (see Supplementary Results); however number of deleted M1 genes was nominally associated with borderline IQ excluding subjects with NDD-risk CNVs (OR=1.33, 95%CI[1.00,1.76], *p*=.049, *q*=.10). M1 deletions weighted by LOF-intolerance were nominally associated with borderline IQ when subjects with NDD-risk CNVs were included in analyses (OR=1.51, 95%CI[1.06,2.16], *p*=.023, *q*=.061), and were significantly associated excluding subjects with NDD-risk CNVs (OR=2.07, 95%CI[1.27,3.37], *p*=.004, *q*=.018). M1 deletions weighted by kWithin score were not associated with borderline IQ, including (OR=1.33, 95%CI[0.91,1.94], *p*=.14, *q*=.16) or excluding subjects with NDD-risk CNVs (OR=1.34, 95%CI[0.83,2.16], *p*=.23, *q*=.23).

**Figure 3.**
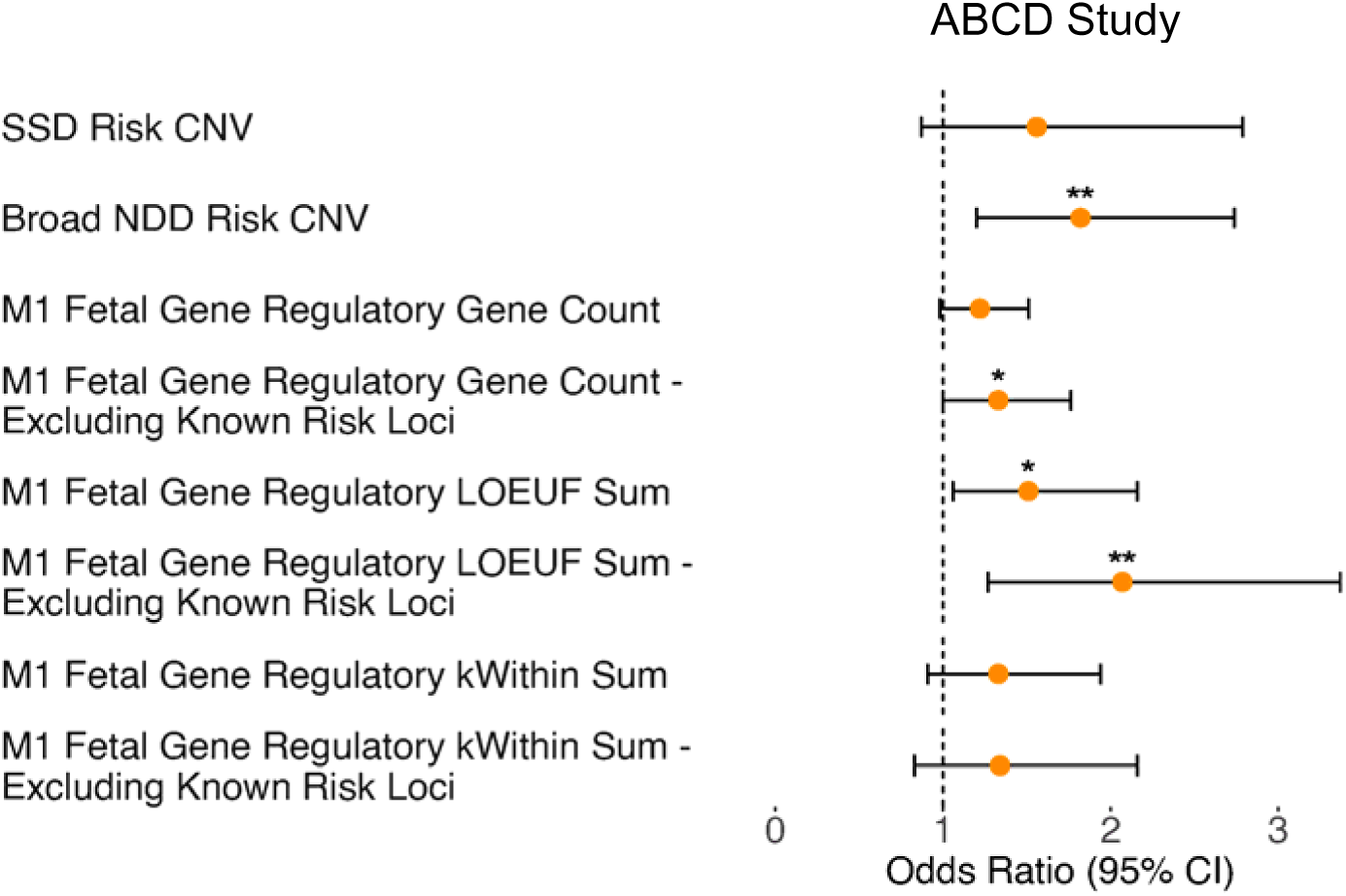
Odds ratio (OR) for borderline intellectual functioning in ABCD study subjects associated with SSD- and broad NDD-risk CNVs, number of M1 genes deleted, M1 deletion burden weighted by LOEUF score, and M1 deletion burden weighted by kWithin score, including (*n* = 9,930) or excluding subjects (*n* = 9,776) with known risk CNVs. \**p*<0.05; \*\**q*<0.05, corrected for number of scores tested.

In 9,378 youth with QC-passing MRI data, excluding subjects with NDD-risk CNVs, M1 deletion burden was not associated with centile scores in a consistent direction, *p*s=*ns* (Fig. 4A-C), but was associated with deviance in SA centile (i.e., both larger and smaller SA; *b*=0.02, 95%CI[0.001,0.03], *p*=.041, Fig. 4F). Furthermore, M1 deletions weighted by LOF-intolerance were associated with lower GMV (*b*=-0.06, 95%CI[-0.11,-0.003], *p*=.038; Fig. 4G) and SA centiles (*b*=-0.06, 95%CI[-0.11,-0.006], *p=*.029; Fig. 4I), as well as increased deviance in CT centile (*b*=0.04, 95%CI[0.01,0.07], *p*=.005, Fig. 4K). M1 deletion burden weighted by kWithin connectivity was associated with increased deviance in CT centile (*b*=0.023, 95%CI[0.001,0.05], *p*=.040, Fig. 4N), but not with other MRI metrics. Thus, in late childhood, M1 deletions weighted by LOF-intolerance were associated with smaller age-normalized GMV and SA and M1 deletions weighted by LOF-intolerance and kWithin connectivity were associated with deviation in age-normalized CT in both directions.

**Figure 4.**
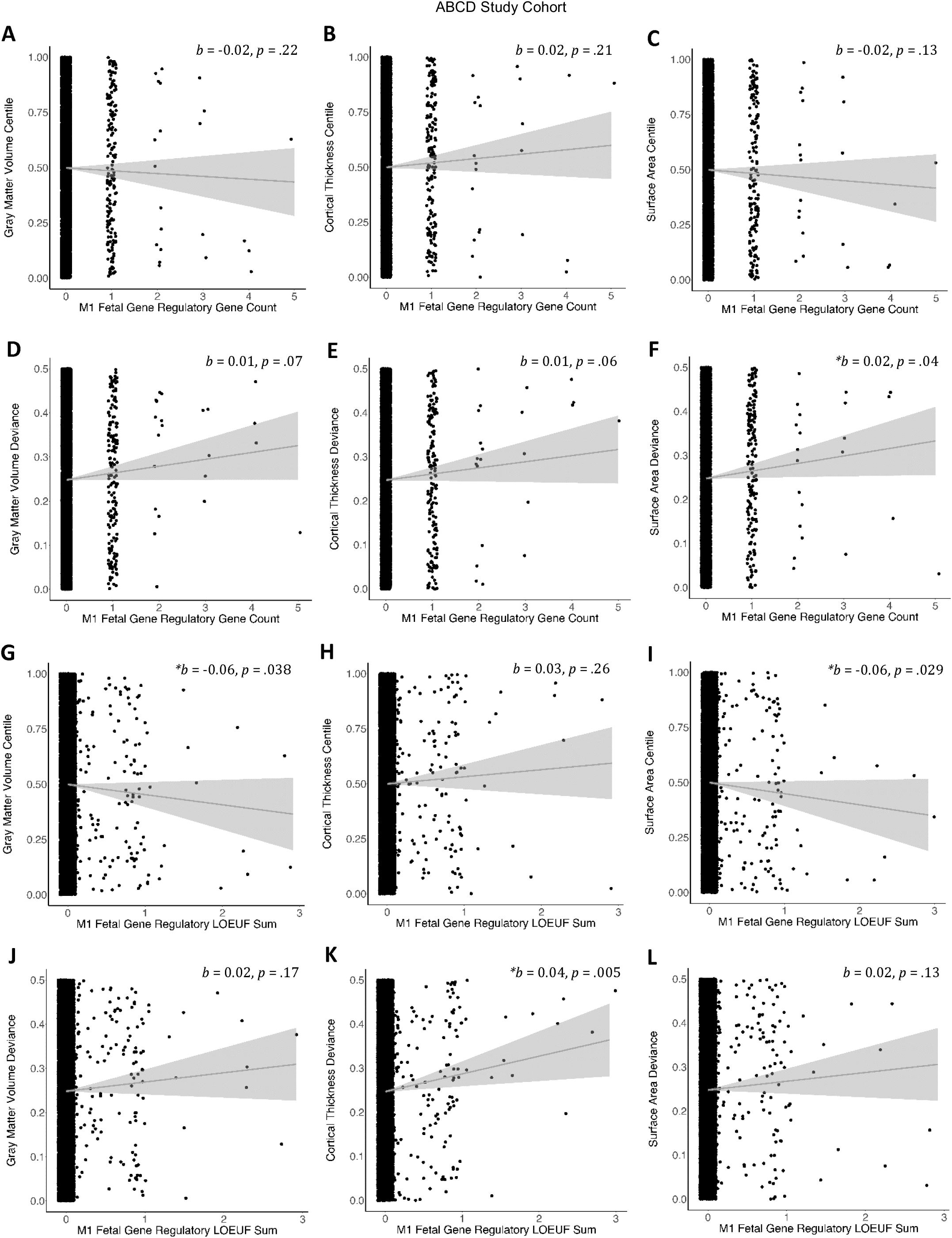
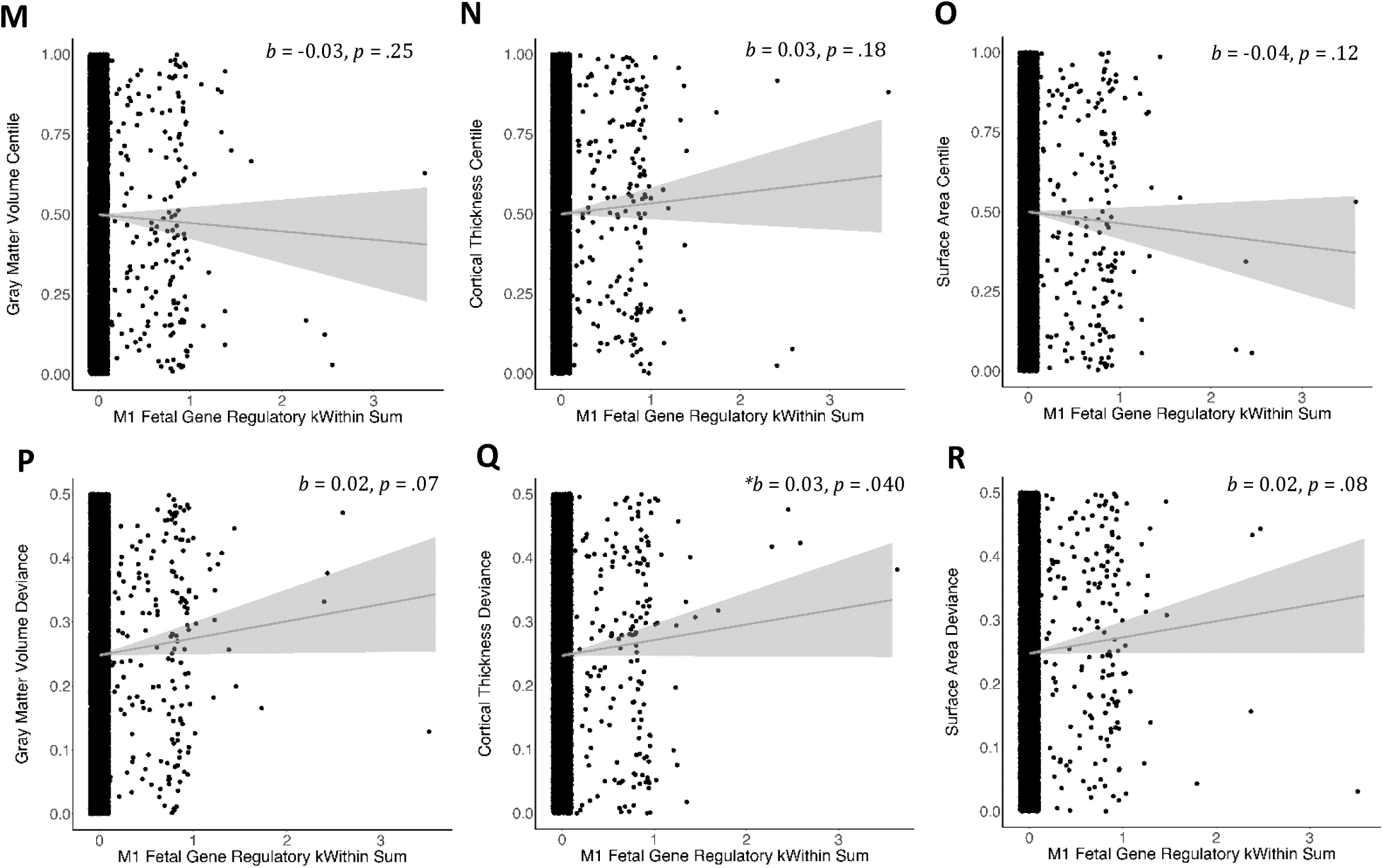
Associations between M1 deletion gene count and (A) gray matter volume, (B) cortical thickness, or (C) surface area centile or deviance in these centile scores (D-F) in ABCD study subjects without known risk CNVs (*n* = 9,378). Parallel associations between M1 LOEUF score and (G) gray matter volume, (H) cortical thickness, or (I) surface area centile or deviance in these centile scores (J-L). Parallel associations between M1 kWithin score and (M) gray matter volume, (N) cortical thickness, or (O) surface area centile or deviance in these centile scores (P-R). Relationships are plotted using raw centile and M1 scores; reported effect sizes and p-values are from the mixed models. \**p* < 0.05.

Using a more conservative set of CNV calls for ABCD from (43), that were a minimum of 20 SNPs and 50 kb in length, yielded a similar pattern of results for borderline IQ (Fig. S19), whereas results for centiles were not significant (Fig. S20; see Supplementary Methods and Results for details). Models controlling for tissue type and batch showed similar effects for borderline IQ and MRI metrics (Table S7). In models for M1 deletion burden controlling for non-M1 deletion burden, results for borderline IQ were no longer significant, while CT centile deviance remained associated with M1 deletions weighted by LOF-intolerance, and SA centile deviance remained associated with number of deleted M1 genes (see Supplementary Results; Table S7). Difficulty isolating M1 effects may reflect noisier signal intensity data in ABCD, where the typical deletion called was larger in size (Fig. S17) compared to the primary SSD-focused cohort (Fig. S3), despite selective pressure against larger CNVs.

## DISCUSSION

The present study investigated relationships between severe SSD-relevant phenotypes and CNV burden scores in two cohorts and found, for the first time, that deletion of fetal gene regulatory genes was associated with severe phenotypes in SSD individuals and the broader population. In line with prior studies, known SSD- and NDD-risk CNVs were associated with impaired cognitive function in SSD, while NDD-risk deletions were nominally associated with child-onset psychosis. Importantly, beyond known risk CNVs, deletions impacting genes that coordinate the fetal gene expression changes that drive early neuronal development in the brain also increased risk for poor cognitive functioning and were associated with altered cortical morphology. Associations were observed across SSD cases and non-cases, and in the ABCD youth cohort. Results demonstrate the utility of a neurodevelopmental framework for identifying biological processes that shape phenotypic variation and implicate genetically-mediated disruptions to early brain development in severe SSD-relevant phenotypes, in particular.

Established risk CNVs for SSD and NDDs are often large, multi-gene CNVs and span genes that are intolerant to LOF(7,32,39). Our observation that 1.5% and 2.6% of SSD patients carried an established SSD- or NDD-risk CNV, respectively, is in line with prior estimates(7,16). Furthermore, the nominal association observed between child-onset psychosis and NDD-risk deletions partially replicates the one prior study that examined NDD-risk CNVs (considering deletions and duplications together) in child-onset psychosis and found elevated rates(17). Our finding suggests that NDD-risk deletions may confer stronger risk for child-onset SSD than duplications; however, this requires replication. Known risk CNVs were also associated with borderline IQ in SSD cases, with similar but attenuated effects in non-cases. This is consistent with pleiotropic associations of many of these CNVs with ID and DD(10) and recent associations with cognitive functioning in community samples(13,19). As genetic diagnoses can facilitate access to specialized services(44) and child-onset and poor cognitive function in SSD are both associated with a more chronic course of illness, this suggests that genetic testing for CNVs could be useful when a SSD patient presents with childhood-onset or poor cognitive functioning. However, as only 7.7% of child-onset SSD cases and 5.8% of SSD cases with borderline IQ had a NDD-risk CNV, this highlights the importance of other potential factors, such as other damaging variants.

Importantly, our novel neurodevelopmental annotation of CNV scores identified that deletion of fetal gene regulatory genes is also associated with borderline IQ and altered cortical morphology in SSD cases and non-cases. Associations with borderline IQ remained after excluding subjects with known risk CNVs, while associations for total number of deleted genes and total LOEUF sum did not. This suggests that this gene-set annotation may explain more variance in cognitive function that is independent from effects of known NDD-risk CNVs compared global LOF-intolerance scores. Similar effects were observed in the ABCD cohort, particularly when M1 deletions were weighted by LOF-intolerance, which is consistent with prior findings in community samples that global CNV burden scores weighted by LOF-intolerance are more strongly associated with cognitive performance than unweighted scores(20–22). Overall attenuated CNV effects in ABCD may reflect lower quality CNV calls, differences in the specific genes deleted in the SSD-versus ABCD cohort, and/or lower heritability of cognitive functioning in childhood (h^2^=∼0.4) compared to adulthood (h^2^=∼0.80)(45).

Associations between M1 deletion burden and altered cortical morphology in both directions across cohorts are consistent with a recent finding that high LOEUF-based CNV scores were associated with extreme neuroanatomic centiles, in either direction, in the Philadelphia Neurodevelopmental Cohort, and effects of known pathogenic CNVs on cortical morphology in both directions(20). Interestingly the average increase in GMV and CT centile associated with each deleted M1 gene in SSD patients (i.e., 0.15 and 0.16 on centile scale, respectively) was larger than the overall group difference for SSD patients versus controls in the reverse direction in the current study (i.e., −0.099 and −0.114, respectively) and similar to the magnitude of SSD versus control GMV and CT differences in the reverse direction in the original BrainChart paper (i.e., −0.165 and −0.167, respectively, 29). Although the opposing direction of M1 gene deletions on cortical morphology in SSD is not immediately intuitive, this is similar to effects of multiple known risk CNVs for SSD, such as 22q11.2 deletions(46) and 15q11.2 BP1-BP2 deletions(47), which are associated with increased cortical thickness and also increase risk for intellectual disability and broader NDDs. While M1 deletion effects on cortical morphology were attenuated in the younger ABCD cohort, overall, these findings suggest that deletion of some M1 genes can substantially alter cortical morphology.

Genes in the M1 fetal gene regulatory gene-set are enriched for fetal-specific expression and hub genes include the histone demethylase enzyme, *KDM5B*, and transcription factors, *SOX11* and *SOX4,* known to critically regulate neurogenesis, neuronal differentiation, and dendritic morphogenesis(48,49). This gene-set was also previously found to be strongly enriched for risk variants for ASD(23). While these M1 hub genes were not deleted in subjects in the current study, deleted M1 genes in SSD patients with borderline IQ outside known risk CNVs included *SEMA3C*, *ZNF568, MACROD2*, and *CDKAL1*. Each of these genes showed their highest expression during early-mid fetal development (Fig. S21), as did M1 genes of potential interest in the ABCD Study cohort (Fig. S22).

*SEMA3C* encodes a glycoprotein that is secreted as an axonal guidance cue for developing neurons and facilitates nervous-system patterning during embryonic development(50). *ZNF568* is a transcription factor essential for maintaining the neural stem cell pool during fetal development and is associated with brain size in mice and humans(51–53). *MACROD2* encodes a deacetylase involved in DNA repair and chromatin structure that is expressed predominantly during fetal brain development(54) and is a candidate ASD-risk gene(55). *CDKAL1* encodes a brain-expressed transfer RNA modifying enzyme involved in protein translation(56). Follow-up studies are needed to clarify the function of these genes. Nevertheless, these findings suggest that deletions of genes involved in orchestrating the gene expression changes that shape early neuronal development are important contributors to altered cognitive functioning and brain development for a subset of SSD patients and the broader population.

Several limitations should be noted. First, psychotic diagnoses in the SSD-cohort were broad, which could result in noisier genotype-phenotype associations. However, including broader diagnoses offers opportunities to understand relationships across the diagnostic spectrum and associations in subjects with narrow schizophrenia or schizophreniform diagnoses showed a similar pattern of effects. Second, phenotyping methods differed across sub-studies in the primary SSD-cohort and MRI data was included from multiple sites and scanners. Post-hoc harmonization was necessary to generate a cohort with sufficient statistical power for within-case analyses but may have contributed noise to our models. Nevertheless, we were able to replicate associations between key CNV scores and cognitive and cortical metrics across non-cases and to some extent, in the younger ABCD cohort, providing support across assessment protocols. Finally, we had limited power to detect associations between clinical variability and deletions affecting small neurodevelopmental gene-sets. Relatedly, while the neurodevelopmental gene-sets used to annotate CNVs were previously validated for shared biological function(23), exploring deletion burden in other literature-based or gene co-expression-based gene-sets may yield further insights into phenotypic variability among SSD patients and general population samples. Analyses in larger datasets that are better powered for investigating small gene-sets and can accommodate higher multiple-testing correction are necessary to identify the optimal biological parcellation of CNV scores for patient stratification.

In summary, the current study confirmed contributions of known risk CNVs to severe phenotypes in SSD, and beyond known risk CNVs, showed for the first time that deletion of genes involved in regulating the gene expression changes that orchestrate early neuronal development increase risk for poor cognitive function and altered cortical morphology in SSD and the broader population. Implication of a fetal-specific mechanism for a subset of SSD subjects with borderline intellectual functioning may help explain why treatment initiation after psychosis onset, (i.e., usually in adulthood) has lower efficacy in patients with poor cognitive function(57,58). Developing interventions that can be administered earlier in development may be necessary to optimize outcomes for these patients. Results highlight the utility of a neurodevelopmental framework for explaining clinical heterogeneity in SSD and offer a promising direction for nominating specific genes that may contribute to severe phenotypes for future investigation.

## Disclosures

RAIB, JS and AFA-B are co-founders of Centile Bioscience. RAIB and JS additionally serve as directors for Centile Bioscience Inc. and Ltd.

## Data Availability

Data used in the current student that was collected under informed consent procedures consistent with broad sharing of data are available through the NIH Data Archive Collection #3226

## Acknowledgements

The authors thank Evan Eichler for his feedback and advice on analyses. This work was supported by NIMH (K08 MH118577 to JKF; MH101506 to KHK; R01 MH37705, R01 MH110544 and P50 MH066286 to KHN; U01 MH082004 to DOP; U01 MH082004 to TDC; U01 MH081988 to EFW; R37 MH085953, U01 MH081902, P50 MH066286, R01 MH129858, U01 MH124639, R01 MH123575, and U01 MH119736 to CEB; F31 MH119786 to CKD; F31 MH124421 to AG), the Brain & Behavior Research Foundation (NARSAD Young Investigator Award to JKF), the National Center for Advancing Translational Sciences UCLA CTSI Grant UL1TR001881 (Award to CEB & JKF), the UCLA Brain Research Institute (Postdoctoral Award to JKF), and the Shear Family Foundation. Supplementary Figure 2 (i.e., the schematic of analyses) was created using images from Flaticon.com.

## Notes

### Author Declarations

The IRB of the University of California, Los Angeles gave ethical approval for this work.

### Summary of Updates

To include additional sensitivity analyses and results following initial review. The primary results remain the same.

