## Supplementary Information for "Fetal Gene Regulatory Gene Deletions are Associated with Poor Cognition in Schizophrenia and Community-Based Samples"

**Supplementary Methods**

**Clinical, Cognitive, and MRI Data in the SSD Cohort**

Studies were included in the current study if the original protocol included collection of DNA samples from SSD patients without a known genetic syndrome, structured clinical interviews for diagnosis (i.e., the Structured Clinical Interview for DSM Disorders (SCID) versions III-R, IV(1), or 5(2), or the Kiddie Schedule for Affective Disorders and Schizophrenia (KSADS)(3)), and standard cognitive measures. Recruitment foci of included studies included recent-onset adult SSD patients(4–9), children and adolescents with SSD(10–12), adults with chronic SSD(13), and youth aged 12-30 years at clinical high risk for SSD who were followed longitudinally for up to ~2 years to identify individuals who developed a SSD(14, 15). Self-reported racial and ethnic identities of subjects was diverse, in line with the urban setting of most study sites. See Table S1 for details. Subjects were included in the primary analyses as a SSD case if they had or developed a diagnosis of schizophrenia (*n*=345), schizophreniform disorder (*n*=93), schizoaffective disorder (*n*=49), psychotic disorder not otherwise specified (*n*=75), bipolar disorder with psychotic features (*n*=20), major depressive disorder with psychotic features (*n*=8), brief psychotic disorder (*n*=1), delusional disorder (*n*=6), or unspecified SSD (*n*=20). Subjects with diagnoses of psychotic disorder due to a general medical condition (*n*=3) or substance-induced psychotic disorder (*n*=1) were excluded. The DNA sample for 1 additional subject was excluded because insufficient phenotypic data was available to allow them to be included in analyses. The only exclusion criteria applied for control and SSD-relative subjects for the current analyses was that they had no history of a psychotic disorder. Other inclusion and exclusion varied by study. For example, some studies did not complete full assessment batteries on subjects with a history of head injury or current or premorbid IQ estimates <70, or excluded these subjects from analyses in prior publications from the study (e.g., NAPLS 2 & 3). However, given the focus of the present study on examining relationships between genetic heterogeneity and clinical heterogeneity among SSD individuals, these subjects were included in the present analyses if DNA samples and clinical, cognitive, and/or MRI data was available and considered valid. The distribution of IQ estimates and age of psychosis onset for SSD subjects are shown in Fig. S8.

IQ estimates were based on the most comprehensive measure available per subject, derived from the Wechsler Adult Intelligence Scale (WAIS)(16), the Wechsler Abbreviated Scale of Intelligence (WASI)(17), the Measurement and Treatment Research to Improve Cognition in Schizophrenia (MATRICS) Consensus Cognitive Battery(18), or the Wechsler Test of Adult Reading (WTAR)(19). For MATRICS battery-based IQ estimates, a linear regression was run on a subset of 191 subjects with both MATRICS and WASI scores available to generate a formula for weighting subtest scores, which was then applied to the subset of individuals for whom only MATRICS scores were available. If multiple comprehensive IQ scores (i.e., including both verbal and performance-based subtests) were available for subjects, the score nearest to psychosis onset was used. Thus, for studies involving SSD subjects who already had a psychotic diagnosis, the IQ estimate from the first assessment was used, whereas for studies involving clinical-high risk for SSD subjects who were followed longitudinally (i.e., NAPLS 2 and 3), the IQ estimate from the last available assessment was used.

T1-weighted structural MRI data was processed with Freesurfer version 7.1.1 followed by standard QC procedures(20), including visual inspection of cortical parcellations and exclusion of poor quality scans. Global cortical metrics for total gray matter volume (GMV), average cortical thickness (CT), and total surface area (SA) were extracted. Scan protocols varied by study. Protocols included in the current study are listed in Table S2. Combat from the SVA R package, which uses an empirical Bayes framework, was first used to adjust for scanner/protocol effects on neuroanatomic metrics(21), with covariates for diagnostic group, sex, and age included in the models for GMV and SA, and age^2^ additionally included as a covariate for CT. Individual subject-level centile scores were then generated relative to age- and sex-normative trajectories using the BrainCharts tool for GMV, SA, and CT(22). Subjects were included in analyses if a minimum of 10 controls with usable structural MRI data was available to account for scanner/protocol effects with Combat and during the generation of centile scores. To maximize the number of scanners/protocols with at least 10 control subjects for scanner/protocol normalization, T1 data from controls was included in the normalization steps regardless of whether DNA samples and CNV calls were available for these subjects. Similar to IQ estimates, if multiple QC-passing T1 scans were available per subject, the scan closest to psychosis onset was analyzed, such that studies involving SSD subjects who already had a psychotic diagnosis used the first available scan, whereas studies involving clinical-high risk for SSD subjects (i.e., NAPLS 2 and 3) used the last available scan.

All individual studies obtained Institutional Review Board (IRB) approval from their local IRB during data collection and written informed consent from subjects, and the harmonization study protocol was approved by the UCLA IRB.

**CNV Calling, Quality Control, and Annotation**

CNV calling followed standard pipelines(23–25). Briefly, DNA was extracted from whole blood, saliva, or buccal samples, followed by genome-wide genotyping at 700,078 sites using the Illumina Global Screening Array in two batches. CNVs were called in autosomal chromosomes using PennCNV(26) and QuantiSNP(27), with the “hhall.hmm” and “levels-hd.dat” hidden markov models specified, respectively, to identify CNVs, and GC content correction. CNV calls were iteratively merged if the gap between neighboring calls was <20% of the total length using the PennCNV clean_cnv.pl script. CNVs were retained if they were concordant in direction (i.e., deletion or duplication), showed ≥50% overlap between PennCNV and QuantiSNP, had a minimum PennCNV confidence score and QuantiSNP Log Bayes Factor score of 10, with at least one algorithm with a confidence score ≥15, and were a minimum of 10 SNPs and 20 kilobases (kb) in length. CNVs overlapping ≥50% with centromeres, telomeres, or segmental duplications were excluded. Rare CNVs were defined as those with <50% overlap of any CNV found with a population maximum frequency ≥1% in gnomAD(28) or regions affected by deletions or duplications within each batch of the sample at ≥1%. Deletion and duplication loci rates within the sample were defined using the populationRanges function from the CNVRanger package in R. Samples with non-matching self-reported versus genetically-imputed sex (*n*=6), identified as duplicates (*n*=11), <98% genotyping rate (*n*=26), or outliers (>3 SD from median) in Log R Ratio SD (LRR SD), absolute waviness factor, B Allele Frequency standard deviation (BAF SD), or total deletion or duplication length (*n*=30) or with total CNV length > 15 Mb (*n* = 5) were excluded.

QC-passing CNVs were annotated for overlap with known risk CNVs for SSD, ASD, developmental disorders (DD), or NDD (see Extended Table 1). Given that deletions versus duplications at the same locus can have different associations with SSD, ASD, DD, and/or NDD, identification of disease-associated loci considered deletions and duplications separately. Thus, CNVs associated with SSD were obtained from Marshall et al., 2017(23), and were defined as deletions (“loss”) and/or duplications (“gain”) associated with SSD at the Benjamini-Hochberg False Discovery Rate (BH-FDR) *q* < 0.05 level in Table 1 of (23). CNVs associated with ASD were defined as deletions and/or duplications associated with ASD in Sanders et al., 2015(29) (i.e., primary CNV direction of effect with FDR *q* < .05 in Table 2 of (29) based on combined the Simons Simplex Collection (SSC) + Autism Genome Project (AGP) cohort). CNVs associated with DD were a list of 66 deletions and duplications downloaded May 2020 from <https://www.deciphergenomics.org/>(30). CNVs associated with broader NDDs were defined as deletion or duplication syndromes associated with NDDs at the FDR *q* < .05 threshold in Table 1, Table S2, and Table S3 in Coe et al., 2014(31). Compiled CNVs were lifted over to hg38 as needed (see Extended Table 1). Disorder-associated CNVs were identified if a QC-passing CNV call overlapped ≥40% with one of these known loci in a convergent direction (i.e., deletion or duplication), except for NRXN1 deletions, which were called if any NRXN1 exon was overlapped by a deletion(32,33).

QC-passing CNVs were then annotated for overlap with any exons(23) of protein-coding genes in RefSeq(34). Total number of genes with one or more exons spanned by deletions and number of genes with exons spanned by deletions for each of the 18 mutually-exclusive BrainSpan neurodevelopmental gene-sets described above (see [Extended Table 2](https://docs.google.com/spreadsheets/d/1n6bs16wPe7NGT_k2_vmZ9DSKTfLooBKq5oYplpaSPyI/edit?usp=sharing) for detailed summary) were summed to generate total and gene-set-specific deletion scores for each subject, respectively. Additionally, the “loss-of-function observed/expected upper bound fraction” (LOEUF) metric from gnomAD, describing the ratio of observed versus expected LOF variants in 141,456 humans per protein-coding gene, was converted to percentiles across genes, ordered such that genes with higher percentiles had lower LOEUF scores (i.e., percentile for 1 - LOEUF, where higher LOEUF scores in the current paper indicate greater mutational constraint)(35). For each subject, LOEUF percentiles were summed across all deletion-spanning genes to generate a total deletion LOEUF sum score, capturing total deletion burden weighted by affected genes’ LOF-intolerance. Similarly, a metric of within-gene-set connectivity was extracted from the WGCNA package in R, the “kWithin” score, which represents the sum of the weights of all connections (i.e., edges or correlations) between a given gene and all other genes assigned to the same module/gene-set. To create a normalized connectivity score parallel to the total LOEUF sum score, each genes’ kWithin score was converted to a percentile (i.e., where higher kWithin score indicate greater within-gene-set connectivity). For each subject, normalized kWithin scores were summed across all deletion-spanning genes to generate a total kWithin sum score, capturing total deletion burden weighted by affected genes’ within-gene-set connectivity, across all gene-sets. Exploratory analyses were conducted for deletion LOEUF sum and kWithin sum within each neurodevelopmental gene-set.

**Brainspan Module Gene-Sets**

As described previously(36), the neurodevelopmental gene-sets used to annotate CNVs were derived by applying weighted gene co-expression network analysis to the publicly-available Brainspan transcriptomic data for 17,216 genes in 1,061 tissue samples aged 6 weeks post-conception through 30 years of age, obtained across 16 regions of the brain, including 11 cortical regions, cerebellar cortex and subcortical regions. The resulting 18 mutually-exclusive modules (M1-M18), or gene-sets, were validated and functionally annotated using independent datasets, as recommended(37,38), including identifying significant gene-ontology (GO) term enrichment, brain cell-type specific marker enrichment, enrichment for genes expressed in specific brain regions at specific times in development, transcription factor binding motif enrichment, and known protein-protein interaction enrichment. Hub genes were also identified for each module, representing the most highly connected genes within the module(37). The first principal component (PC) of each module was derived (i.e., eigengene) and its median expression across samples from different developmental stages was plotted to summarize the expression trajectory of each module. To facilitate interpretation for the current analyses, analyses involving these gene-sets are shown with a short functional interpretation label. Module labels were selected to succinctly summarize distinguishing features of enrichment analyses (i.e., based on developmental stage specific expression enrichment patterns, top biological/molecular function GO term enrichment, and cell-type enrichment patterns) and functions of hub genes in each module in normal brain development and function, when known. A summary of the functional characterization of each module is provided in Extended Table 2.

**Quality Control of ABCD Study CNV Calls**

ABCD is a study of over 11,000 demographically diverse youth in the United States (see Fig. S13 for top genetic ancestry principal components). Quality control (QC) and annotation of CNV calls for the ABCD study, based on genotyping signal intensity data for ~500K markers across the genome from the Affymetrix Smokescreen array in hg19 coordinates available in data release 4.0, followed a similar pipeline to the primary SSD-focused cohort. However, given differences in genotyping platform, the pre-computed “affygw6.hmm” model was used to identify CNVs for PennCNV and the “levels-affy.dat” model was used for QuantiSNP. In addition, given lower overall CNV call rates in ABCD compared to the SSD-focused cohort genotyped on the Illumina Global Screening Array and known issues with calling CNVs on sex chromosomes in ABCD from the Affymetrix signal intensity data (https://wiki.abcdstudy.org/release-notes/non-imaging/genetics.html), as well as recommendations to drop samples from plate 461 in data release 4.0 notes, additional QC procedures were implemented. Samples with LRR SD > 0.35 or outliers (>3 SD from the median) in LRR SD, BAF SD, or absolute waviness factor were excluded from analyses. Outliers in total deletion and duplication length were not excluded, as given overall noisier signal intensity data and an overall lower CNV call rate in ABCD, many subjects with large, known pathogenic CNVs that appeared credible on visual inspection of signal intensity plots would have been excluded. QC metrics for LRR SD, absolute waviness factor, total number of deleted or duplicated genes, and total number of top 10% LOEUF genes deleted were additionally assessed by genotyping plate. Plates with mean QC metrics >3 SD from the median on LRR SD or absolute waviness factor or with an excessively high mean number of top 10% LOEUF genes deleted were excluded from analysis (i.e., plates 424, 469, 460, 411, 455, and 461; see Fig. S14-16). Finally, given substantial variance in signal intensity metrics and CNV scores by genotyping plate, plate was included as a random effect covariate in each analysis model.

**MRI Data in ABCD Cohort**

Cortical metrics derived from the T1-weighted MRI data processed with Freesurfer 7.1.1 were obtained from the ABCD Study data release 4.0. Centile generation followed similar steps as in the primary SSD cohort; however, combat correction was not applied to scans prior to scanner/site normalization within the BrainCharts tool given the increased sample sizes across scanning sites in ABCD (<https://brainchart.shinyapps.io/brainchart/>). Individual subject-level centile scores were generated for cortical thickness relative to age- and sex-normative trajectories using the BrainChart tool(22), with all subjects set as controls for scanner/site normalization purposes.

**Sensitivity Analyses for ABCD CNVs Using Alternate CNV Calls**

Analysis of ABCD CNVs was repeated using a recently shared set of calls based on Sha et al., 2025(39). CNV calls were similarly derived using the intersection of PennCNV and QuantiSNP calls; however, quality control procedures differed slightly. Most notably, (39) used more conservative QC metric thresholds, restricting their analyses to CNVs that were a minimum of 20 SNPs and 50 kb in length, and with confidence scores >= 30 for both algorithms. Sample-level QC criteria also differed slightly (see (39) for details). These alternative CNV calls were downloaded from [https://nda.nih.gov/study.html?id=2589](https://urldefense.com/v3/__https:/nda.nih.gov/study.html?id=2589__;!!K-Hz7m0Vt54!j1npZ_rTc7AT5aRhppJYa4CTvEiNUk9l_TPOe9MClWJAI01icWuYm0_mVAtCHJUnMSI7QZer_d1kzLjNxZPZEi-j5keV$).

To maximize comparability with our pipeline, the downloaded calls were re-assessed for >50% overlap with centromeres, telomeres, or segmental duplications for exclusion, as defined for our main pipeline, and rare CNVs retained for analysis were defined as those with <50% overlap of any CNV found with a population maximum frequency ≥1% in gnomAD(28) or within the sample. Our CNV annotations were then applied to the dataset and key CNV variables of interest based on the primary SSD-focused cohort were again tested for replication for association with borderline IQ and MRI-based metrics.

A full list of deletion and duplication calls in ABCD meeting inclusion for analysis based on our original pipeline and for the alternate Sha et al., 2025(39) calls are available on the NIH Data Archive: <https://nda.nih.gov/study.html?id=3105>, with notation for overlapping regions observed across the two CNV calling pipelines and sets of inclusion criteria. Given the more liberal thresholds used for CNV call inclusion in our original pipeline, specified to match the thresholds used for our primary SSD-focused cohort, a large number of smaller CNV calls identified in our original pipeline had no overlapping calls in (39). Based on the 9,305 samples considered to pass QC in both our original pipeline and the pipeline used by (39), the original number of deletions or duplications called per subject was compared to the number of deletions or duplications called per subject, including only CNVs that had overlapping regions called in our original pipeline versus (39). When all original CNV calls were included (i.e., regardless of size thresholds), the correlation in number of calls per subject was *r =* 0.52 for deletions and *r =* 0.63 for duplications. However, when our original CNV calls were subset using a similar set of size and confidence criteria (i.e., minimum confidence score between PennCNV and QuantiSNP of 30, minimum size of 50 kb, minimum SNP size of 20), the correlation in number of calls per subject increased substantially to *r =* 0.80 for deletions and *r =* 0.87 for duplications.

**Supplementary Results**

**CNV Rates Comparing SSD Cases Versus SSD-Relatives Versus Controls**

Logistic mixed models using Wald tests for significance to compare rates of rare deletions and duplications and presence of known risk CNVs between SSD, SSD-relative, and control groups showed similar rates of CNVs in SSD-relatives and controls overall, and similar patterns of difference for both groups compared to SSD cases.

Specifically, the rate of having any rare CNV did not differ significantly between groups (Wald test: χ²=1.30, p=0.52) nor did the rate of having any rare deletions (Wald test: χ²=5.28, p=0.07). However, the presence of rare deletions spanning genes did differ significantly between groups (Wald test: χ²=9.93, p=0.007). Follow-up, pairwise logistic mixed models indicated that SSD cases were more likely to have a rare genic deletion than both controls (OR=1.43, 95%CI[1.08,1.88], *p*=.011) and SSD-relatives (OR=1.67, 95%CI[1.14,2.46], *p*=.009). Conversely, SSD-relatives were not more likely to have rare genic deletions than controls (OR=0.87, 95%CI[0.58,1.29], *p*=.49). Likelihood of having any rare duplication (Wald test: χ²=0.58, p=0.75) or any rare duplication spanning genes (Wald test: χ²=0.83, p=0.66) did not differ between groups.

Due to the low rates of SSD-risk CNVs in SSD-relatives and controls, a model examining differences in SSD-risk CNV rates between the three groups did not converge; however, the three-group comparison for NDD-risk CNVs showed a significant overall group effect (Wald test: χ²=8.20, p=0.017). Pairwise comparisons indicated significantly higher rates of broad NDD-risk CNVs in SSD cases compared to both controls (OR=3.34, 95%CI[1.23,9.10], *p*=.018) and SSD-relatives (OR=8.79, 95%CI[1.12,68.79], *p*=.038), with no difference between SSD-relatives versus controls (OR=0.40, 95%CI[0.05,3.55], *p*=.41).

**CNV Scores versus Child-onset psychosis and Borderline IQ in Narrow SSD Subjects in the SSD-Focused Cohort**

Sensitivity analyses repeating analyses restricted to a narrow SSD definition including schizophrenia and schizophreniform diagnoses only were conducted to ensure that differences between diagnostic groups were not driving association patterns. These sensitivity analyses showed results that were highly similar to the larger sample. Subjects with schizophreniform diagnoses were included in this sensitivity analysis as schizophreniform disorder is a provisional diagnosis given until individuals have had psychotic and related symptoms consistent with schizophrenia for more than 6 months. Given the nature of our recruitment streams which heavily oversampled from recent- and early-onset psychosis clinics, the large majority of individuals initially given schizophreniform diagnoses, if given a SCID at a follow-up time point, would likely meet criteria for schizophrenia. Indeed, all subjects with schizophreniform diagnoses at initial assessment who were re-assessed with a structured clinical interview more than 6 months post initial assessment had converted to schizophrenia diagnoses by follow-up assessment (*n* = 17). In line with this, while ANCOVA comparing the nine SSD groups to one-another for IQ estimate and age of psychosis onset revealed significant overall group differences for IQ, *F*(8,587) = 2.17, *p* =.029, and age of psychosis onset, *F*(8,596) = 2.84, *p* = .004, follow-up pair-wise Tukey comparisons revealed significant group differences only for psychotic disorder not otherwise specified versus schizophrenia for IQ, *p* = .045, and for psychotic disorder not otherwise specified versus schizophrenia and schizophreniform for age of psychosis onset, *p* = .002 and *p* = .004, respectively. The mean IQ estimate and age of psychosis onset in schizophrenia and schizophreniform subjects in the study were highly similar (see Table S2).

Twenty-four of the 434 narrow SSD subjects with age of onset information had psychosis-onset in childhood (5.5%). Of these subjects, only 1 had a SSD- or broader NDD-associated CNV (4.2%). There were no significant associations between known risk CNVs, total burden of deleted genes, total LOEUF score, total kWithin score, nor deletions in any specific neurodevelopmental gene-set and child-onset psychosis in narrow SSD subjects (Figure S8A,C; Extended Table 7).

Eighty-six of the 424 narrow SSD subjects with cognitive data had borderline intellectual functioning (20.3%). Among all narrow SSD subjects, presence of a NDD CNV was associated with borderline IQ (OR = 5.97, 95%[1.45,24.49], p = .013, FDR q = .031; Fig. S6B). Presence of a SSD risk CNV showed a similar effect size, although with reduced power, this was not significant (OR = 5.90, 95%[0.96,36.18], p = .055, FDR q = .098). Deletions in the M1 fetal gene regulatory (OR = 2.83, 95%CI[1.49,5.37], p = .001, FDR q = .023), M6 protein lifecycle (OR = 10.74, 95%CI[1.72,66.99], p = .011, FDR q = .031), and M11 glia and immune signaling (OR = 1.98, 95%CI[1.25,3.15], p = .004, FDR q = .030) gene-sets were associated with borderline IQ, as was total number of deleted genes (OR = 1.12, 95%CI[1.03,1.22], p = .011, FDR q = .031), total deletion LOEUF sum (OR = 1.29, 95%CI[1.05,1.58], p = .013, FDR q = .031), and total deletion kWithin sum (OR = 1.35, 95%CI[1.09,1.68], p = .007, FDR q = .031; Fig. S8D). Deletions in the M15 postnatal synaptic signaling and plasticity gene-set were nominally associated with borderline IQ (OR = 2.12, 95%CI[1.00,4.48], p = .049, FDR q = .098).

Excluding 10 narrow SSD subjects with known risk CNVs revealed a nominal association with borderline IQ for deletions in the M1 fetal gene regulatory gene-set only (OR = 2.81, 95%CI[1.26,6.28], p = .012, FDR q = .139; Figure S8E).

**CNVs of Uncertain Significance versus Child-onset psychosis and Borderline IQ in the SSD-Focused Cohort**

Exploratory analysis of rare deletions or duplications >500 kb in length, excluding subjects with known pathogenic CNVs (i.e., a common definition for potential pathogenic variants of “uncertain significance”; *n* SSD subjects with deletions *=* 25, *n* SSD subjects with duplications = 111), showed no significant associations with borderline IQ (OR = 1.63, 95%CI[0.62,4.28], p = .32 for deletions; OR = 0.74, 95%CI[0.40,1.36], p = .33 or duplications; OR = 0.93, 95%CI[0.54,1.59], p = .79 for deletion or duplication) or child-onset psychosis (OR = 1.36, 95%CI[0.29,6.42], p = .70 for deletions; OR = 1.02, 95%CI[0.42,2.48], p = .97 for duplications, OR = 1.14, 95%CI[0.50,2.58], p = .76 for deletion or duplication).

No deletions spanned genes significantly associated with SSD in the largest extant study of SSD(40), precluding examination of associations between deletions affecting these genes and phenotypic variability in SSD.

**Bonferroni Corrected SSD-Risk CNVs versus Child-Onset Psychosis, and Borderline IQ in the SSD-Focused Cohort**

Restricting analyses of SSD-associated risk CNVs to only loci associated with SSD following Bonferroni correction in Marshall et al., 2017(23) (i.e., excluding 7p36.3 deletions or duplications, 8q22.2 deletions, 9p24.3 deletions or duplications, 15q11.2 deletions, and Xq28 distal duplications) resulted in the exclusion of 4 SSD subjects, who each had 15q11.2 deletions, and 2 control subjects, one with a 15q11.2 deletion and 1 with a 9p24.3 duplication, from the set of subjects identified as having a SSD-risk CNV. Of note, the subjects with 15q11.2 deletions were still included in the set of subjects identified as having a broader NDD-risk CNV, as 15q11.2 deletions were identified as associated with broader developmental disorders in Coe et al., 2014(31).

Thus, using this narrower definition of SSD-risk CNVs, known SSD- and broader NDD-risk CNVs were identified in 5 (0.8%) and 16 (2.6%) SSD subjects, respectively, compared to 0 (0%) and 6 (0.7%) non-cases, respectively. Models comparing rates of SSD-risk CNVs between the SSD, SSD relative, and control groups (i.e., across the 3 groups), or comparing SSD cases to all non-cases did not converge, but the increased rate of broad NDD-risk CNVs in SSD cases versus non-cases was significant (OR=4.49, 95%CI[1.64,12.29], *p*=.003).

As in the main analyses, SSD-risk CNVs were not significantly associated with child-onset psychosis, using the more conservative definition for SSD-risk CNV (OR =3.15, 95%CI[0.31,32.32], *p*=.334), but conservatively-defined SSD-risk CNVs were associated with increased likelihood of borderline IQ among SSD cases (OR=10.82, 95%CI[1.44,81.45], *p*=.021). Analyses relating broader NDD-risk CNVs to severe phenotypes in SSD cases, incorporating the conservative definition for SSD-risk CNVs, were not re-run, as the number of SSD cases affected by broad NDD risk CNVs did not change. The association of broader NDD-risk CNVs in non-cases with borderline IQ, incorporating the conservative definition for SSD-risk CNVs, was not significant, as in the main analysis (OR=2.26, 95%CI[0.20,25.11], *p*=.51), but extending analyses to all SSD cases and non-cases, broader NDD-risk CNVs - defined including only conservatively defined SSD-risk CNVs - was associated with borderline IQ (OR=4.56, 95%CI[1.48,14.10], *p*=.006).

**NDD Risk CNVs Excluding SSD Risk CNVs versus Child-Onset Psychosis and Borderline IQ in the SSD-Focused Cohort**

Restricting the definition of broad NDD risk CNVs to exclude CNVs associated with SSD (hereafter non-SSD NDD-risk CNVs; i.e., CNVs *not* associated with SSD but associated with NDD, ASD, and/or DD), resulted in the exclusion 1 control and 4 SSD subjects carrying 15q11.2 deletions, 1 control subject with a 9p24.3 duplication, 1 SSD subject with a 22q11.2 deletion, 1 SSD subject with a 1q21.1 deletion, 1 SSD subject with a 16p11.2 duplication, and 2 SSD subjects with NRXN1 deletions. With the exception of the 9p24.3 duplication, all of these loci are associated with SSD as well as NDD, ASD, and/or DD. Among the remaining subjects, non-SSD NDD-risk CNVs were observed in 7 of 608 remaining SSD cases (1.1%) and 5 of 812 remaining non-cases (0.6%). Among these remaining subjects, the rate of non-SSD NDD-risk CNVs was elevated but did not differ significantly between cases and non-cases (OR=2.57, 95%CI[0.74,8.98], *p*=.14). There was no association between having a non-SSD NDD-risk CNV in SSD cases and child-onset psychosis (OR=1.97, 95%CI[0.20,19.71], *p*=.56). The rate of borderline IQ in SSD subjects with a non-SSD NDD risk CNV was elevated compared to those without a non-SSD NDD risk CNV, but with reduced power and reduce effect size, this association was not significant (OR=2.48, 95%CI[0.41,14.95], *p*=.32). The association was also elevated but not significant in non-cases alone (OR=2.74, 95%CI[0.22,34.88], *p*=.44), or when extended to all SSD cases and non-cases (OR=2.84, 95%CI[0.64,12.58], *p*=.12) Thus, although the direction of effect was similar to the primary analyses, when CNVs associated with SSD were excluded in the definition of broad NDD-risk CNVs, the effect sizes were weaker and associations with borderline IQ were no longer significant.

**Sensitivity Analyses for Gene-Set Burden Weighted by LOEUF Score or kWithin Connectivity Score versus Child-Onset Psychosis and Borderline IQ in the SSD-Focused Cohort**

The pattern of associations between child-onset psychosis and borderline IQ with each neurodevelopmental gene-set were similar when gene-set burden was weighted by LOEUF score, although associations were slightly attenuated in significance compared to the main analyses. Thus, across all SSD subjects, there were no associations between LOEUF-weighted burden in any neurodevelopmental gene-set or total deletion LOEUF sum and child-onset psychosis (see Extended Table 5; Fig. S6). Across all SSD subjects, total deletion LOEUF sum (OR=1.25, 95%CI[1.05,1.50], *p*=.013, FDR *q*=.065) and LOEUF-score weighted deletion burden in the M1 fetal gene regulatory (OR=3.67, 95%CI[1.46,9.21], *p*=.006, FDR *q*=.065), M6 protein lifecycle (OR=36.43, 95%CI[1.77, 750.42], *p*=.011, FDR *q*=.224), and M11 glia and immune signaling gene-sets (OR=3.30, 95%CI[1.25,8.71], *p*=.016, FDR *q*=.065) were nominally associated with borderline IQ. Excluding SSD subjects with known risk CNVs, only M1 deletions weighted by LOEUF score remained nominally associated with borderline IQ (OR=15.02, 95%CI[1.78,126.63], *p*=.012, FDR *q*=.14). In non-cases, no associations were significant, although the association for M1 was strongest in terms of significance (OR=7.58, 95%CI[0.69,83.67], *p*=.098, FDR *q*=.57). Across all SSD cases and non-cases, excluding subjects with known risk CNVs, only LOEUF-score weighted deletion burden in the M1 fetal gene regulatory gene-set showed an association with increased likelihood of borderline IQ, although this did not survive correction for multiple testing (OR=9.09, 95%CI[1.94,42.69], *p*=.005, FDR *q*=.072).

Alternatively, weighting gene-set burden by within-gene-set connectivity showed associations that were slightly stronger in terms of significance compared to the main analyses. Thus, across all SSD subjects, there were no associations between child-onset psychosis and kWithin-weighted burden in any neurodevelopmental gene-set or total deletion kWithin sum (see Extended Table 6, Fig. S7). Across all SSD subjects, total deletion kWithin sum (OR=1.30, 95%CI[1.07,1.58], *p*=.007, FDR *q*=.046) and kWithin-weighted deletion burden in the M1 fetal gene regulatory gene-set (OR=6.21, 95%CI[2.21,17.42], *p*=.0005, FDR *q*=.007) were significantly associated with borderline IQ (see Extended Table 6). KWithin-weighted M6 protein lifecycle (OR=18.22, 95%CI[1.15, 288.34], *p*=.039, FDR *q*=.128), and M11 glia and immune signaling deletion burden (OR=1.97, 95%CI[1.06,3.65], *p*=.032, FDR *q*=.128) were also nominally associated with borderline IQ. Excluding SSD subjects with known risk CNVs, M1 deletion burden weighted by kWithin score was significantly associated with borderline IQ (OR=5.88, 95%CI[1.85,18.68], *p*=.0026, FDR *q*=.032); kWithin-weighted deletion burden in all other gene-sets were non-significant. In non-cases, kWithin score-weighted M1 deletion burden was nominally associated with borderline IQ (OR=7.38, 95%CI[1.64,33.14], *p*=.009, FDR *q*=.091). Across all SSD cases and non-cases, excluding subjects with known risk CNVs, only kWithin-weighted deletion burden in the M1 fetal gene regulatory gene-set showed a significant association with increased likelihood of borderline IQ (OR=6.20, 95%CI[2.46,15.64], *p*=.0001, FDR *q*=.002).

**Sensitivity Analyses Controlling for Cognitive Assessment Type versus Borderline IQ in the SSD-Focused Cohort**

Analyses of borderline IQ controlling for WAIS or WASI-based cognitive assessments that included both verbal and performance cognitive subtests versus only verbal subtests versus the MCCB showed a similar pattern of results (see Extended Table 8). For these sensitivity analyses, subjects with only performance-based IQ measures (e.g., only the matrix reasoning subtest of the WASI; *n* SSD = 1, *n* SSD-relative = 19, *n* control = 2) were excluded from analysis due to issues with model convergence because only a small number of subjects had this type of IQ estimate. Thus, across all SSD subjects, SSD-risk CNVs and broad NDD-risk CNVs were associated with borderline IQ, (OR=7.20, 95%CI[1.59,32.70], *p*=.011, FDR *q*=.032 and OR=4.66, 95%CI[1.49,14.60], *p*=.008, FDR *q*=.032). Total number of deleted genes (OR=1.11, 95%CI[1.03,1.20], *p*=.009, FDR *q*=.031), total LOEUF sum (OR=1.25, 95%CI[1.04,1.50], *p*=.016, FDR *q*=.042), and total kWithin sum (OR=1.30, 95%CI[1.07,1.57], *p*=.009, FDR *q*=.031) were also associated with borderline IQ, as was deletion burden in the M1 fetal gene regulatory (OR=2.22, 95%CI[1.30,3.77], *p*=.003, FDR *q*=.031), M6 protein lifecycle (OR=7.04, 95%CI[1.36,36.28], *p*=.020, FDR *q*=.044), and M11 glia and immune signaling gene-sets (OR=1.82, 95%CI[1.20,2.76], *p*=.005, FDR *q*=.031). Excluding SSD subjects with known risk CNVs, only M1 deletions remained nominally associated with borderline IQ (OR=2.53, 95%CI[1.22,5.24], *p*=.012, FDR *q*=.173). Analyses in non-cases were not re-run controlling for cognitive assessment types, as all remaining non-cases (i.e., aside from those who received only the matrix reasoning subtest of the WASI) were assessed via comprehensive cognitive assessments that included performance and verbal subtests. In analyses across all SSD cases and non-cases, excluding subjects with known risk CNVs, only deletion burden in the M1 fetal gene regulatory gene-set was significantly associated with increased likelihood of borderline IQ (OR=2.53, 95%CI[1.41,4.54], *p*=.002, FDR *q*=.029).

**Sensitivity Analyses Controlling for DNA Tissue Type, Batch, or Deletion Burden Outside Gene-Set of Interest versus Child-Onset Psychosis and Borderline IQ in the SSD-Focused Cohort**

Batch and tissue source were partially conflated with recruitment stream and therefore potentially partially conflated with clinical characteristics of interest. For example, all participants in NAPLS 2 and 3 had blood DNA samples, while the vast majority of subjects from the MEND study of adolescent-onset SSD patients had saliva DNA samples. Similarly, participants from the Child-Onset Schizophrenia Family Study (i.e., where the largest portion of child-onset psychosis patients came from) were genotyped in batch 1, whereas all participants from the NAPLS 2 and 3 and MEND study were genotyped in batch 2.

Nevertheless, to examine if similar associations exist controlling for DNA tissue type, logistic regressions testing associations between primary CNV scores and child-onset psychosis and borderline IQ were re-run excluding 1 subject with a buccal swab DNA sample and 1 with an unknown DNA tissue type. Across all SSD subjects controlling for saliva versus blood sample, there were no significant associations between any CNV scores and likelihood of childhood-onset SSD, *p*s>.05 (see Extended Table 9). Results of exploratory analyses of known risk deletions versus duplications were not significant for broad NDD deletions (OR=3.83, 95%CI[0.87,16.76], *p*=.075) or SSD deletions (OR=4.47, 95%CI[0.78,25.51], *p*=.09), and the model for NDD risk duplications controlling for DNA tissue type did not converge.

Results for borderline IQ controlling for saliva versus blood sample were also highly similar, as summarized in Extended Table 9. Across all SSD subjects, SSD risk CNVs and broad NDD risk CNVs were associated with borderline IQ, (OR=7.09, 95%CI[1.59,31.53], *p*=.010, FDR *q*=.030 and OR=4.61, 95%CI[1.48,14.37], *p*=.008, FDR *q*=.030). Total number of deleted genes (OR=1.11, 95%CI[1.03,1.20], *p*=.007, FDR *q*=.030), total LOEUF sum (OR=1.26, 95%CI[1.05,1.51], *p*=.013, FDR *q*=.033), and total kWithin sum (OR=1.31, 95%CI[1.08,1.59], *p*=.007, FDR *q*=.030) were also associated with borderline IQ, as was deletion burden in the M1 fetal gene regulatory (OR=2.32, 95%CI[1.36,3.94], *p*=.002, FDR *q*=.030), M6 protein lifecycle (OR=6.65, 95%CI[1.40,31.71], *p*=.017, FDR *q*=.039), and M11 glia and immune signaling gene-sets (OR=1.84, 95%CI[1.21,2.80], *p*=.004, FDR *q*=.030). Excluding SSD subjects with known risk CNVs, only M1 deletions remained nominally associated with borderline IQ (OR=2.61, 95%CI[1.26,5.42], *p*=.010, FDR *q*=.14). Across non-cases, broad NDD risk CNVs (OR=7.94, 95%CI[1.02,61.69], *p*=.048, FDR *q*=.32) and M1 fetal gene regulatory deletion burden were nominally associated with borderline IQ (OR=2.62, 95%CI[1.00,6.81], *p*=.049, FDR *q*=.32). In analyses across all SSD cases and non-cases, excluding subjects with known risk CNVs, only deletion burden in the M1 fetal gene regulatory gene-set was significantly associated with increased likelihood of borderline IQ (OR=2.65, 95%CI[1.49,4.72], *p*=.0009, FDR *q*=.015).

Results were also similar when logistic regressions were run controlling for batch. Thus, across all SSD subjects or across SSD subjects excluding those with known risk CNVs, there were no significant associations between any CNV scores and likelihood of childhood-onset SSD, *p*s>.05 (see Extended Table 10). In exploratory analyses of known risk deletions versus duplications, the relationships for broad NDD deletions (OR=3.71, 95%CI[0.83,16.56], *p*=.086) and SSD deletions were not significant (OR=4.64, 95%CI[0.75,28.71], *p*=.10), and the model for NDD risk duplications controlling for batch did not converge.

Results for borderline IQ controlling for batch were also highly similar as the main analyses, as summarized in Extended Table 10 Across all SSD subjects, SSD risk CNVs and broad NDD risk CNVs were associated with borderline IQ, (OR=6.81, 95%CI[1.53,30.21], *p*=.012, FDR *q*=.030 and OR=4.66, 95%CI[1.50,14.51], *p*=.008, FDR *q*=.028). Total number of deleted genes (OR=1.12, 95%CI[1.03,1.21], *p*=.005, FDR *q*=.028), total LOEUF sum (OR=1.26, 95%CI[1.05,1.51], *p*=.012, FDR *q*=.030), and total kWithin sum (OR=1.31, 95%CI[1.08,1.59], *p*=.006, FDR *q*=.028) were also associated with borderline IQ, as was deletion burden in the M1 fetal gene regulatory (OR=2.26, 95%CI[1.33,3.85], *p*=.003, FDR *q*=.028), M6 protein lifecycle (OR=6.62, 95%CI[1.43,30.74], *p*=.016, FDR *q*=.036), and M11 glia and immune signaling gene-sets (OR=1.81, 95%CI[1.20,2.72], *p*=.005, FDR *q*=.028). Excluding SSD subjects with known risk CNVs, only M1 deletions remained nominally associated with borderline IQ (OR=2.50, 95%CI[1.20,5.19], *p*=.014, FDR *q*=.195). In non-cases, only M1 fetal gene regulatory deletion burden was nominally associated with borderline IQ (OR=2.61, 95%CI[1.00,6.81], *p*=.0496, FDR *q*=.501). Across all SSD cases and non-cases, excluding subjects with known risk CNVs, only deletion burden in the M1 fetal gene regulatory gene-set was significantly associated with increased likelihood of borderline IQ (OR=2.50, 95%CI[1.39,4.50], *p*=.002, FDR *q*=.039).

Logistic regressions testing associations between primary CNV scores and child-onset psychosis and borderline IQ were also re-run for each neurodevelopmental gene-set, covarying for number of genes deleted outside of each gene-set being tested. As summarized in Extended Table 11, results were highly similar overall, with the exception that most individual gene-sets were no longer associated with borderline IQ when subjects with known risk CNVs were included in the analysis, though the M11 glia-immune signaling gene-set remained nominally associated OR=1.64, 95%CI[1.01,2.64], *p*=.045, FDR *q*=.40).This is consistent with the properties of SSD- and NDD-risk CNVs commonly affecting many genes, while being strongly associated with borderline IQ, reducing power to find gene-set specific associations when subjects with NDD-risk CNVs are included in the analysis and all affected genes are accounted for. When SSD subjects with known risk CNVs were excluded from the analysis, M1 deletions showed a similar nominal association with borderline IQ as in the primary analysis (OR=2.85, 95%CI[1.22,6.68], *p*=.016, FDR *q*=.17). Analyses in non-cases were also similar with a nominal association between M1 fetal gene regulatory deletions and borderline IQ (OR=2.77, 95%CI[1.03,7.48], *p*=.044, FDR *q*=.40). Across all SSD cases and non-cases, excluding subjects with known risk CNVs, deletion burden in the M1 fetal gene regulatory gene-set remained significantly associated with increased likelihood of borderline IQ, after controlling for deletion burden affecting genes outside the M1 gene-set (OR=2.69, 95%CI[1.42,5.11], *p*=.002, FDR *q*=.034).

**Sensitivity Analyses Controlling for DNA Tissue Type, Batch, or Non-M1 Genes for MRI Metrics in the SSD-Focused Cohort**

Sensitivity analyses between M1 deletion burden and MRI metrics controlling for DNA tissue type, batch, or burden of deletions affecting non-M1 genes revealed similar patterns of associations.

Thus, controlling for DNA tissue type, excluding subjects with known risk CNVs, M1 deletions were associated with significantly increased GMV (*b*=0.16, 95%CI[0.02,0.29], *p*=.027) and CT in SSD cases (*b*=0.15, 95%CI[0.003,0.30], *p*=.0496), but not SA centile (*b*=0.09, 95%CI[-0.06,0.24], *p*=.23). Extending analyses to all cases and non-cases without known risk CNVs showed associations for GMV (*b*=0.09, 95%CI[0.003,0.17], *p*=.043) and CT (*b*=0.14, 95%CI[0.05,0.24], *p*=.003), but not SA centile (*b*=0.01, 95%CI[-0.08,0.10], *p*=.86).

Controlling for batch, excluding subjects with known risk CNVs, M1 deletions were associated with significantly increased GMV among SSD cases (*b*=0.14, 95%CI[0.008,0.28], *p*=.038) while the association with CT in SSD cases was no longer significant (*b*=0.13, 95%CI[-0.01,0.28], *p*=.075). SA centile was also not associated with M1 deletions (*b*=0.09, 95%CI[-0.06,0.23], *p*=.24). Extending analyses to all cases and non-cases without known risk CNVs, the association with GMV was no longer significant (*b*=0.08, 95%CI[-0.004,0.16], *p*=.062); however, the association with CT remained significant (*b*=0.14, 95%CI[0.04,0.23], *p*=.005). SA centile was again not associated with M1 deletions (*b*=0.01, 95%CI[-0.08,0.10], *p*=.88).

Controlling for the number of genes deleted outside M1, excluding subjects with known risk CNVs, M1 deletions remained associated with significantly increased GMV (*b*=0.15, 95%CI[0.006,0.29], *p*=.040) and CT (*b*=0.16, 95%CI[0.006,0.31], *p*=.041), while SA centile was not associated with M1 deletions (*b*=0.07, 95%CI[-0.08,0.23], *p*=.33). Extending analyses to all cases and non-cases without known risk CNVs, the association with GMV (*b*=0.09, 95%CI[0.006,0.18], *p*=.036) and CT were significant (*b*=0.15, 95%CI[0.05,0.25], *p*=.002), while SA centile was not (*b*=0.00, 95%CI[-0.09,0.10], *p*=.93).

**Exploratory Analyses in ABCD Cohort Excluding One 22q11.2 Deletion Subject**

The lack of significant association between M1 deletion gene count and borderline IQ in the ABCD Study cohort when subjects with known NDD-risk CNVs were included in the analysis is likely due to a combination of noisier CNV calls in the ABCD study, noisier NIH toolbox-based IQ estimates, and/or lower heritability of cognitive functioning in childhood compared to adulthood(41). Indeed, while few ABCD Study subjects were identified as having any M1 genes deleted (*n* = 231 or 2.3% of the sample) and the majority of these subjects had only 1 M1 gene deleted (*n* = 185 or 80.1% of subjects with M1 deletions), one of the two subjects in the ABCD cohort with an extreme number of M1 deleted genes due to having a 22q11.2 deletion, was just above the threshold for borderline IQ. Notably, 22q11.2 deletions are well-known to substantially increase risk for schizophrenia and broader NDDs, including intellectual disability. In line with this, the one other subject in the ABCD cohort who had a 22q11.2 deletion and the same number of deleted M1 genes deleted had an IQ estimate in the borderline IQ range. An exploratory analysis excluding the 22q11.2 deletion subject with an IQ estimate just outside the borderline IQ range showed a significant association between M1 deletion gene count and likelihood of borderline IQ when all other subjects with known NDD-risk CNVs were included in the analysis (OR=1.31, 95%CI[1.05,1.65], *p*=.017). Thus, the inclusion of this 22q11.2 subject as a subject without borderline IQ partially explains the lack of association between number of deleted M1 genes and borderline IQ when subjects with known NDD-risk CNVs were included in the analysis.

**Replication Analyses in ABCD Using the Alternate Sha et al., 2025 CNV Calls**

Using the more conservatively-called CNVs from Sha et al., 2025(39), rates of deletions and duplications were lower, and retained CNV calls were larger, as expected. For example, the number of subjects with deletions retained for analysis that spanned genes dropped from 1426 (14.36%) to 533 (5.37%), and the number of subjects with deletions spanning M1 genes dropped from 231 (2.3%) to 102 (1.0%). See Table S6 and Fig. S18 for summary.

Nevertheless, rates of borderline IQ were similarly found to be significantly elevated in carriers of NDD-risk CNVs (OR=2.17, 95%CI[1.31,3.59], *p*=.003, *q*=.020; Fig. S19). They were also nominally elevated for carriers of SSD-risk CNVs (OR=2.27, 95%CI[1.09,4.76], *p*=.030, *q*=.059). Borderline IQ was not significantly associated with M1 gene deletions, including or excluding subjects with NDD-risk CNVs (OR=1.34, 95%CI[0.96,1.85], *p*=.081, *q*=.13; OR=1.22, 95%CI[0.77,1.93], *p*=.391, *q*=.45, respectively). However, M1 deletions weighted by LOF-intolerance were associated with borderline IQ, both including (OR=2.30, 95%CI[1.28,4.14], *p*=.006, *q*=.022) and excluding subjects with NDD-risk CNVs (OR=2.22, 95%CI[1.09,4.53], *p*=.028, *q*=.059), while M1 deletions weighted by kWithin connectivity were not associated with borderline IQ, including (OR=1.54, 95%CI[0.90,2.61], *p*=.113, *q*=.15) or excluding (OR=1.19, 95%CI[0.58,2.45], *p*=.640, *q*=.64) subjects with NDD-risk CNVs. In the 9,374 youth with QC-passing MRI data, excluding subjects with NDD-risk CNVs, M1 deletion burden was not associated with centile scores, *p*s=*ns* (Fig. S20).

Thus, using the alternate CNV calls with more conservative CNV size thresholds from Sha et al., 2025(39) yielded similar results for borderline IQ, with known risk CNVs showing a significant association with borderline IQ and M1 deletions weighted by LOF-intolerance also associated with borderline IQ. However, M1 deletion burden was not associated with centiles in any direction or with deviance in centiles.

**Sensitivity Analyses Controlling for Genotyping Batch, Tissue Type, and Non-M1 Deletion Burden for Borderline IQ and MRI Metrics in the ABCD Study Cohort**

Controlling for DNA tissue type in the ABCD Study Cohort, rates of borderline IQ remained elevated in carriers of NDD-risk CNVs (OR=1.82, 95%CI[1.21,2.75], *p*=.004, *q*=.018) and non-significantly elevated for SSD-risk CNVs (OR=1.56, 95%CI[0.87,2.80], *p*=.14, *q*=.17). Borderline IQ was not significantly associated with M1 gene deletions when subjects with NDD-risk CNVs were included (OR=1.22, 95%CI[0.98,1.51], *p*=.072, *q*=.12), and was nominally associated excluding subjects with NDD-risk CNVs (OR=1.33, 95%CI[1.00,1.76], *p*=.050, *q*=.100). M1 deletions weighted by LOF-intolerance were nominally associated with borderline IQ, including subjects with NDD-risk CNVs (OR=1.51, 95%CI[1.06,2.16], *p*=.023, *q*=.060), and were significantly associated excluding subjects with NDD-risk CNVs (OR=2.07, 95%CI[1.27,3.37], *p*=.004, *q*=.018). M1 deletions weighted by kWithin score were not associated with borderline IQ, including or excluding subjects with NDD-risk CNVs (OR=1.33, 95%CI[0.91,1.94], *p*=.14, *q*=.16, and OR=1.34, 95%CI[0.83,2.15], *p*=.23, *q*=.23, respectively). Associations with MRI metrics were also similar when controlling for DNA tissue type, with significant associations between M1 gene count and deviance in SA centile (*b*=0.02, 95%CI[0.00,0.03], *p*=.041), M1 LOEUF sum and GMV centile (*b*=-0.06, 95%CI[-0.11,0.0], *p*=.038), SA centile (*b*=-0.06, 95%CI[0.11,-0.01], *p*=.028), and deviance in CT centile (*b*=0.04, 95%CI[0.01,0.07], *p*=.005), and M1 kWithin sum and deviance in CT centile (*b*=0.03, 95%CI[0.00,0.05], *p*=.040; Table S7).

Similarly, controlling for genotyping batch, rates of borderline IQ remained elevated in carriers of NDD-risk CNVs (OR=1.83, 95%CI[1.21,2.76], *p*=.004, *q*=.017) and non-significantly elevated for SSD-risk CNVs (OR=1.57, 95%CI[0.87,2.81], *p*=.13, *q*=.16). Borderline IQ was not significantly associated with M1 gene deletions when subjects with NDD-risk CNVs were included (OR=1.22, 95%CI[0.98,1.52], *p*=.070, *q*=.11), and was nominally associated excluding subjects with NDD-risk CNVs (OR=1.33, 95%CI[1.00,1.76], *p*=.049, *q*=.099). M1 deletions weighted by LOF-intolerance were nominally associated with borderline IQ, including subjects with NDD-risk CNVs (OR=1.51, 95%CI[1.06,2.16], *p*=.023, *q*=.060), and were significantly associated excluding subjects with NDD-risk CNVs (OR=2.07, 95%CI[1.27,3.37], *p*=.004, *q*=.017). M1 deletions weighted by kWithin score were not associated with borderline IQ, including or excluding subjects with NDD-risk CNVs (OR=1.33, 95%CI[0.91,1.94], *p*=.14, *q*=.16, and OR=1.34, 95%CI[0.83,2.16], *p*=.23, *q*=.23, respectively). Associations with MRI metrics were also similar, with significant associations between M1 gene count and deviance in SA centile (*b*=0.02, 95%CI[0.00,0.03], *p*=.043), M1 LOEUF sum and GMV centile (*b*=-0.06, 95%CI[-0.11,0.0], *p*=.039), SA centile (*b*=-0.06, 95%CI[0.11,-0.01], *p*=.031), and deviance in CT centile (*b*=0.04, 95%CI[0.01,0.07], *p*=.006), and M1 kWithin sum and deviance in CT centile (*b*=0.03, 95%CI[0.00,0.05], *p*=.042; Table S7).

When non-M1 deletion burden was controlled for, borderline IQ was not associated with M1 gene deletions when subjects with NDD-risk CNVs were included (OR=0.90, 95%CI[0.69,1.18], *p*=.43, *q*=.81) or excluded (OR=1.07, 95%CI[0.78,1.46], *p*=.67, *q*=.81). M1 deletions weighted by LOF-intolerance were also not associated with borderline IQ, including (OR=0.85, 95%CI[0.52,1.39], *p*=.52, *q*=.81) or excluding subjects with NDD-risk CNVs (OR=1.23, 95%CI[0.70,2.16], *p*=.48, *q*=.81), nor were M1 deletions weighted by kWithin score including (OR=0.87, 95%CI[0.55,1.39], *p*=.56, *q*=.81) or excluding subjects with NDD-risk CNVs, OR=0.97, 95%CI[0.58,1.63], *p*=.91, *q*=.91). However, some associations with MRI metrics remained, specifically, between M1 gene count and deviance in SA centile (*b*=0.02, 95%CI[0.00,0.04], *p*=.017), and M1 LOEUF sum and deviance in CT centile (*b*=0.04, 95%CI[0.01,0.07], *p*=.009). There was also a new association between M1 gene count and deviance in GMV centile (*b*=0.02, 95%CI[0.00,0.03], *p*=.033). See Table S7.

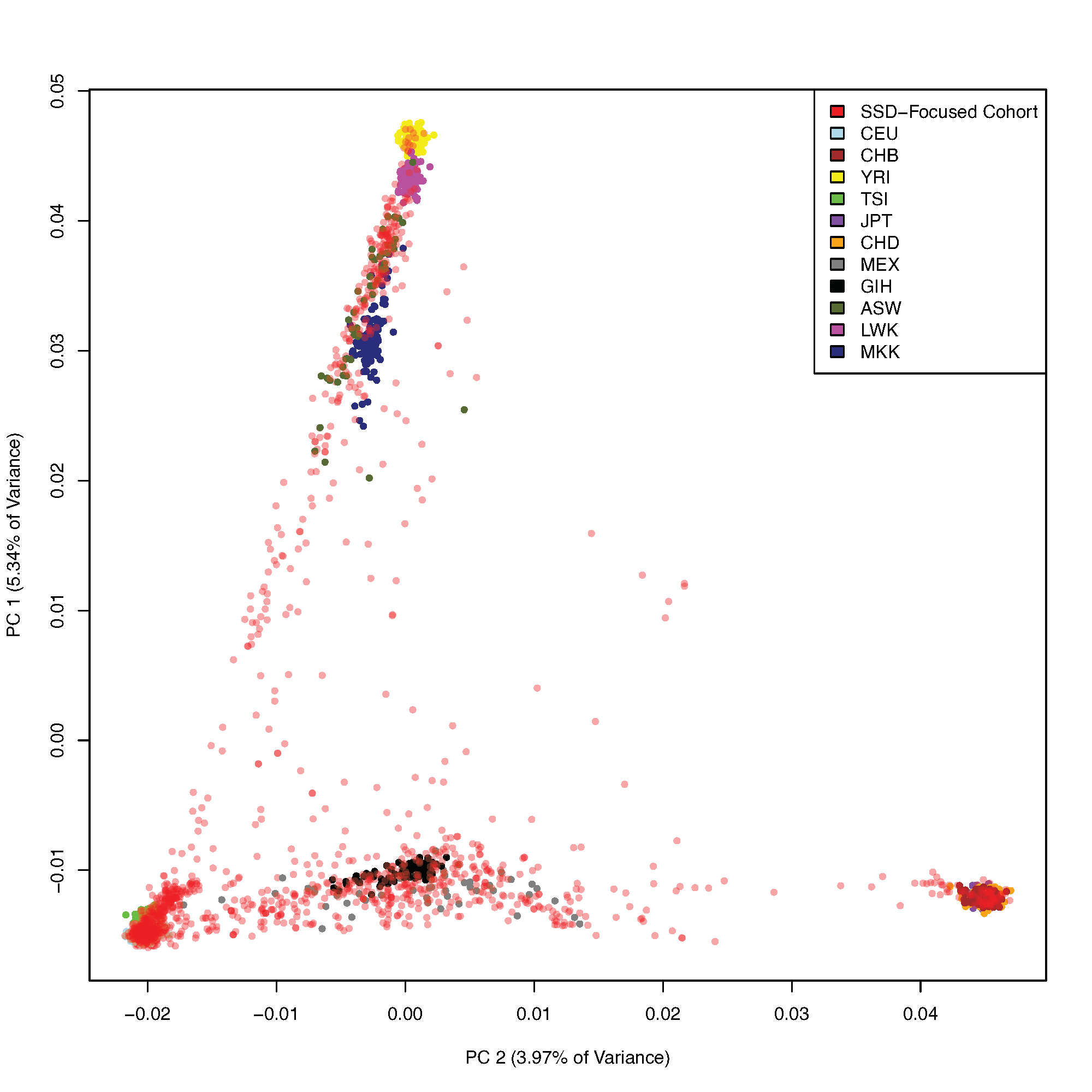

**Supplementary Figure 1.** Top two ancestry principal components (PCs) in the SSD-focused study cohort, overlaid on HapMap3 global populations (CEU = Utah residents with Northern and Western European ancestry, CHB = Han Chinese in Beijing, China, YRI = Yoruba in Ibadan, Nigeria, TSI = Toscans in Italy, JPT = Japanese in Tokyo, Japan, CHD = Chinese in Denver, Colorado, MEX = Mexican ancestry in Los Angeles, California, GIH = Gujarati Indians in Houston, Texas, ASW = African ancestry in Southwest USA, LWK = Luhya in Webuye, Kenya, MKK = Maasai in Kinyawa, Kenya).

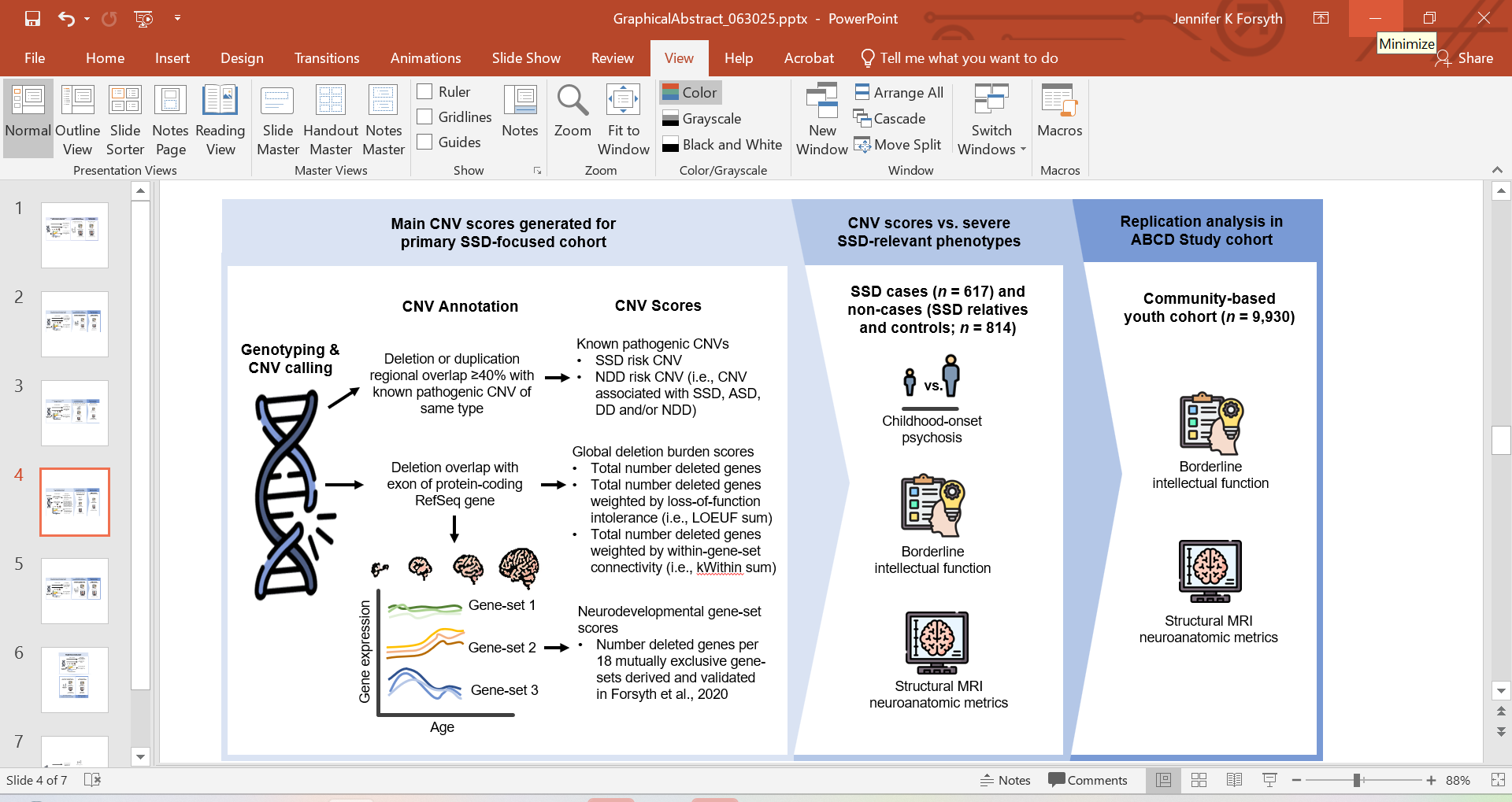

**Supplementary Figure 2.** Schematic of flow of analyses in the primary schizophrenia spectrum disorder (SSD) cohort and replication assessment of key copy number variant (CNV) score associations in the Adolescent Brain Cognitive Development (ABCD) Study cohort. CNV = copy number variant; SSD = schizophrenia spectrum disorder; NDD = neurodevelopmental disorder; ASD = autism spectrum disorder; DD = developmental delay.

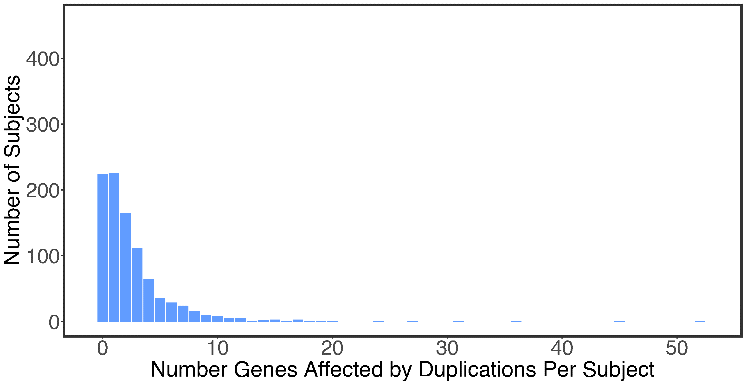

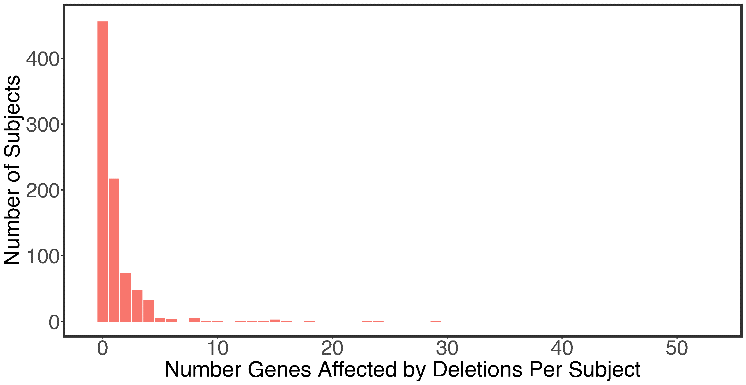

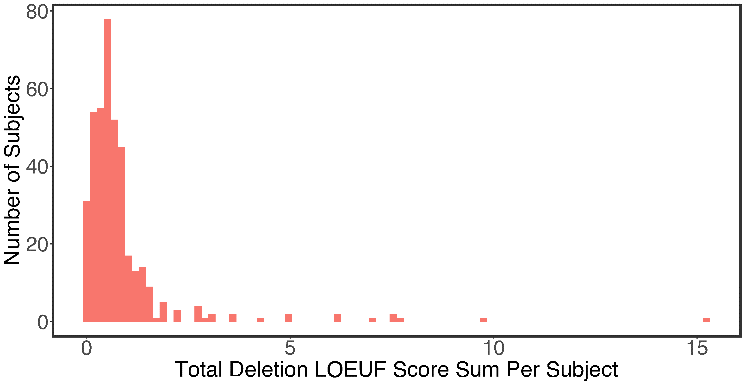

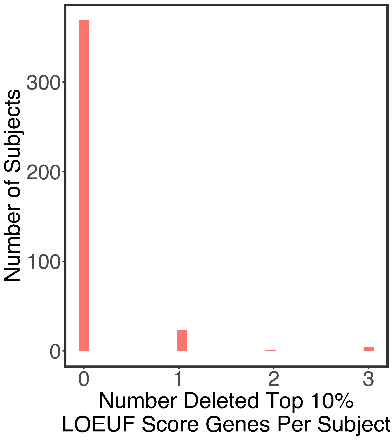

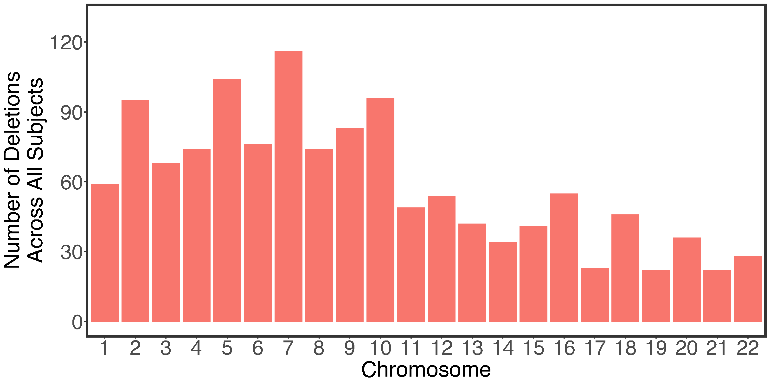

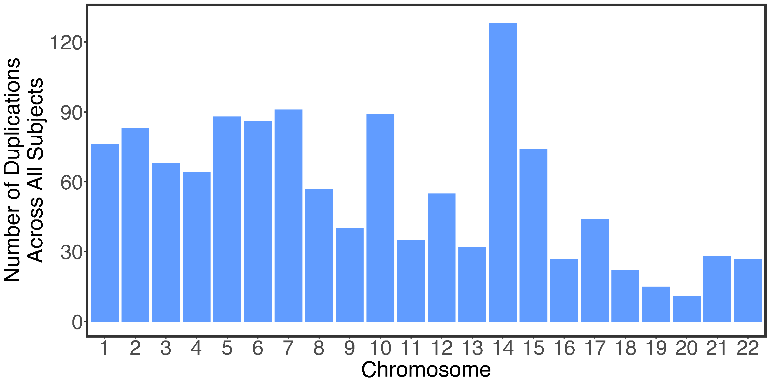

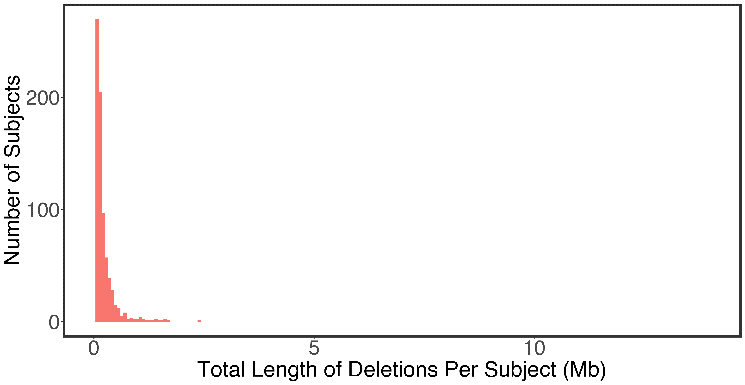

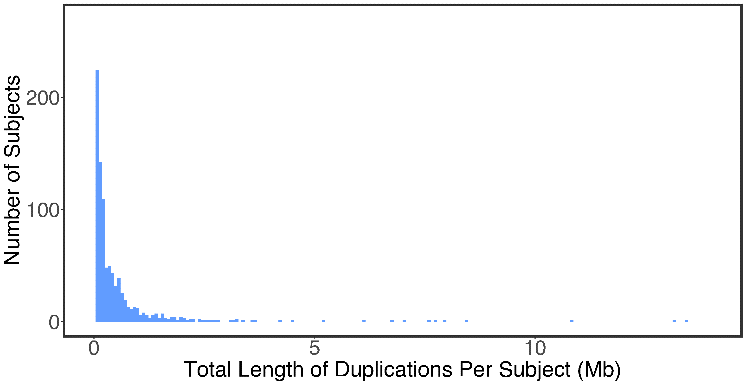

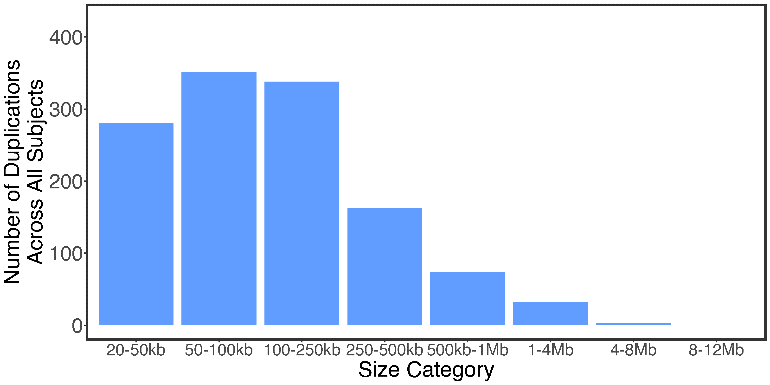

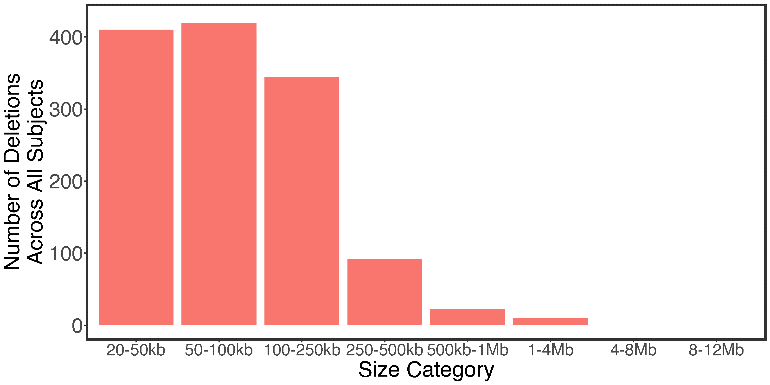

**Supplementary Figure 3.** CNV characteristics in the primary SSD-focused cohort.

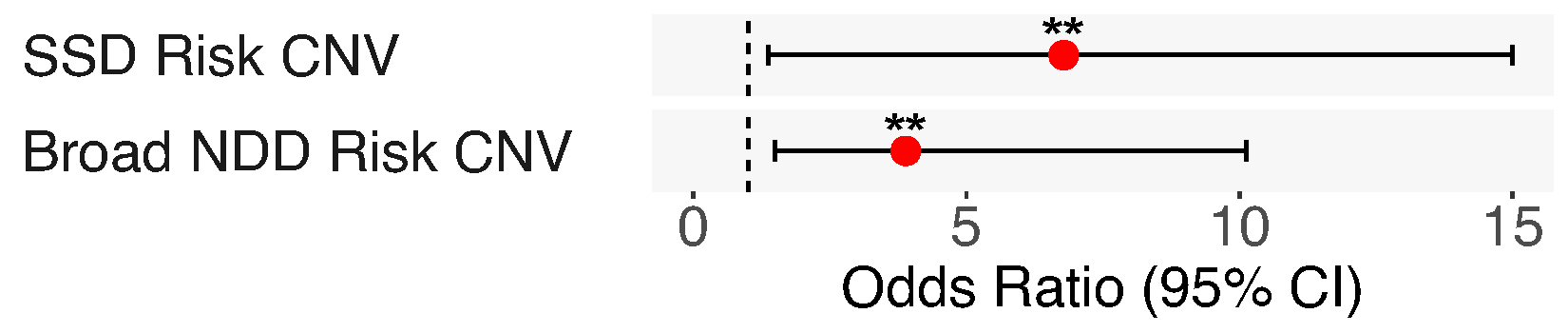

**Supplementary Figure 4.** Odds ratio for SSD case status associated with known risk CNVs. **FDR q < 0.05, corrected for number of independent variables tested. Max OR confidence interval shown = 15.

**A**

**B**

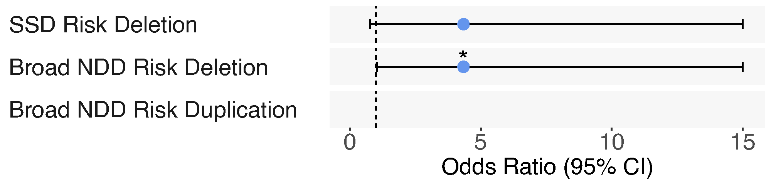

All SSD

Borderline IQ

All SSD

Child-Onset Psychosis

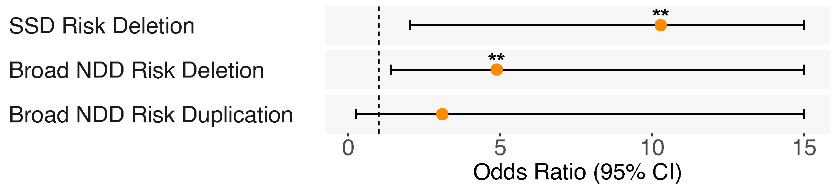

**Supplementary Figure 5.** Odds ratio for (A) child-onset psychosis (*n* with child-onset psychosis = 39, *n* with later-onset psychosis = 568), and (B) borderline IQ (*n* with borderline IQ = 120, *n* without borderline IQ = 478) among SSD cases associated with known SSD-risk deletions, NDD-risk deletions, and NDD-risk duplications. **p* < 0.05, **FDR q < 0.05, corrected for number of independent variables tested per trait. Max OR confidence interval shown = 15.

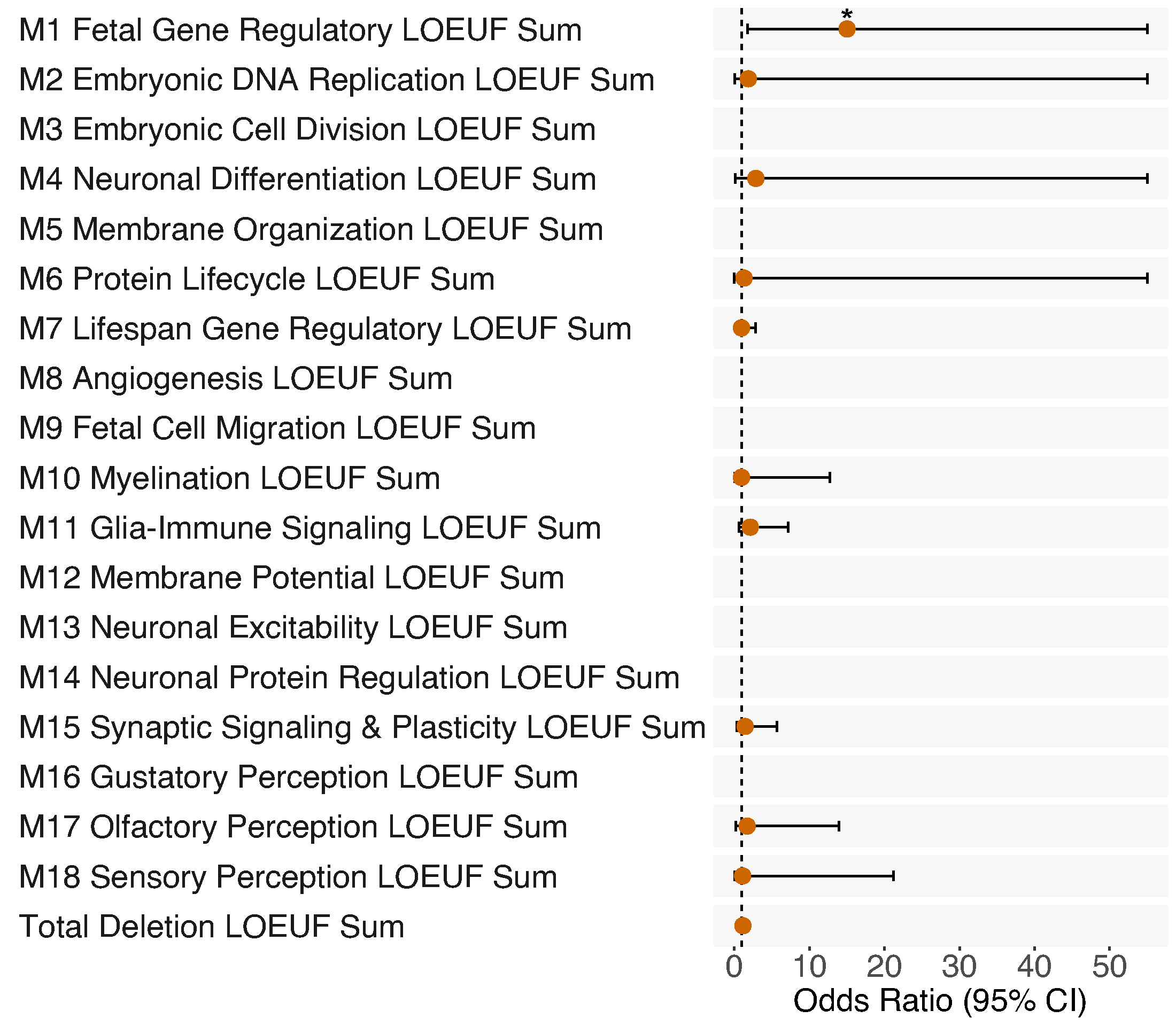

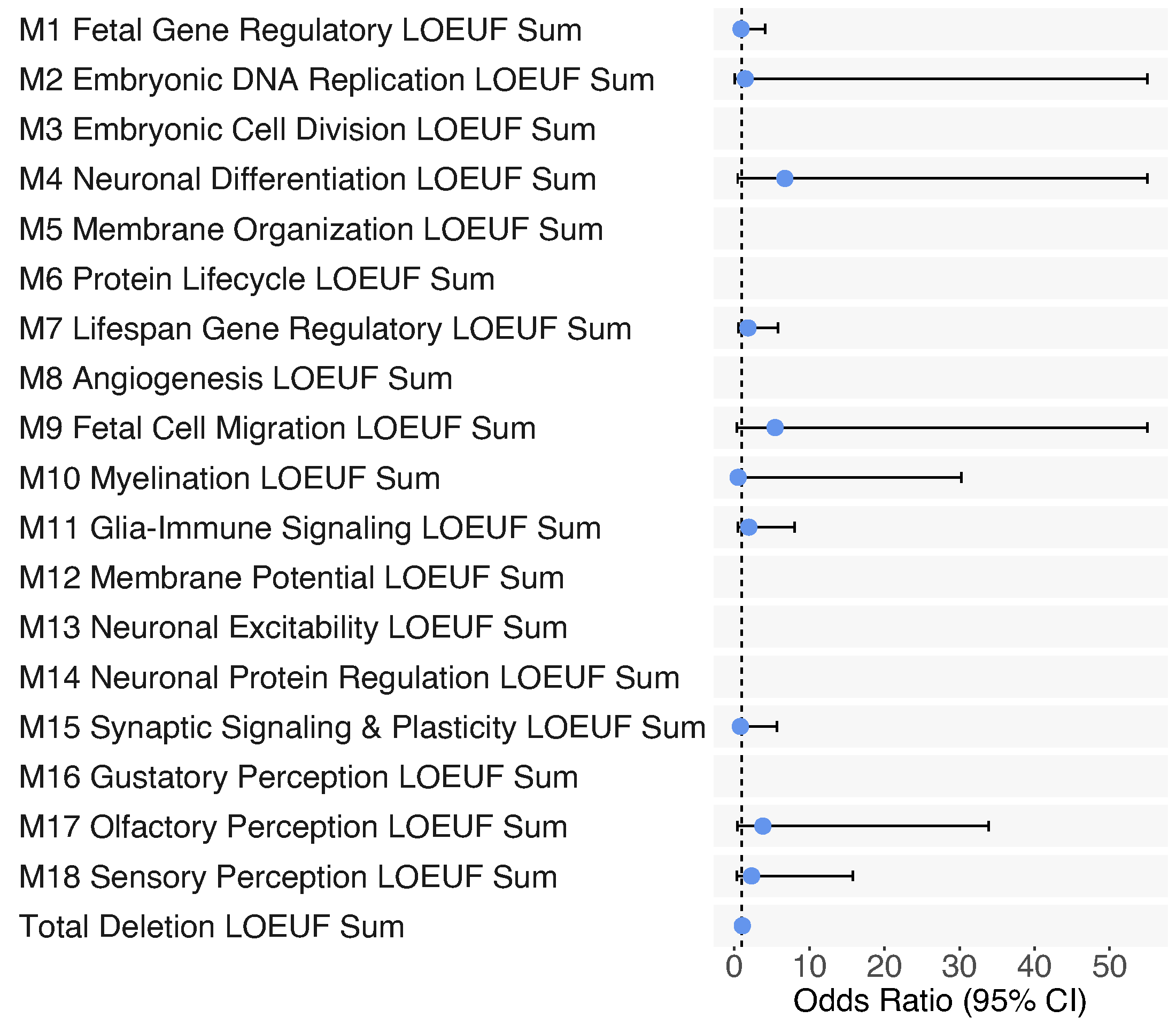

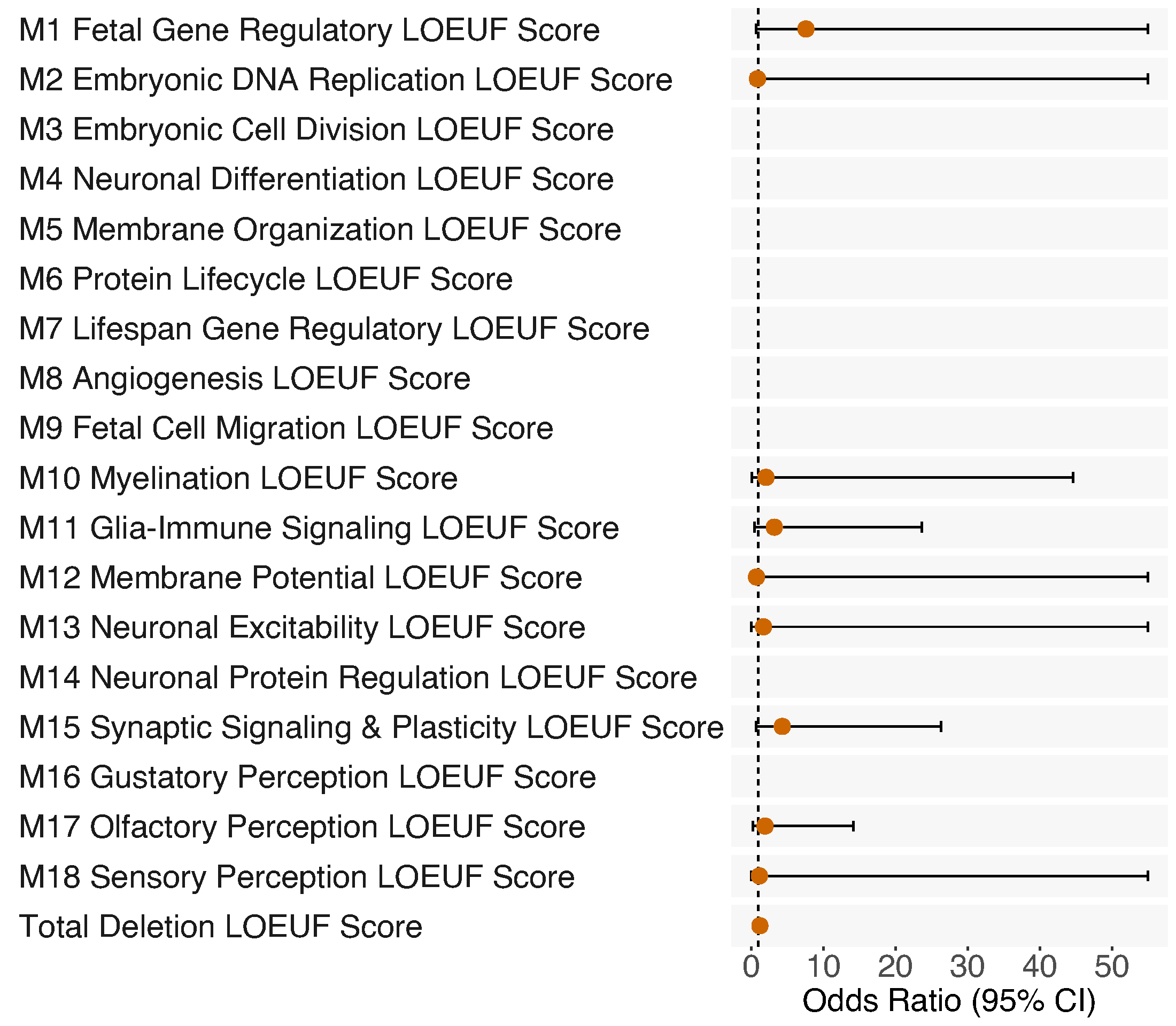

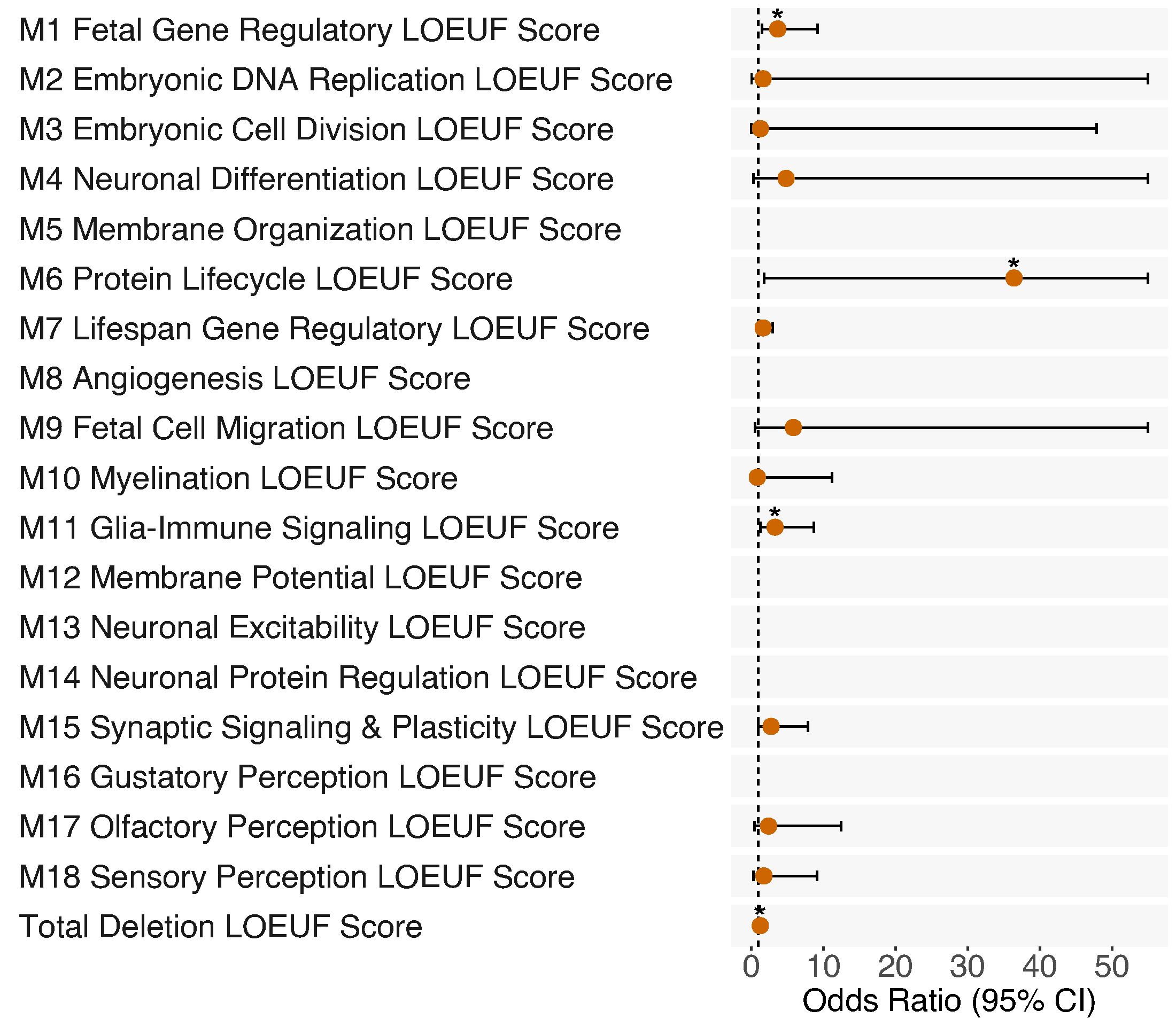

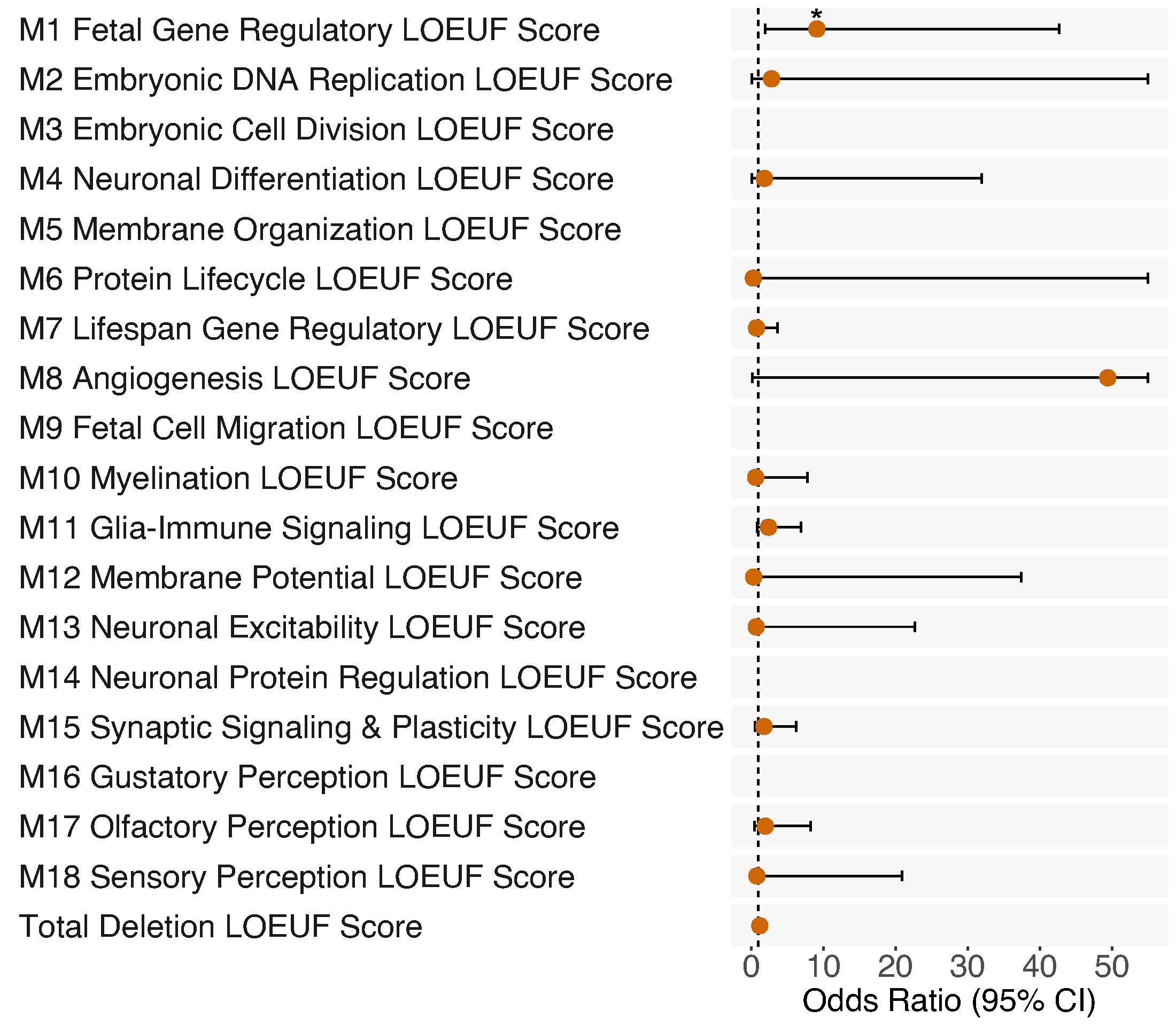

**A**

**B**

**D**

All Non-SSD

Borderline IQ

**C**

**E**

SSD, No Risk CNVs

Borderline IQ

SSD & Non-SSD, No Risk CNVs Borderline IQ

All SSD

Child-Onset Psychosis

All SSD

Borderline IQ

**Supplementary Figure 6.** Odds ratio (OR) associated with sum of number of genes deleted per neurodevelopmental gene-set weighted by LOEUF score versus total deletion LOEUF sum for (A) child-onset psychosis (*n* with child-onset psychosis = 39, *n* with later-onset psychosis = 568) and for borderline IQ across (B) all SSD subjects (*n* with borderline IQ = 120, *n* without borderline IQ = 478), (C) SSD cases excluding those with known risk CNVs (*n* with borderline IQ = 113, *n* without borderline IQ = 469), (D) all non-SSD subjects (*n* with borderline IQ = 57, *n* without borderline IQ = 741), and (E) SSD cases, SSD-relatives, and controls, excluding subjects with known risk CNVs (*n* with borderline IQ = 168, *n* without borderline IQ = 1210). **p* < 0.05. Max OR confidence interval shown = 55; see Extended Table 5 for full statistics.

**A**

**B**

**D**

All Non-SSD

Borderline IQ

SSD & Non-SSD, No Risk CNVs Borderline IQ

**C**

**E**

SSD, No Risk CNVs

Borderline IQ

All SSD

Child-Onset Psychosis

All SSD

Borderline IQ

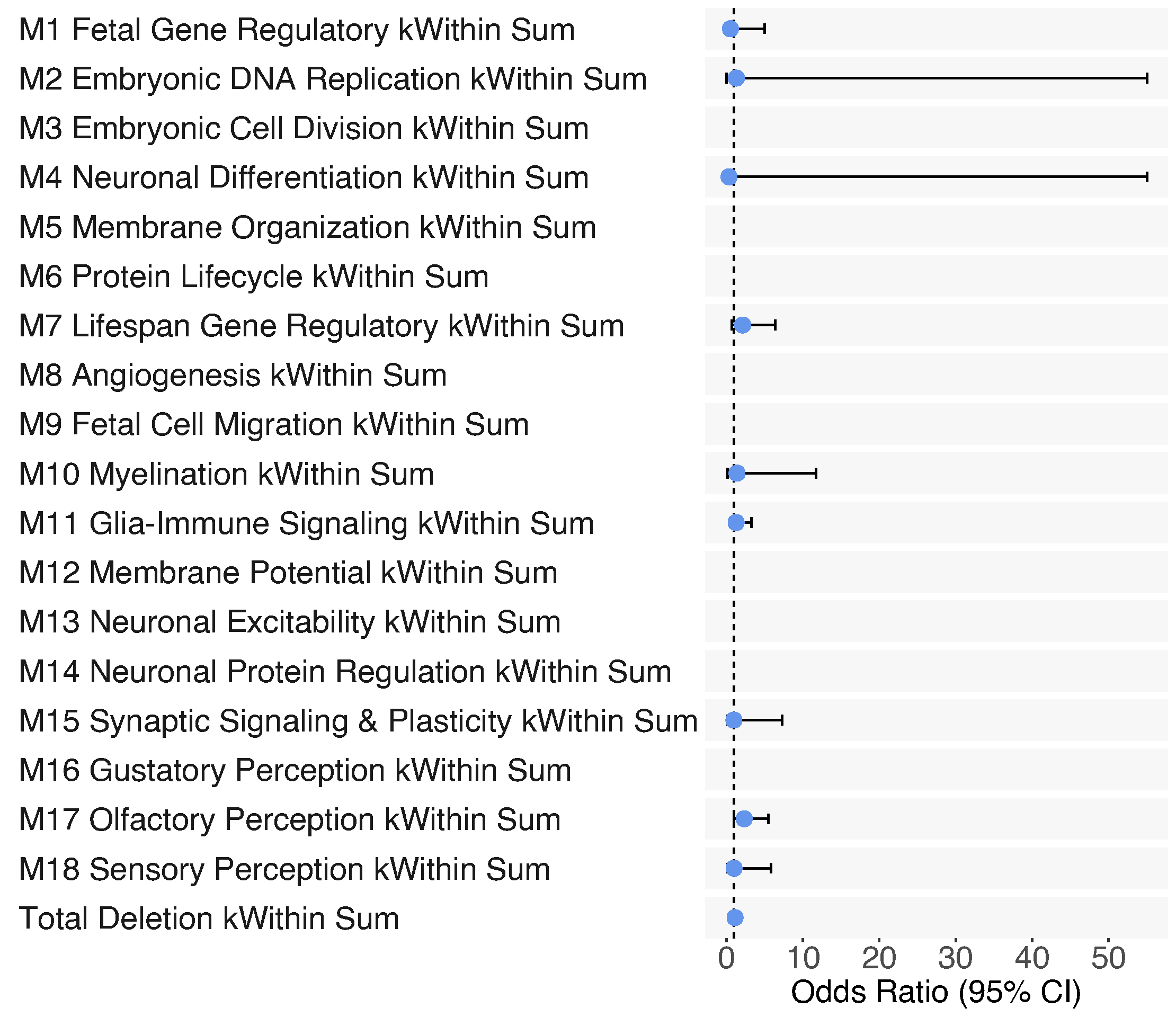

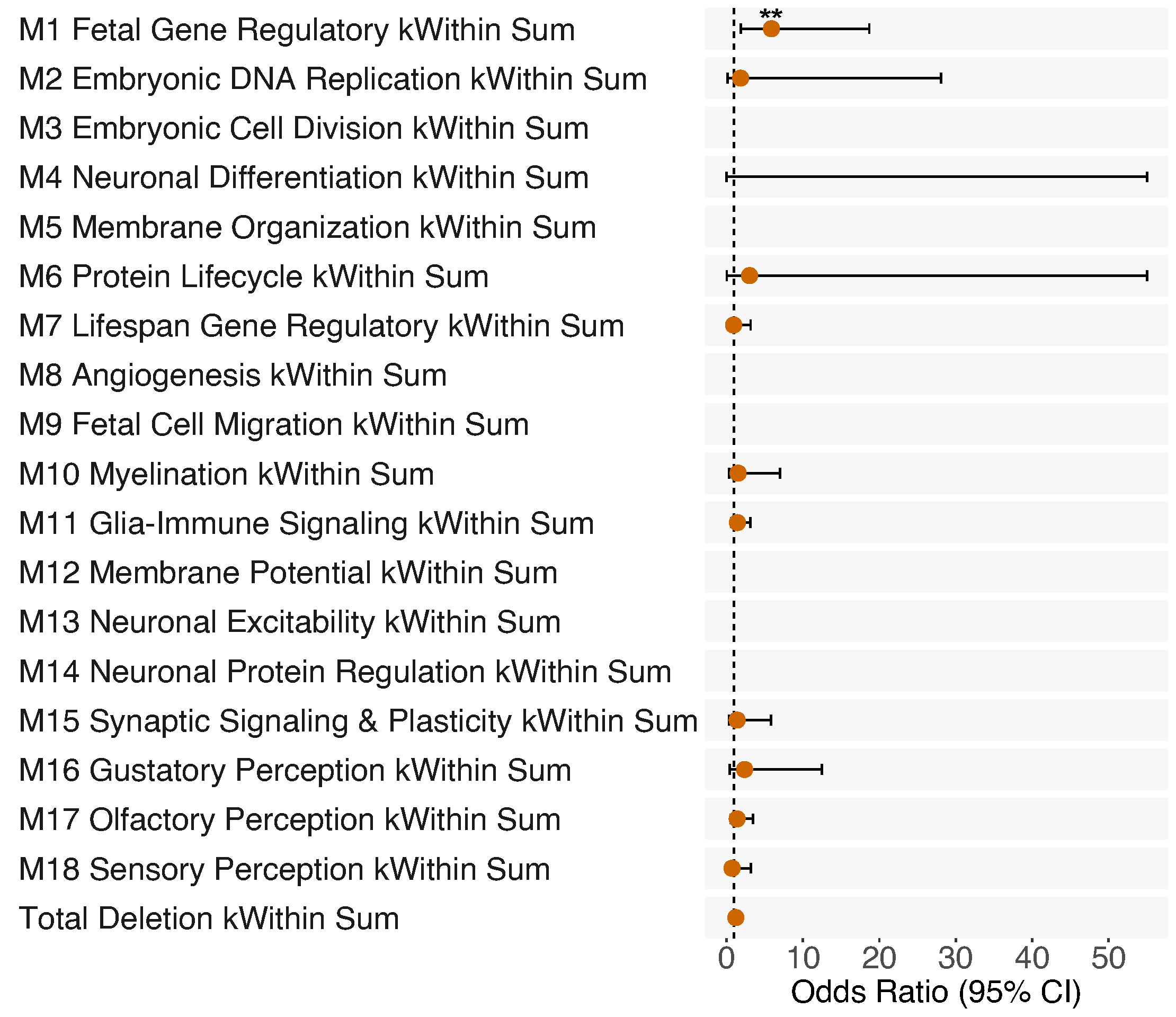

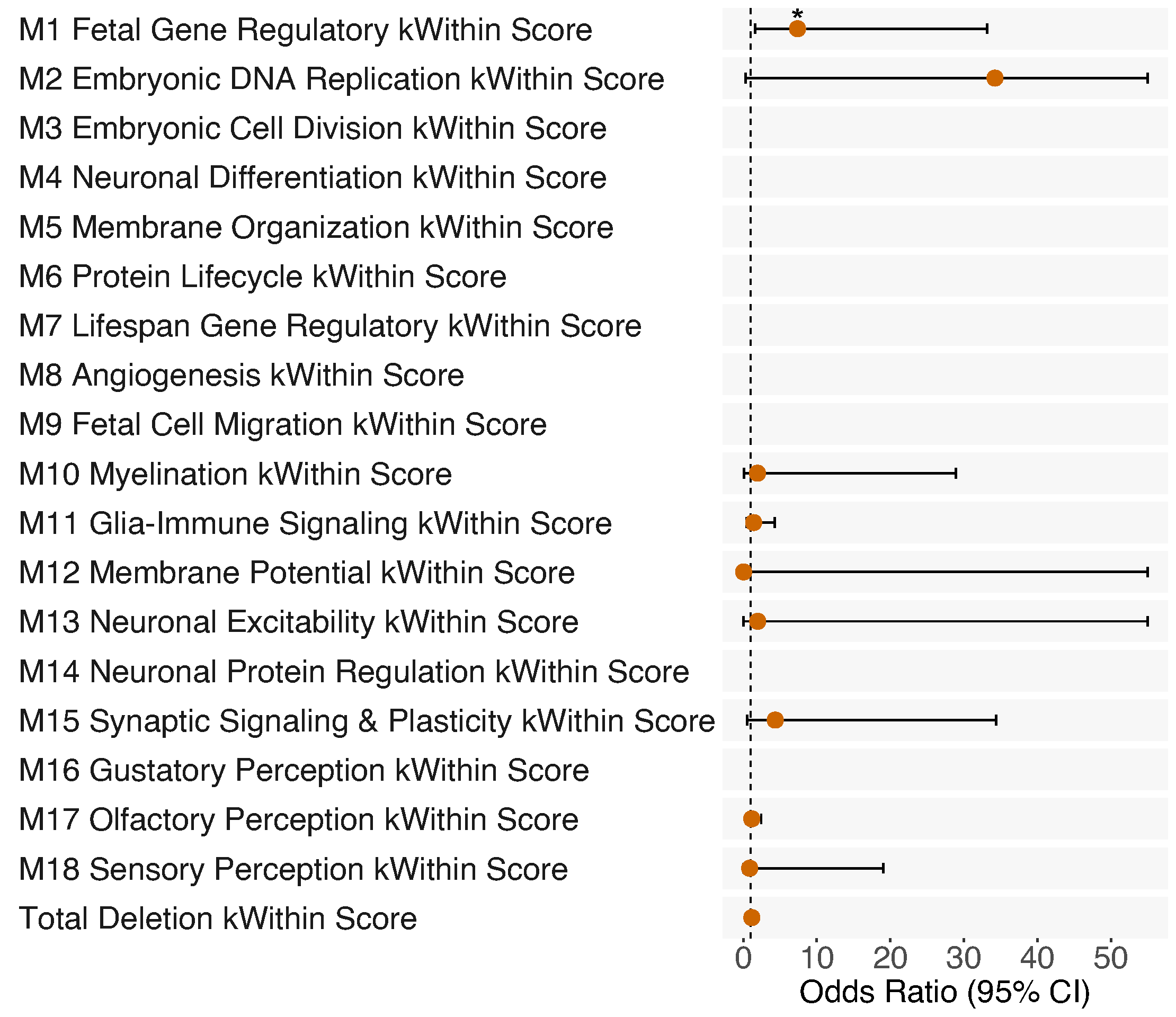

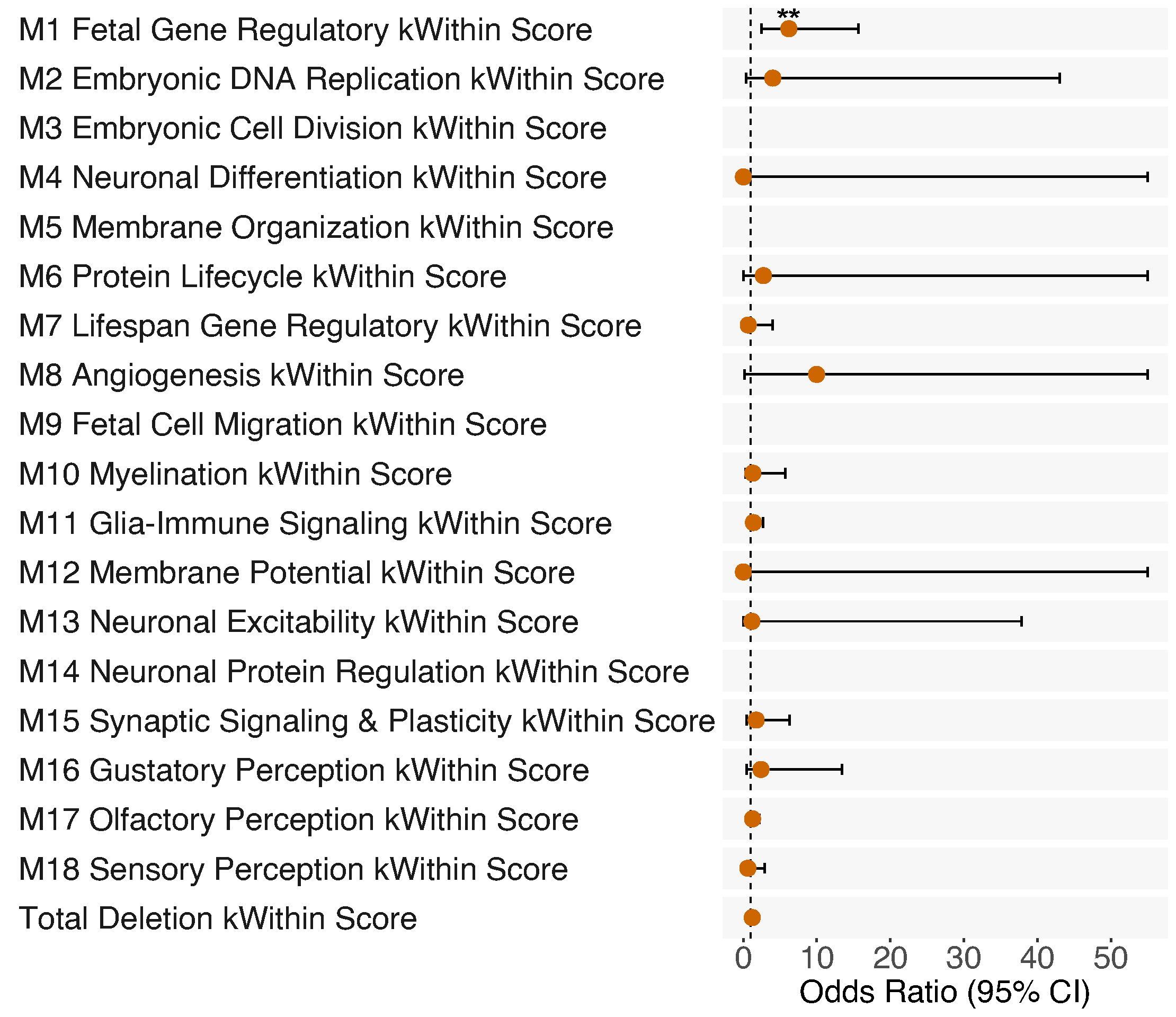

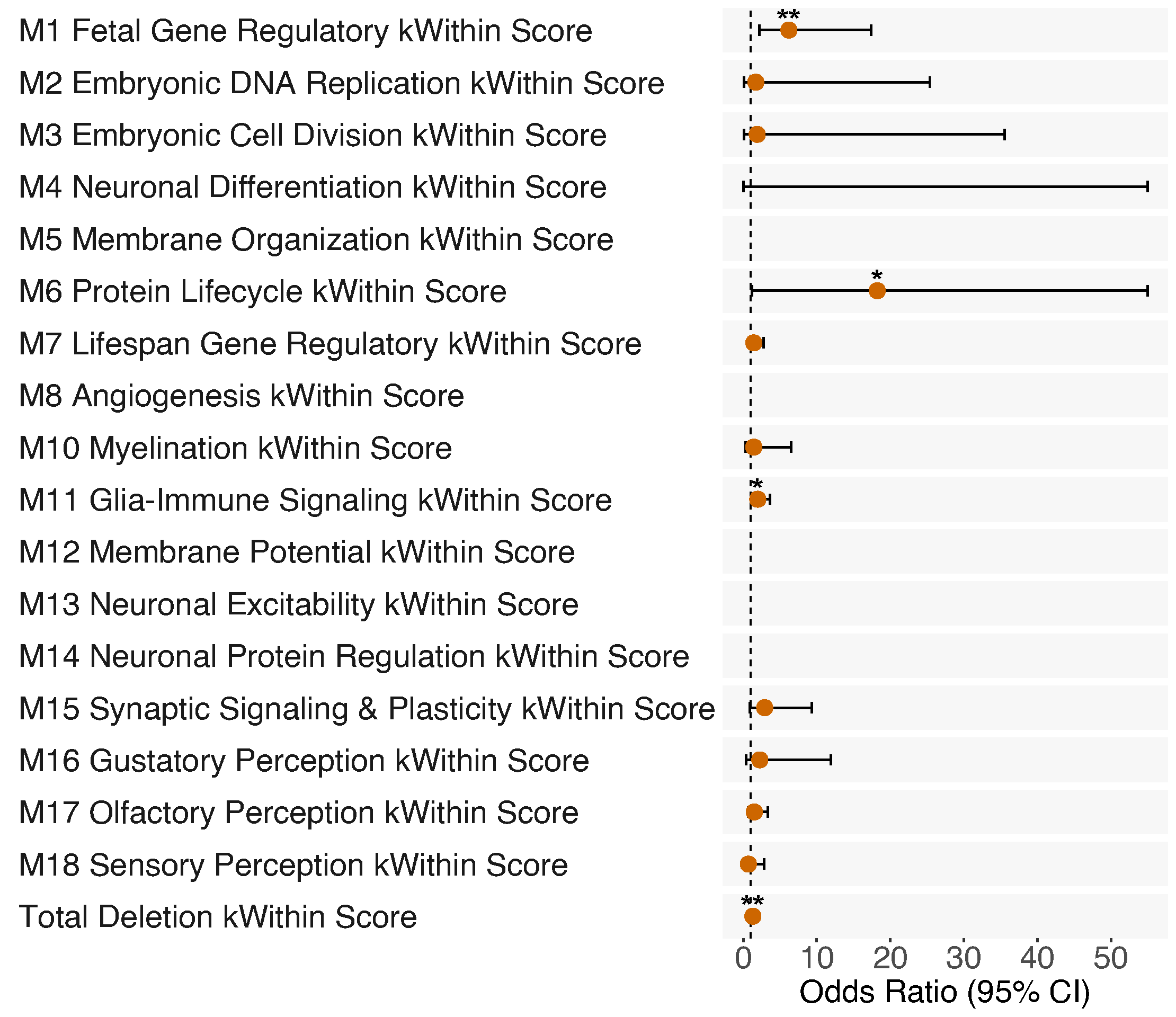

**Supplementary Figure 7.** Odds ratio (OR) associated with sum of number of genes deleted per neurodevelopmental gene-set weighted by kWithin gene-set connectivity score versus total deletion kWithin sum for (A) child-onset psychosis (*n* with child-onset psychosis = 39, *n* with later-onset psychosis = 568) and for borderline IQ across (B) all SSD subjects (*n* with borderline IQ = 120, *n* without borderline IQ = 478), (C) SSD cases excluding those with known risk CNVs (*n* with borderline IQ = 113, *n* without borderline IQ = 469), (D) all non-SSD subjects (*n* with borderline IQ = 57, *n* without borderline IQ = 741), and (E) SSD cases, SSD-relatives, and controls, excluding subjects with known risk CNVs (*n* with borderline IQ = 168, *n* without borderline IQ = 1210). **p* < 0.05. **FDR q < 0.05, corrected for number of independent variables tested per trait. Max OR confidence interval shown = 55; see Extended Table 6 for full statistics.

All Narrow SSD

Child-Onset Psychosis

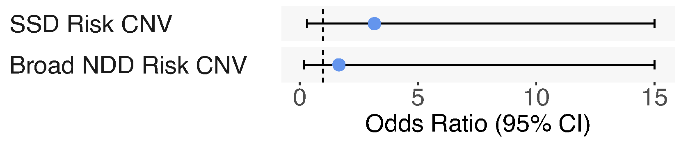

**A**

**B**

All Narrow SSD

Borderline IQ

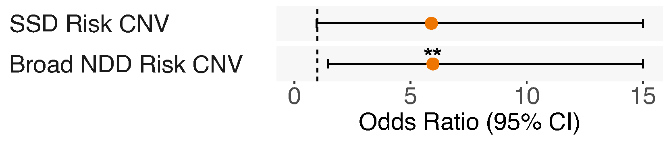

All Narrow SSD

Borderline IQ

All Narrow SSD

Child-Onset Psychosis

**C**

**E**

**D**

Narrow SSD, No Risk CNVs

Borderline IQ

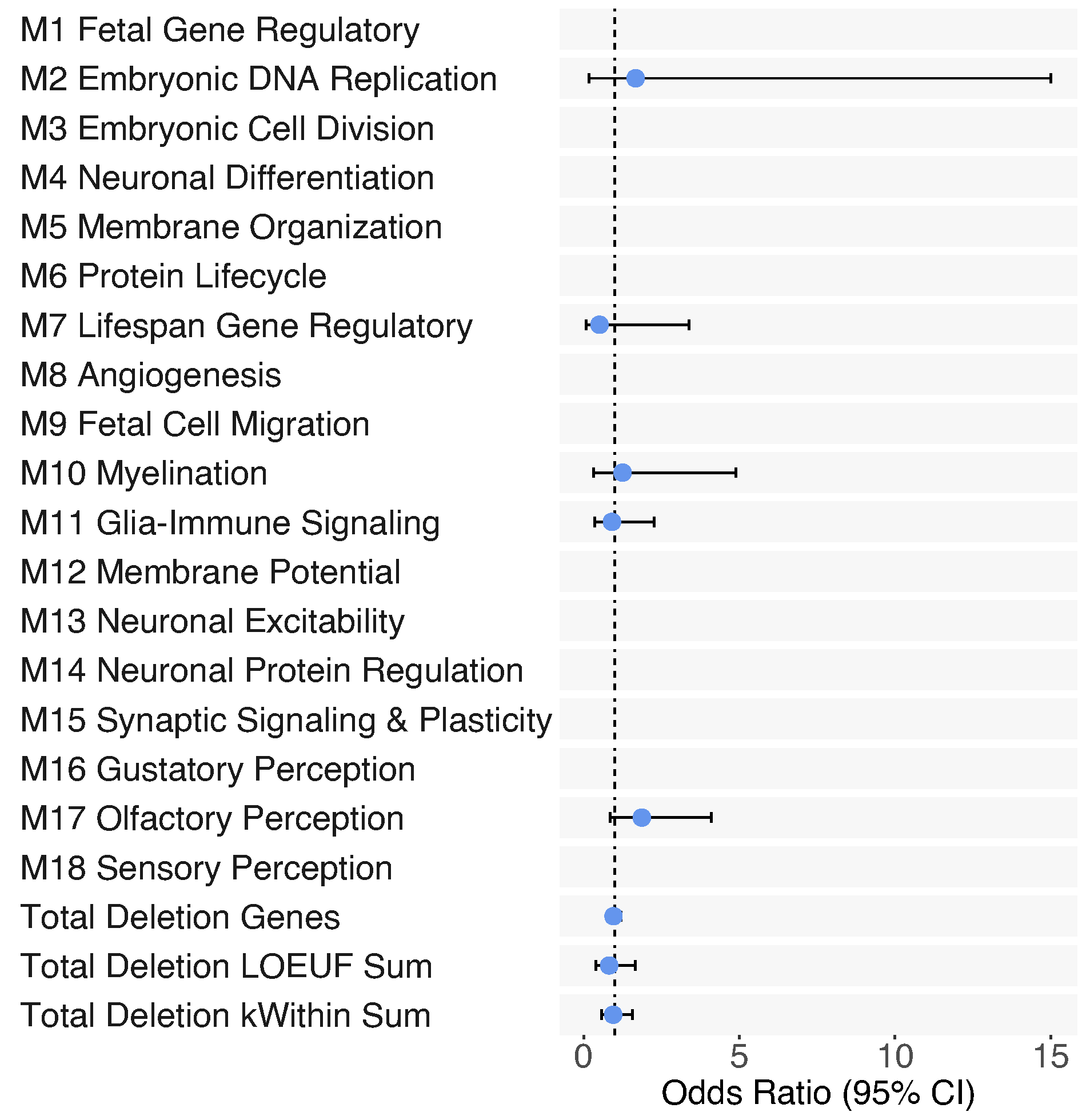

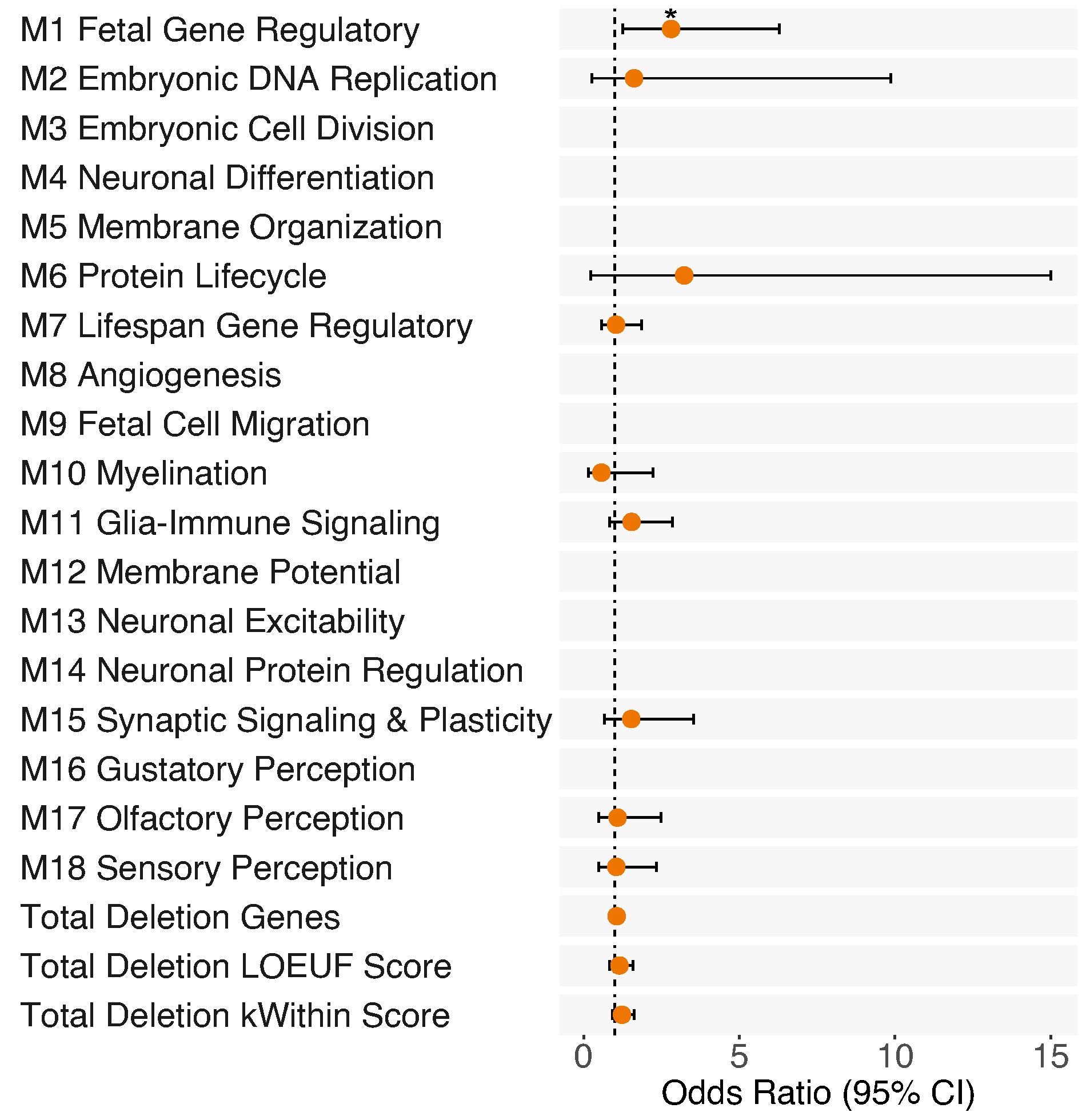

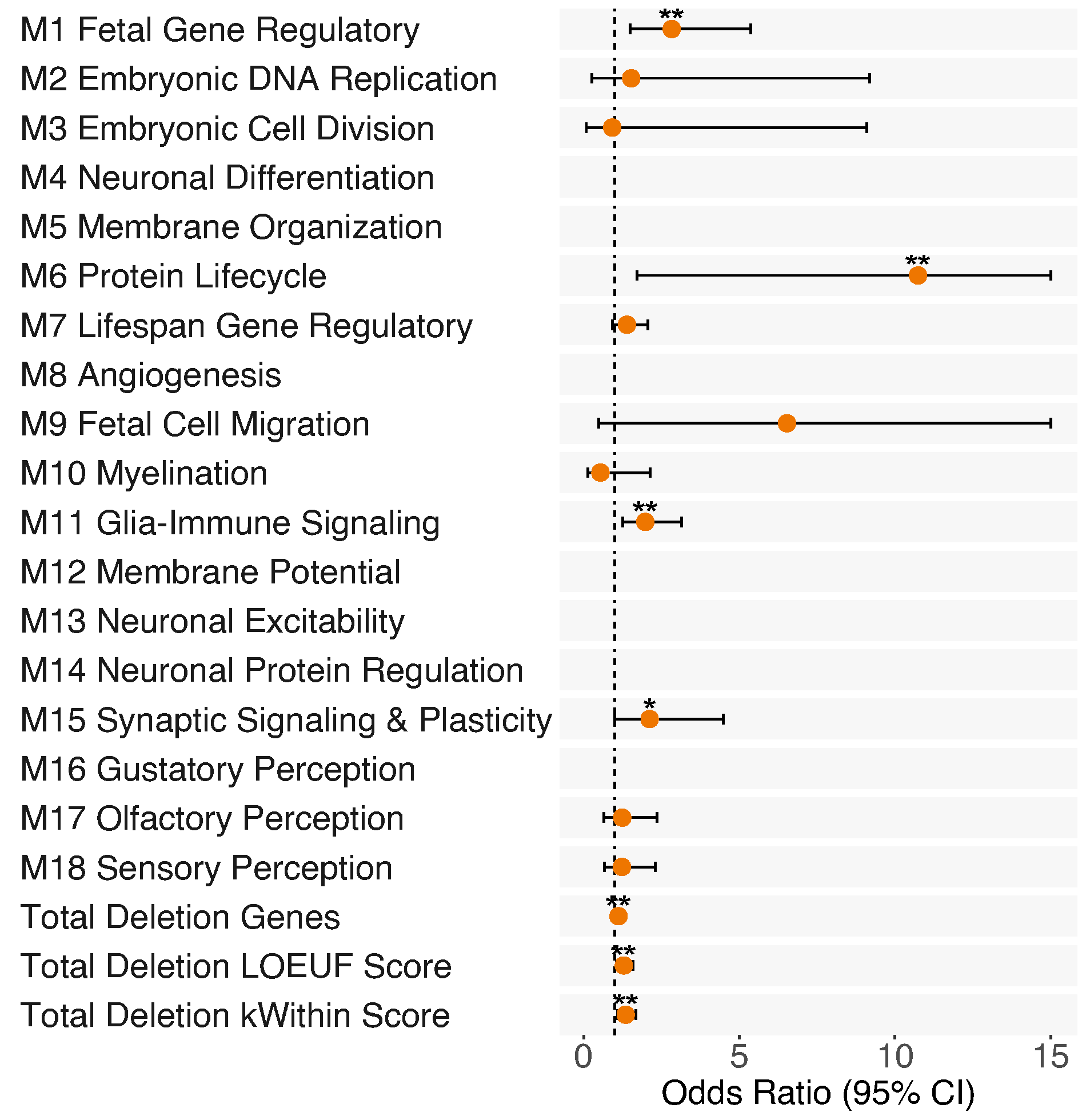

**Supplementary Figure 8.** Odds ratio (OR) associated with presence of known risk CNVs for (A) child-onset psychosis (*n* with child-onset psychosis = 24, *n* with later-onset psychosis = 410) and (B) borderline IQ (*n* with borderline IQ = 86, *n* without borderline IQ = 338) across all narrowly-defined SSD subjects, and for (C) child-onset psychosis and (D) borderline IQ associated with number of genes deleted per neurodevelopmental gene-set and global deletion burden scores. (E) OR associated with number of genes deleted per neurodevelopmental gene-set and global deletion burden scores for borderline intellectual functioning in narrowly-defined SSD subjects after excluding subjects with known risk CNVs (*n* with borderline IQ = 81, *n* without borderline IQ = 333). **p* < 0.05; **FDR q < 0.05, corrected for number of CNV scores tested per analysis group. Max OR confidence interval shown = 15; see Extended Table 6 for full statistics.

**A**

**B**

**D**

All Non-SSD

Borderline IQ

SSD & Non-SSD, No Risk CNVs Borderline IQ

**C**

**E**

SSD, No Risk CNVs

Borderline IQ

All SSD

Child-Onset Psychosis

All SSD

Borderline IQ

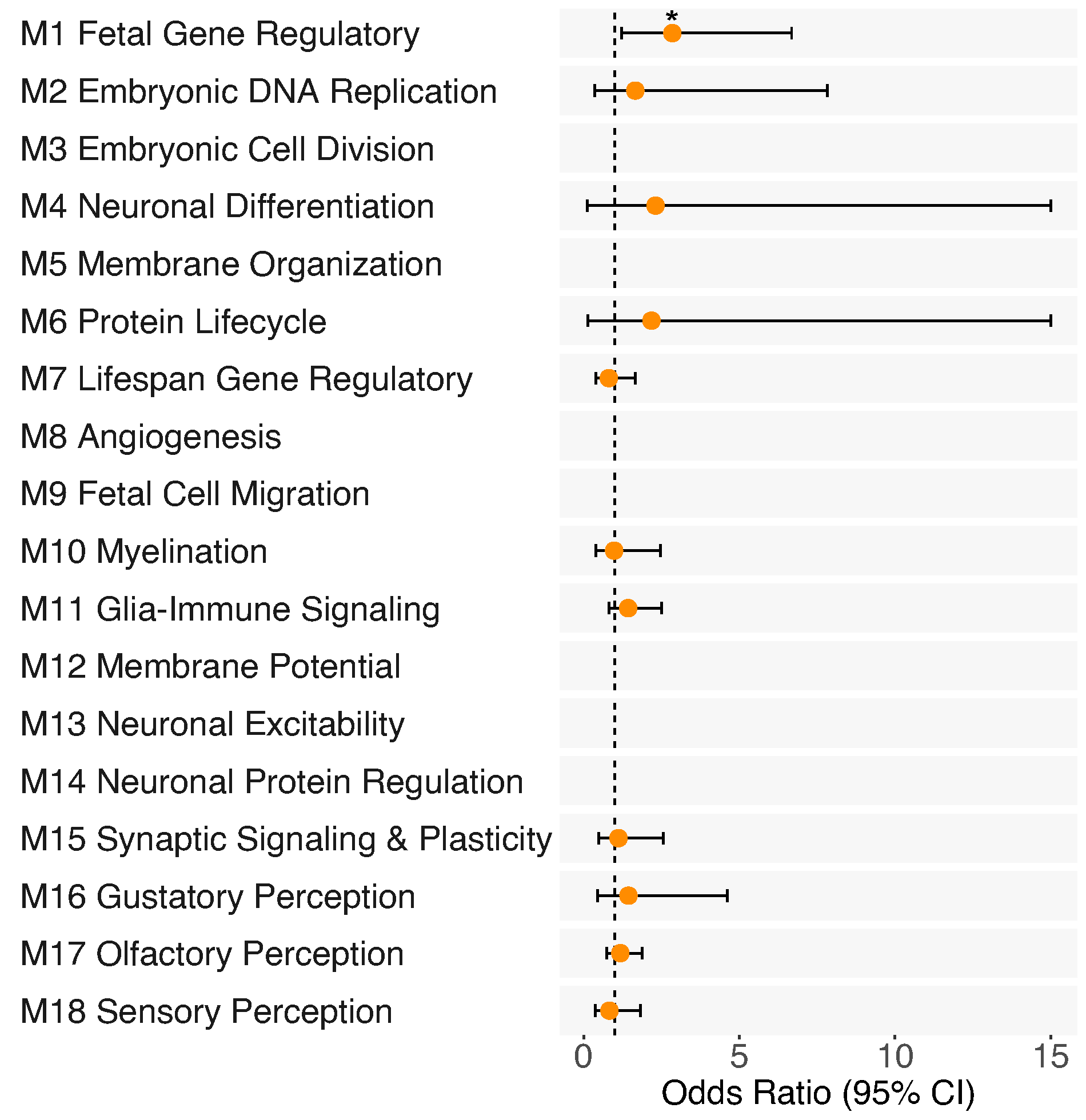

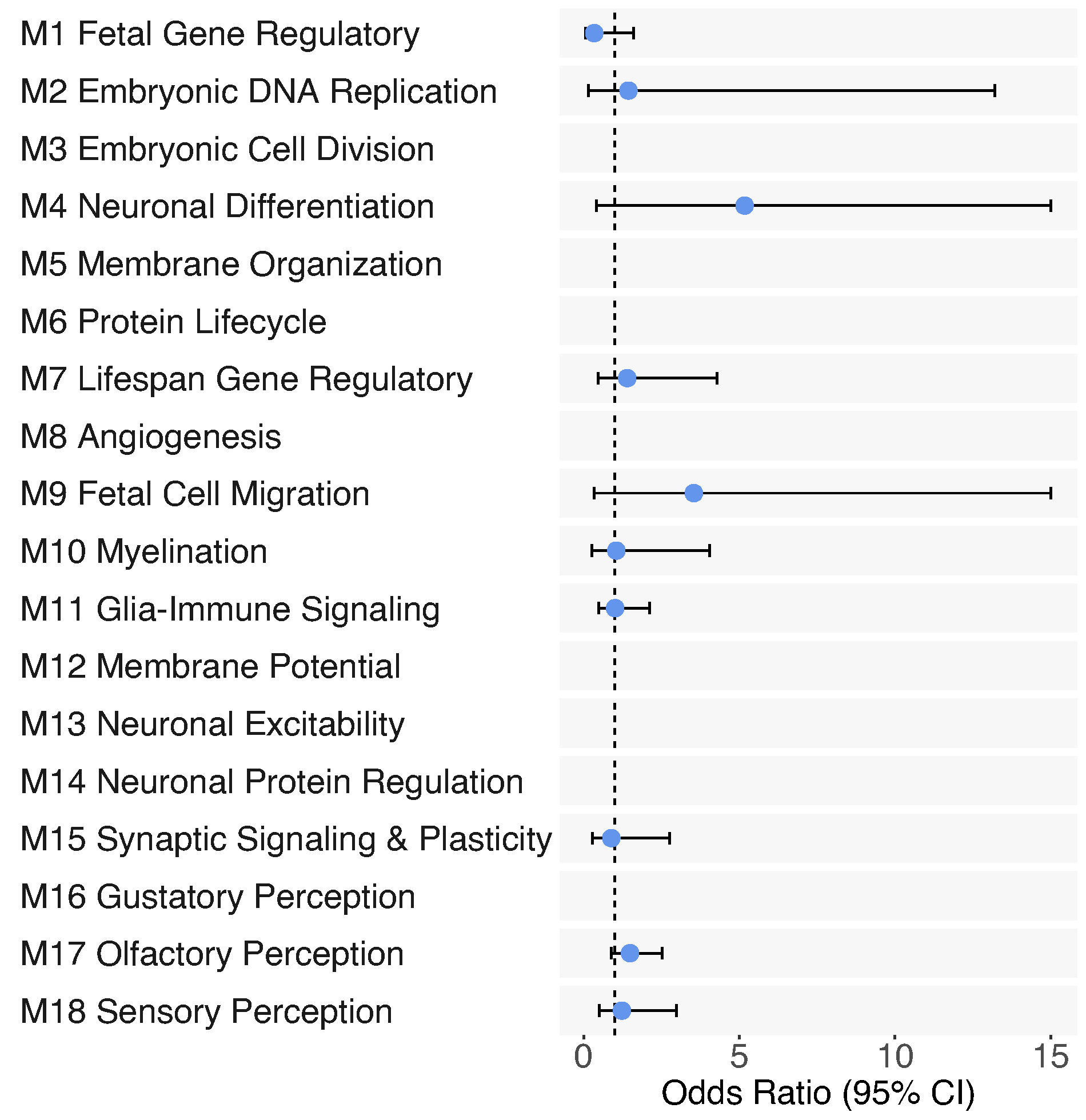

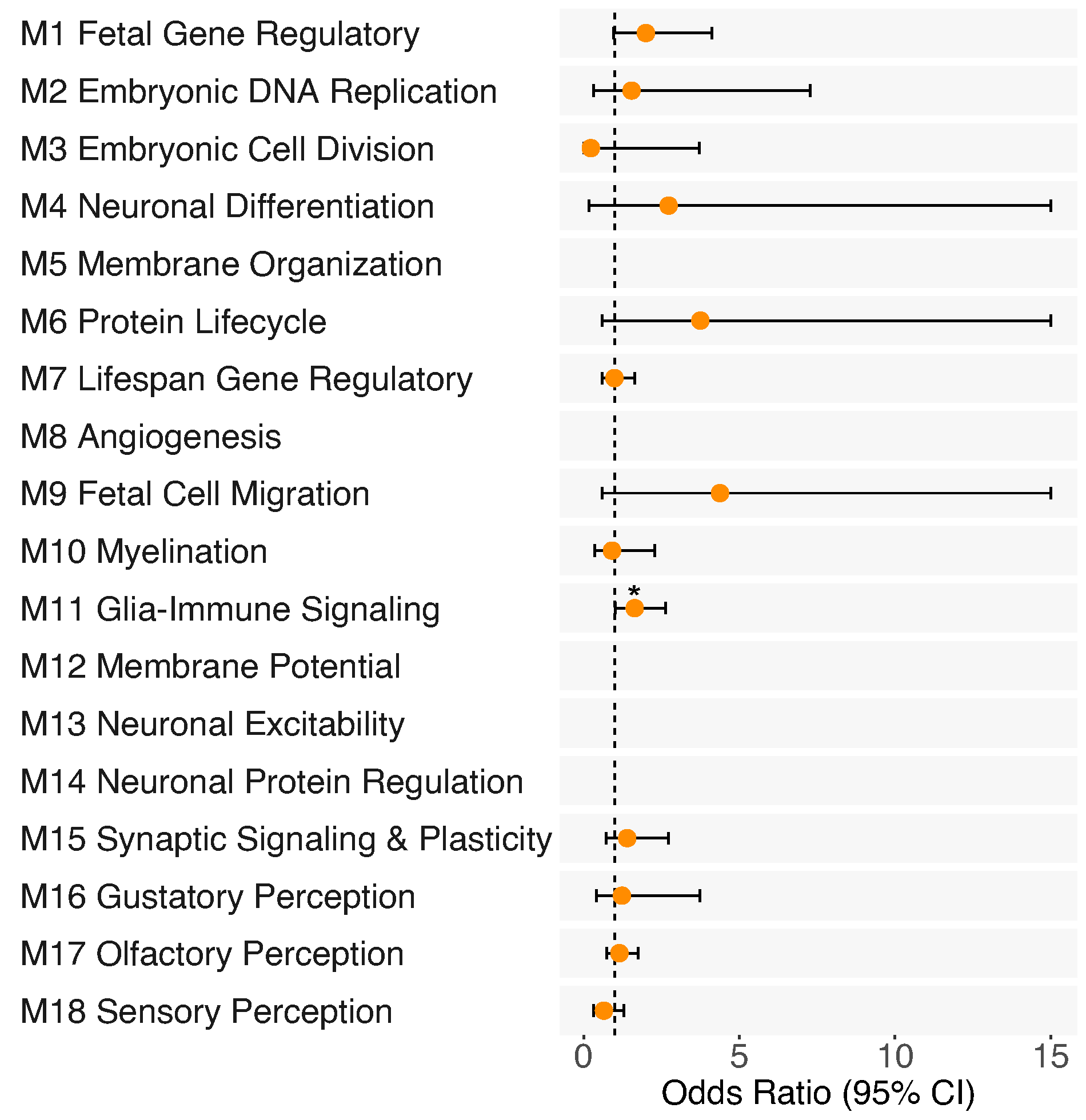

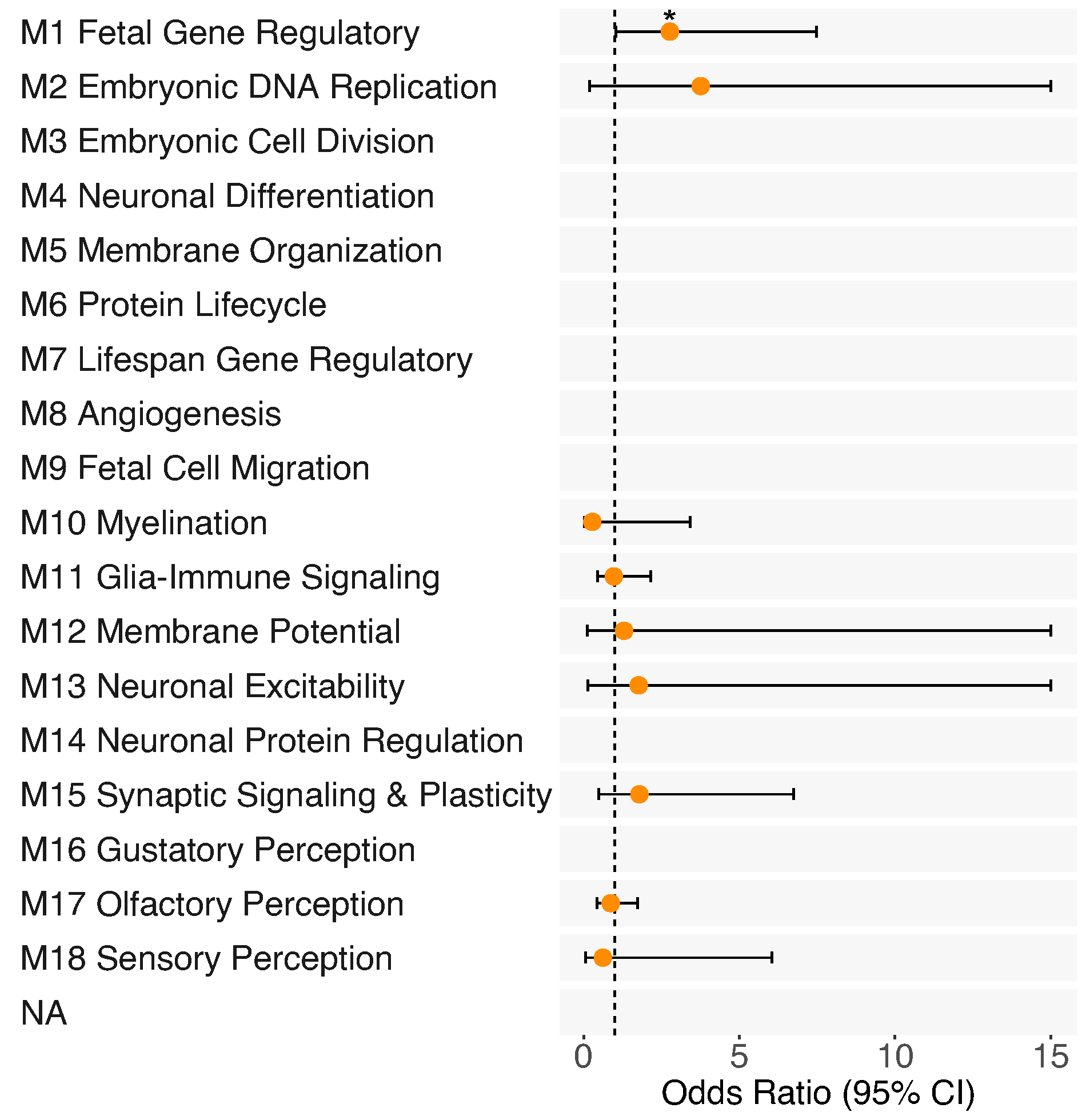

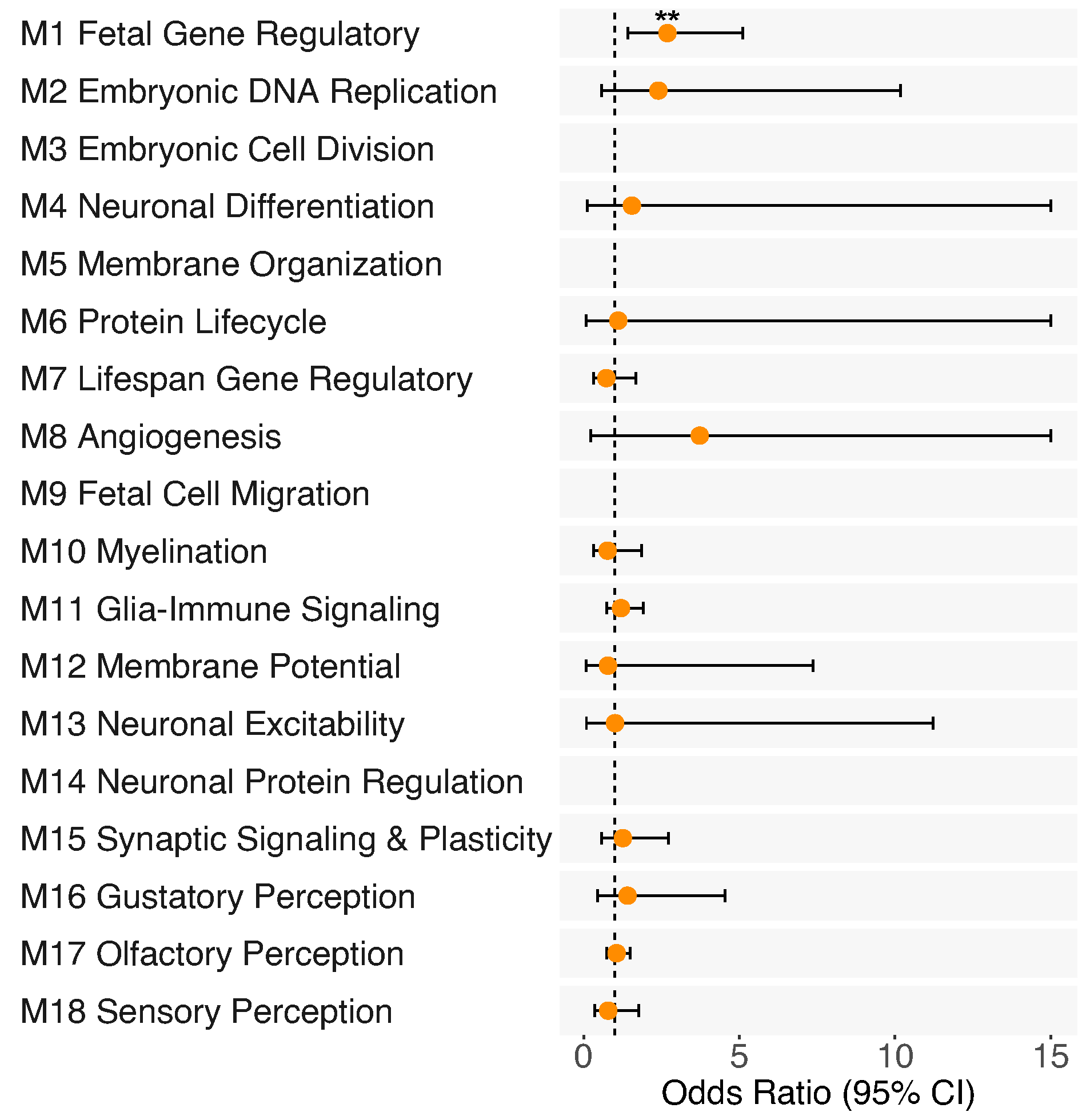

**Supplementary Figure 9.** Odds ratio (OR) associated with number of genes deleted per neurodevelopmental gene-set, controlling for number of genes deleted outside each gene-set of interest, for (A) child-onset psychosis (*n* with child-onset psychosis = 39, *n* with later-onset psychosis = 568) and for borderline IQ across (B) all SSD subjects (*n* with borderline IQ = 120, *n* without borderline IQ = 478), (C) SSD cases excluding those with known risk CNVs (*n* with borderline IQ = 113, *n* without borderline IQ = 469), (D) all non-SSD subjects (*n* with borderline IQ = 57, *n* without borderline IQ = 741), and (E) SSD cases, SSD-relatives, and controls, excluding subjects with known risk CNVs (*n* with borderline IQ = 168, *n* without borderline IQ = 1210). **p* < 0.05. **FDR q < 0.05, corrected for number of independent variables tested per trait. Max OR confidence interval shown = 15; see Extended Table 11 for full statistics.

**Supplementary Figure 10.** Excluding subjects with known risk CNVs, associations between M1 deletions weighted by LOEUF score and (A) gray matter volume, (B) cortical thickness, and (C) surface area centiles for SSD subjects (*n* = 277) and across SSD subjects, SSD-relatives (*n* = 105), and controls (*n* = 313; D-F). To facilitate interpretability, relationships are plotted using raw centile and M1 LOEUF sum values; reported effect sizes and p-values are from the mixed models.

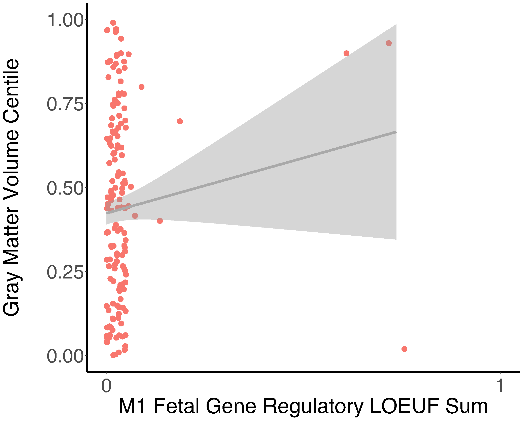

*b* = 0.35, *p* = .09

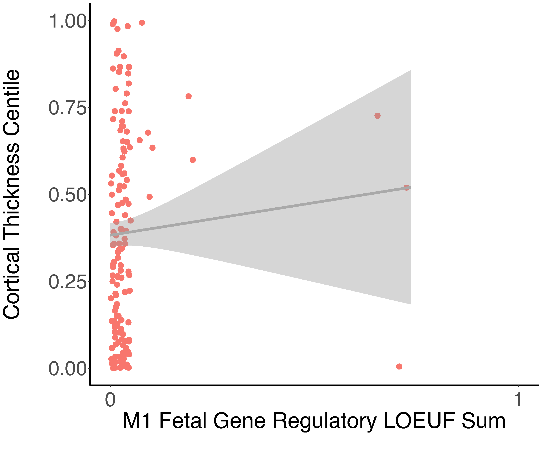

*b* = 0.17, *p* = .46

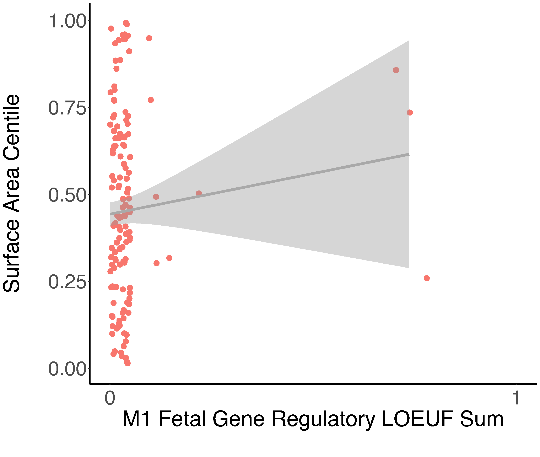

*b* = 0.27, *p* = .22

**C**

**A**

**B**

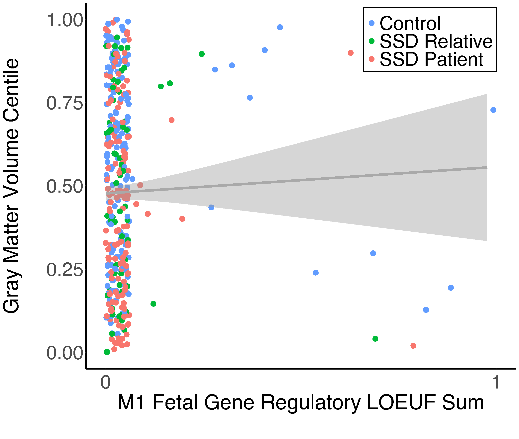

*b* = 0.08, *p* = .53

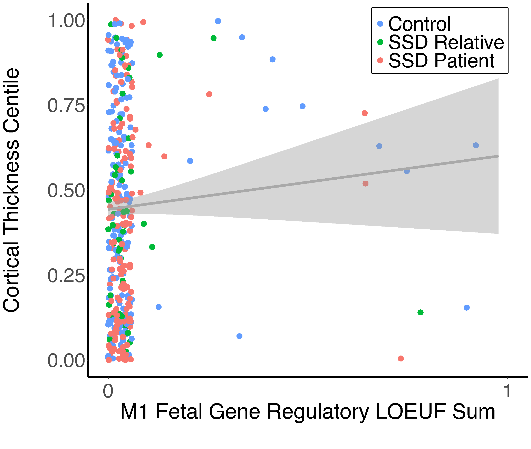

*b* = 0.21, *p* = .15

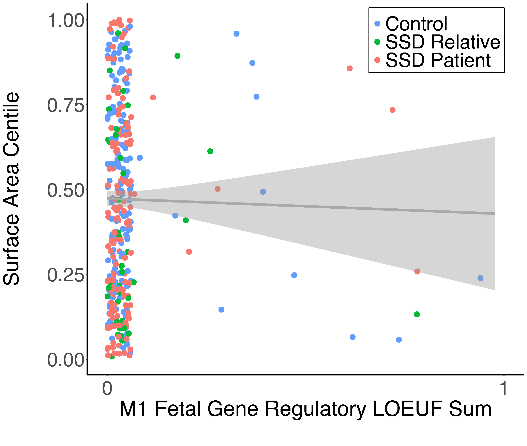

*b* = -0.08, *p* = .58

**D**

**E**

**F**

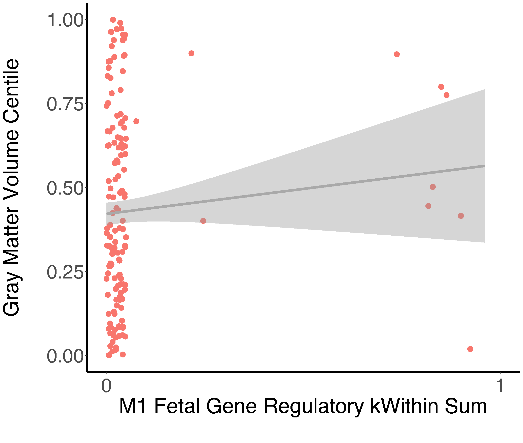

*b* = 0.14, *p* = .14

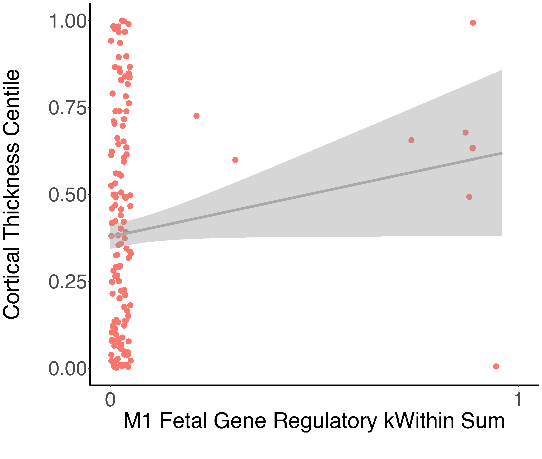

*b* = 0.15, *p* = .14

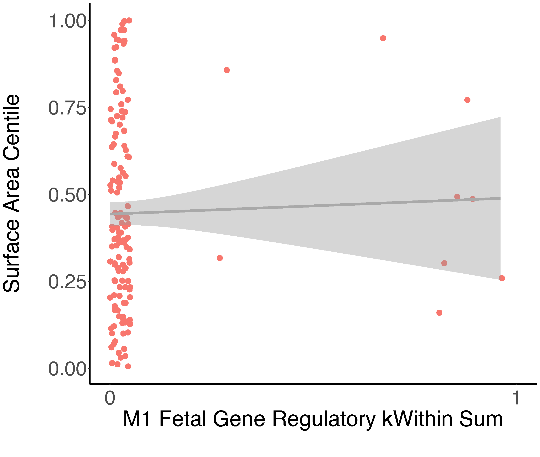

*b* = 0.06, *p* = .56

**C**

**A**

**B**

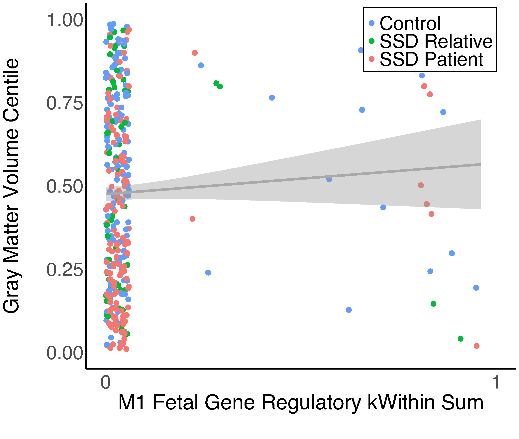

*b* = 0.08, *p* = .21

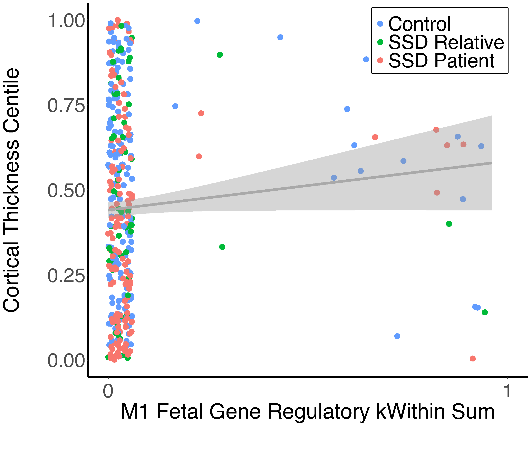

**b* = 0.14, *p* = .048

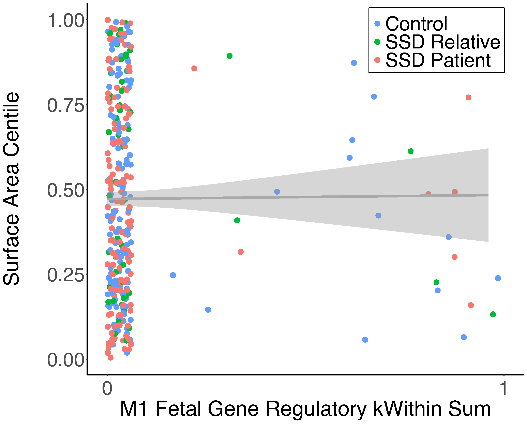

*b* = -0.003, *p* = .96

**D**

**E**

**F**

**Supplementary Figure 11.** Excluding subjects with known risk CNVs, associations between M1 deletions weighted by kWithin score and (A) gray matter volume, (B) cortical thickness, and (C) surface area centiles for SSD subjects (*n* = 277) and across SSD subjects, SSD-relatives (*n* = 105), and controls (*n* = 313; D-F). To facilitate interpretability, relationships are plotted using raw centile and M1 kWithin sum values; reported effect sizes and p-values are from the mixed models.

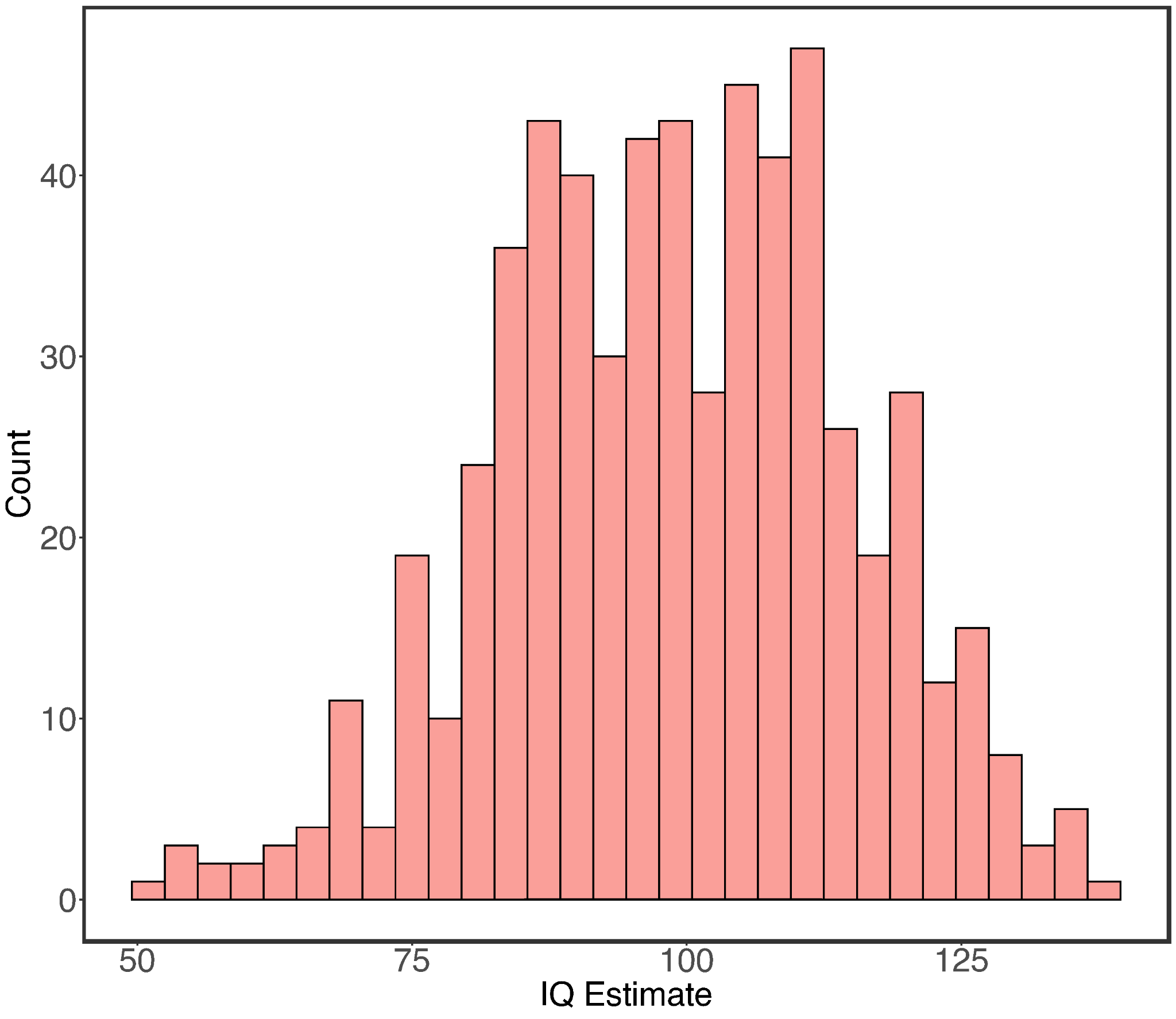

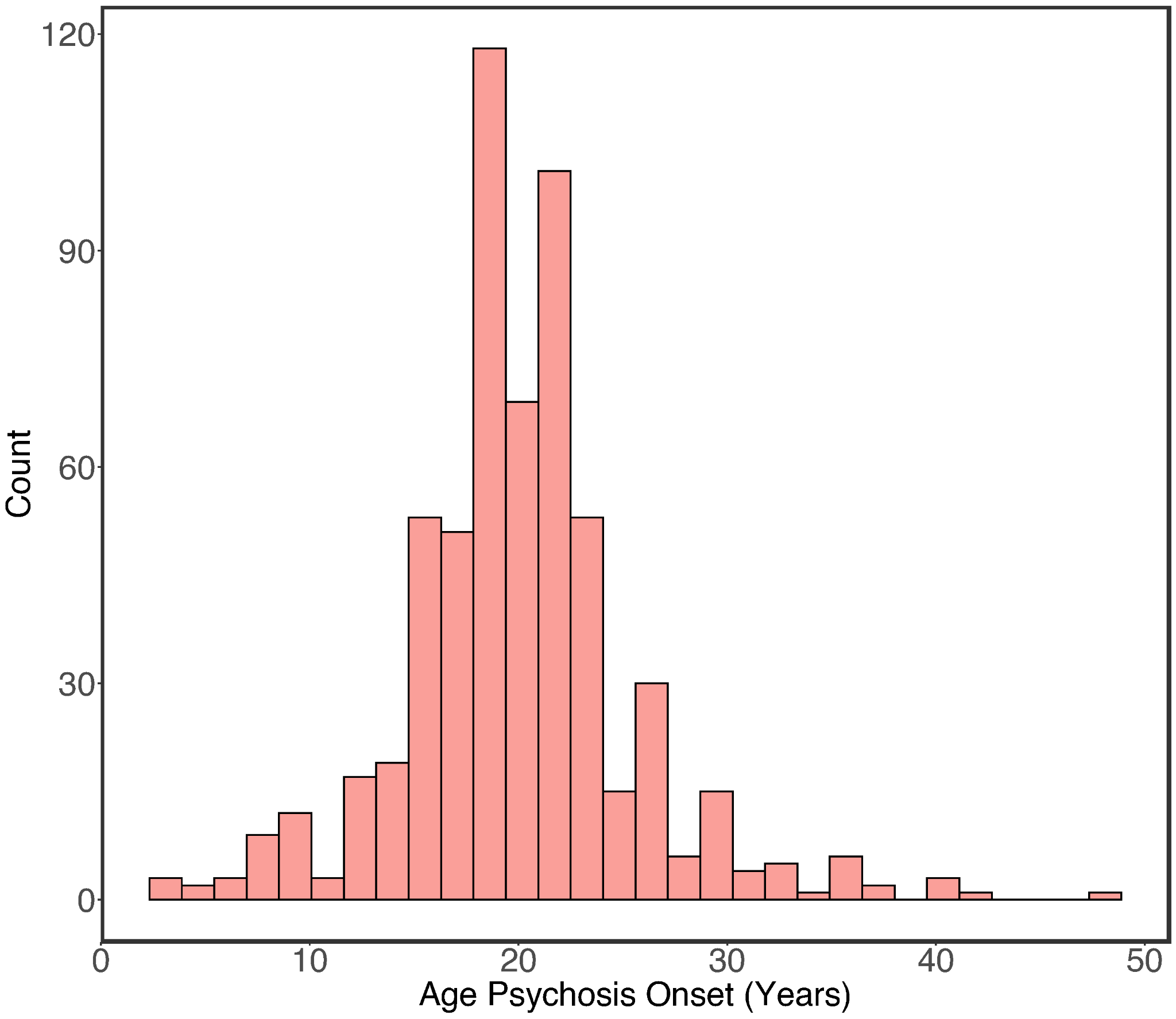

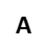

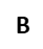

**Supplementary Figure 12.** Distribution of (A) IQ estimates and (B) age of psychosis onset among SSD patients.

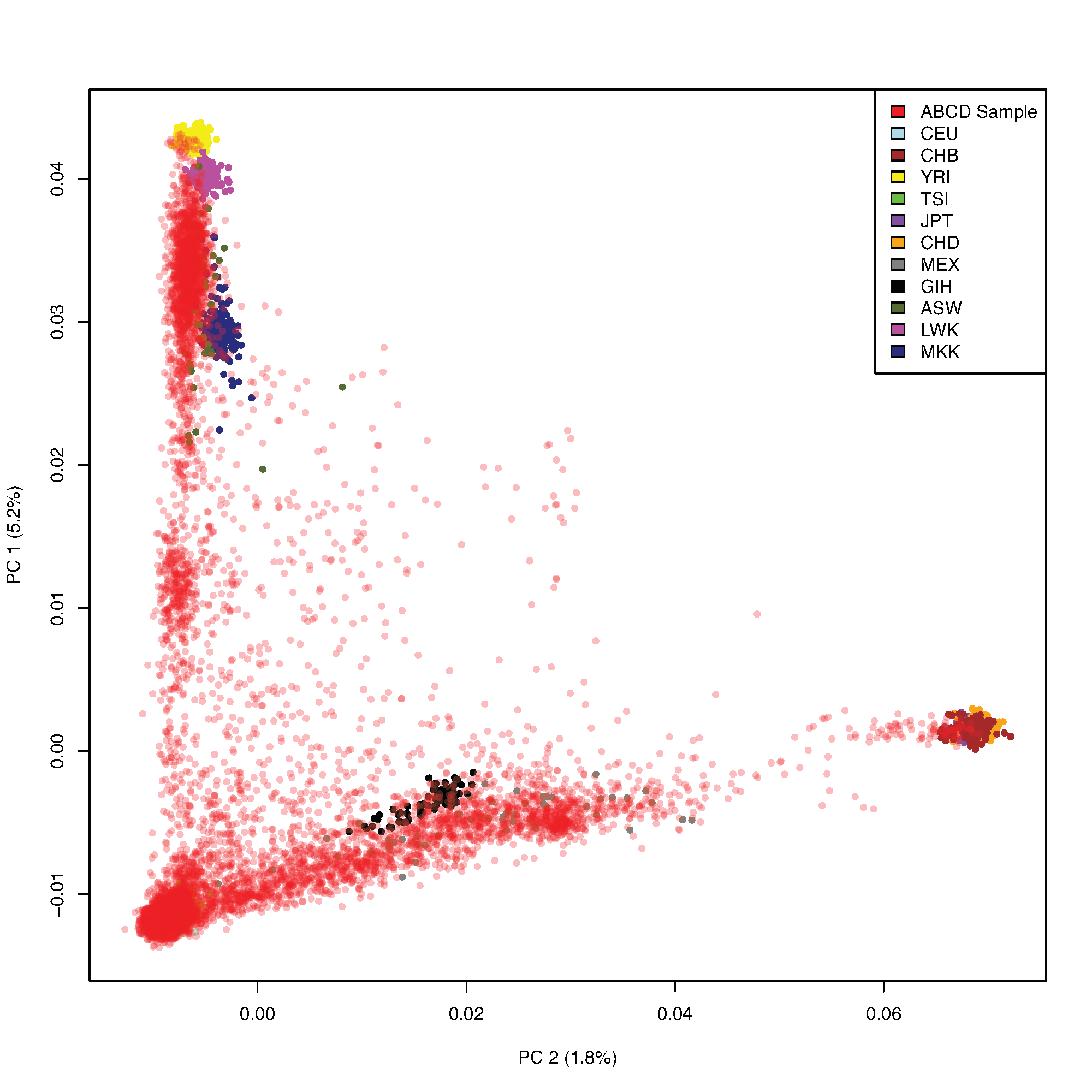

**Supplementary Figure 13.** Top two ancestry principal components (PCs) in the ABCD study cohort, overlaid on HapMap3 global populations (CEU = Utah residents with Northern and Western European ancestry, CHB = Han Chinese in Beijing, China, YRI = Yoruba in Ibadan, Nigeria, TSI = Toscans in Italy, JPT = Japanese in Tokyo, Japan, CHD = Chinese in Denver, Colorado, MEX = Mexican ancestry in Los Angeles, California, GIH = Gujarati Indians in Houston, Texas, ASW = African ancestry in Southwest USA, LWK = Luhya in Webuye, Kenya, MKK = Maasai in Kinyawa, Kenya).

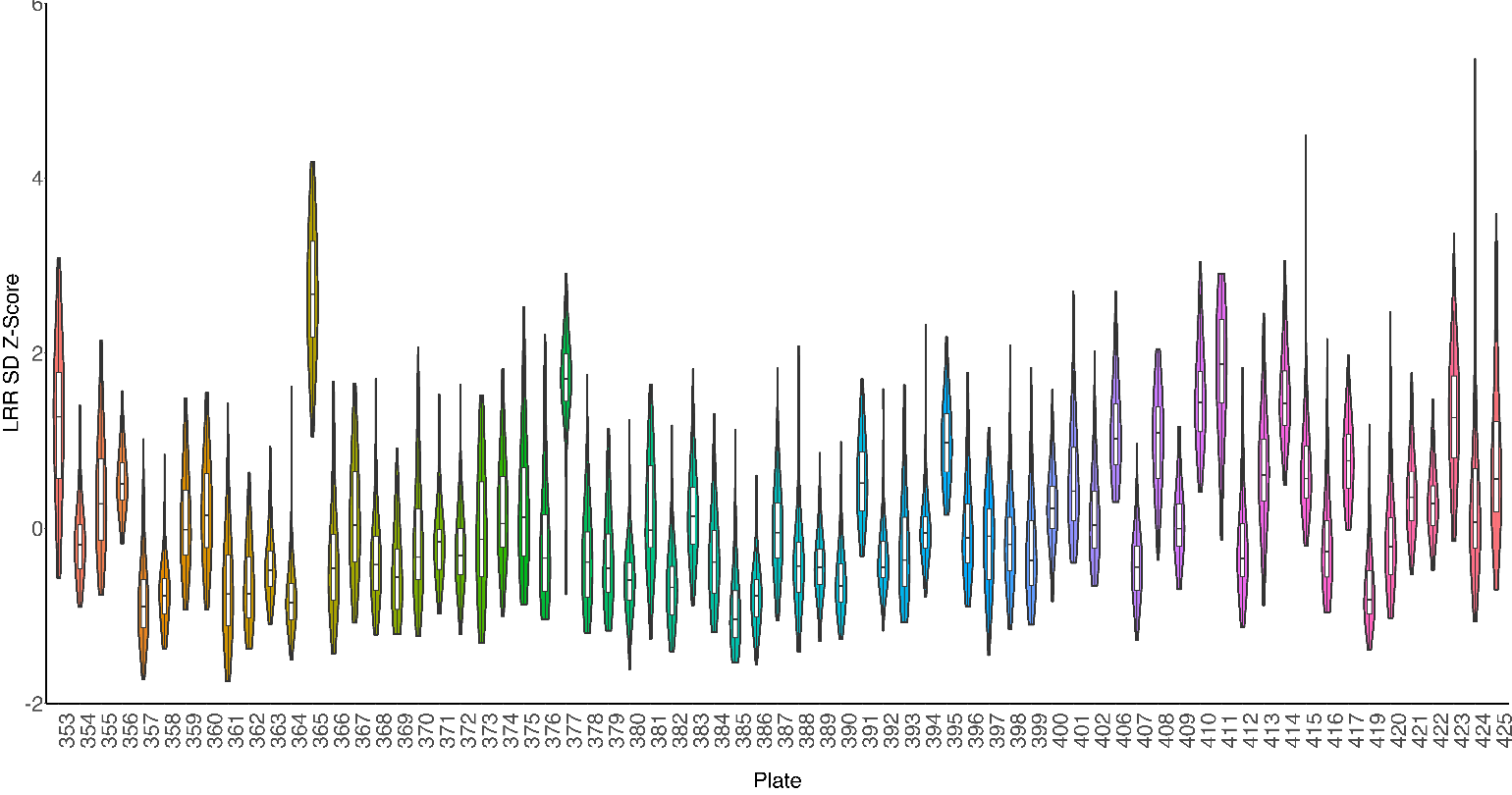

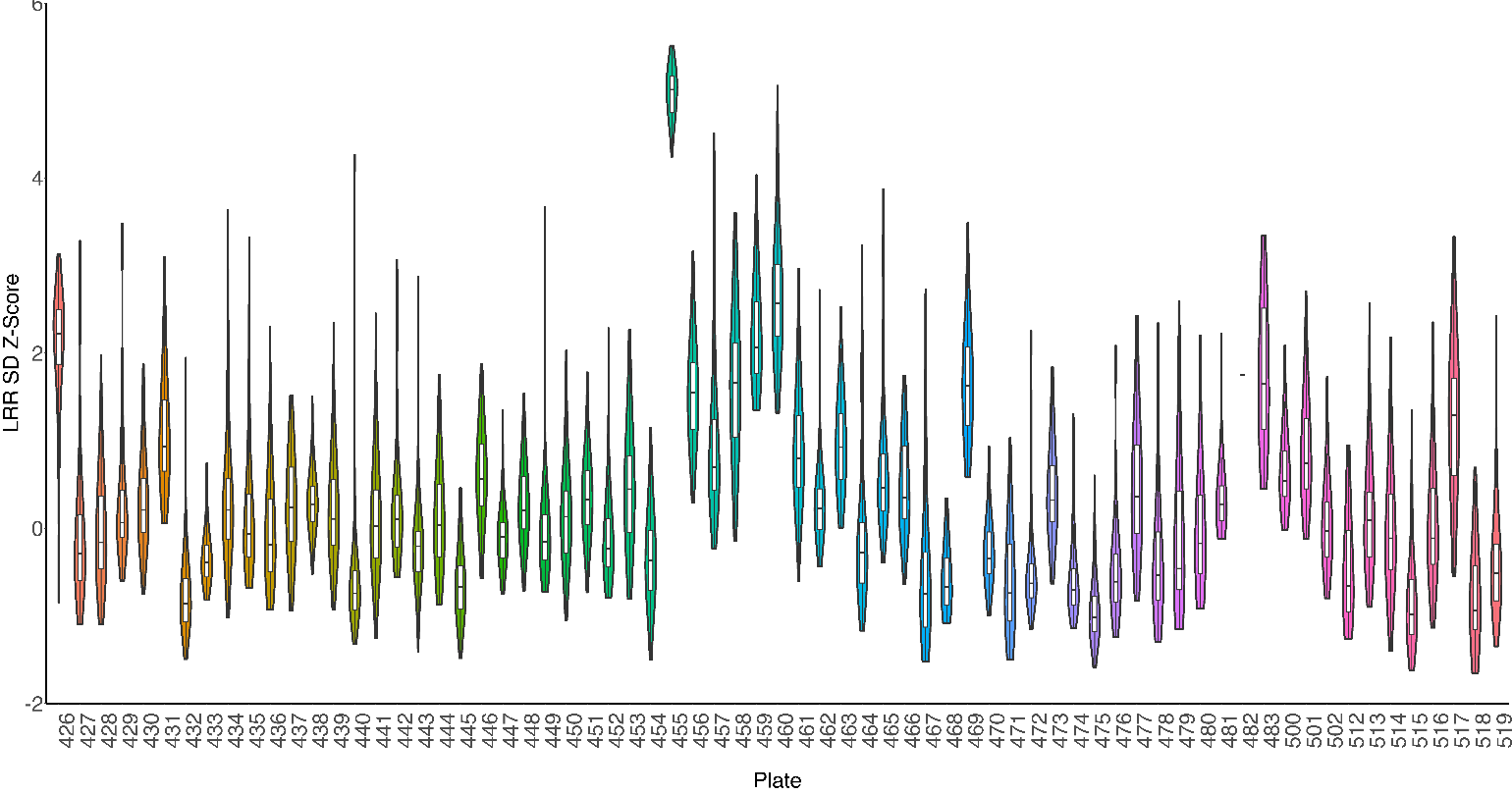

**Supplementary Figure 14.** Distribution of Log R Ratio standard deviation (LRR SD) z-scores across subjects/samples per genotyping plate in the ABCD study. Boxplots mark the median LRR SD z-score per sample per plate, with lower and upper hinges corresponding to the 25^th^ and 75^th^ quartiles.

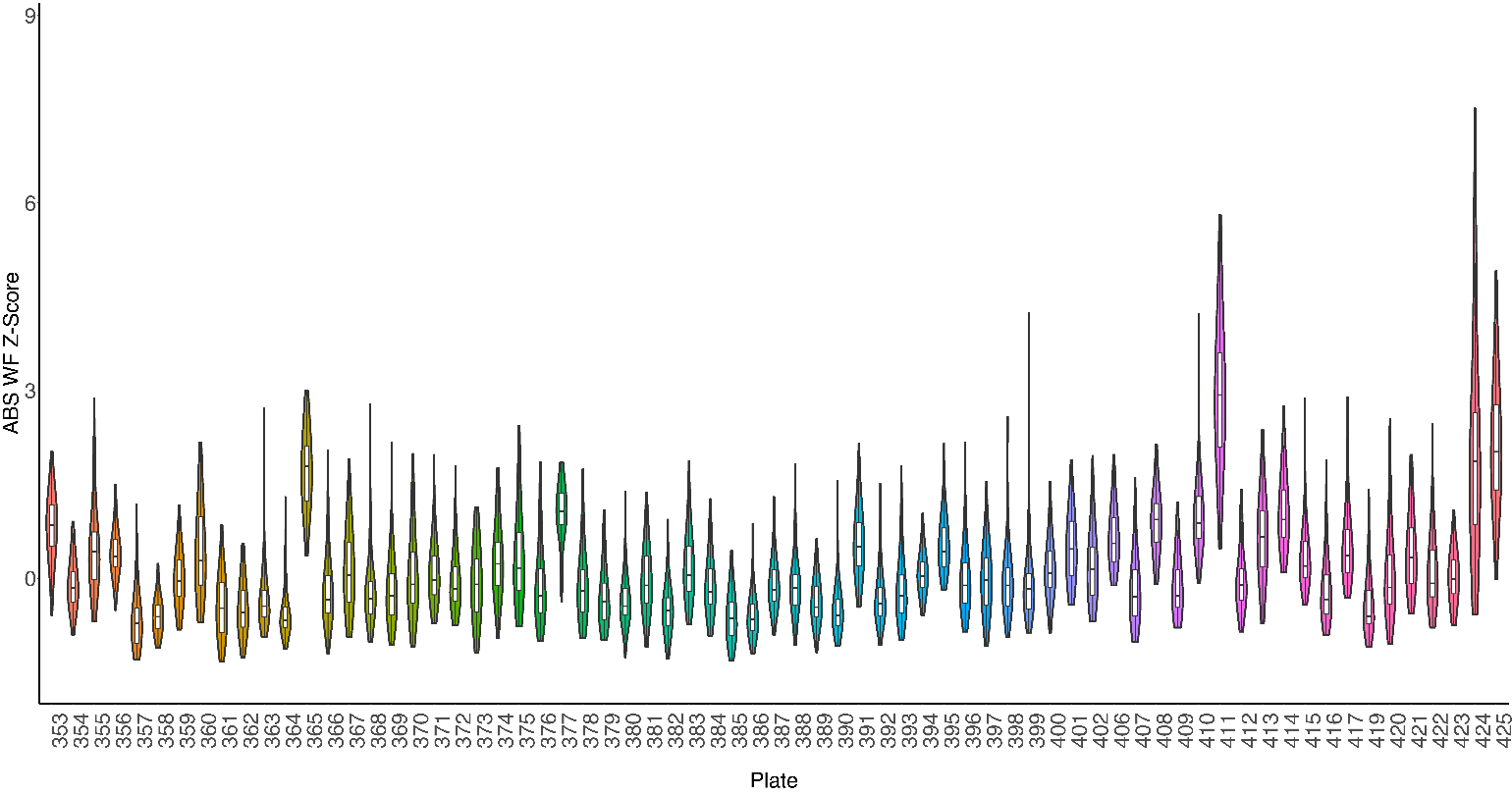

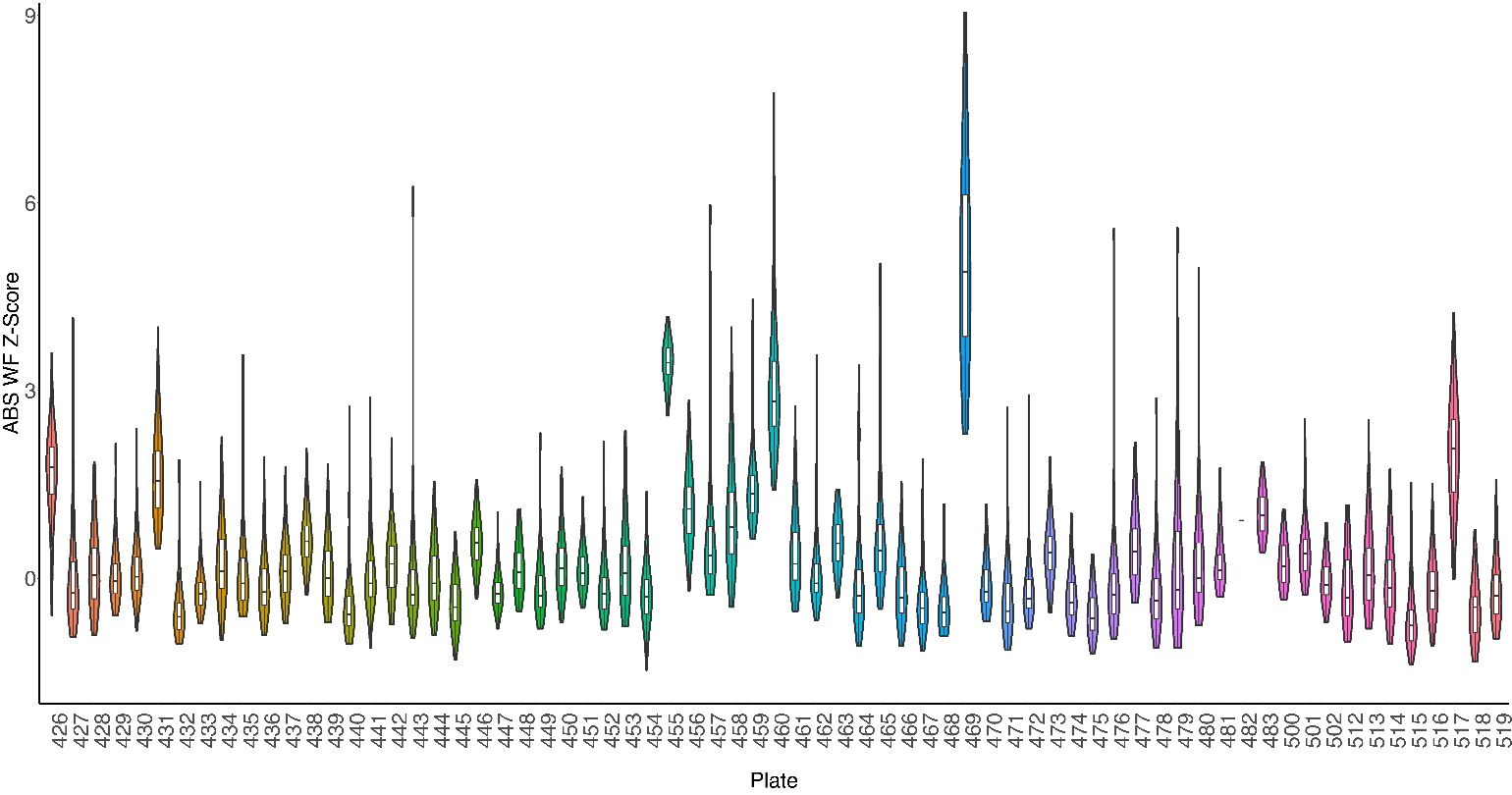

**Supplementary Figure 15.** Distribution of absolute waviness factor (ABS WF) z-scores across subjects/samples per genotyping plate in the ABCD study. Boxplots mark the median ABS WF z-score per sample per plate, with lower and upper hinges corresponding to the 25^th^ and 75^th^ quartiles.

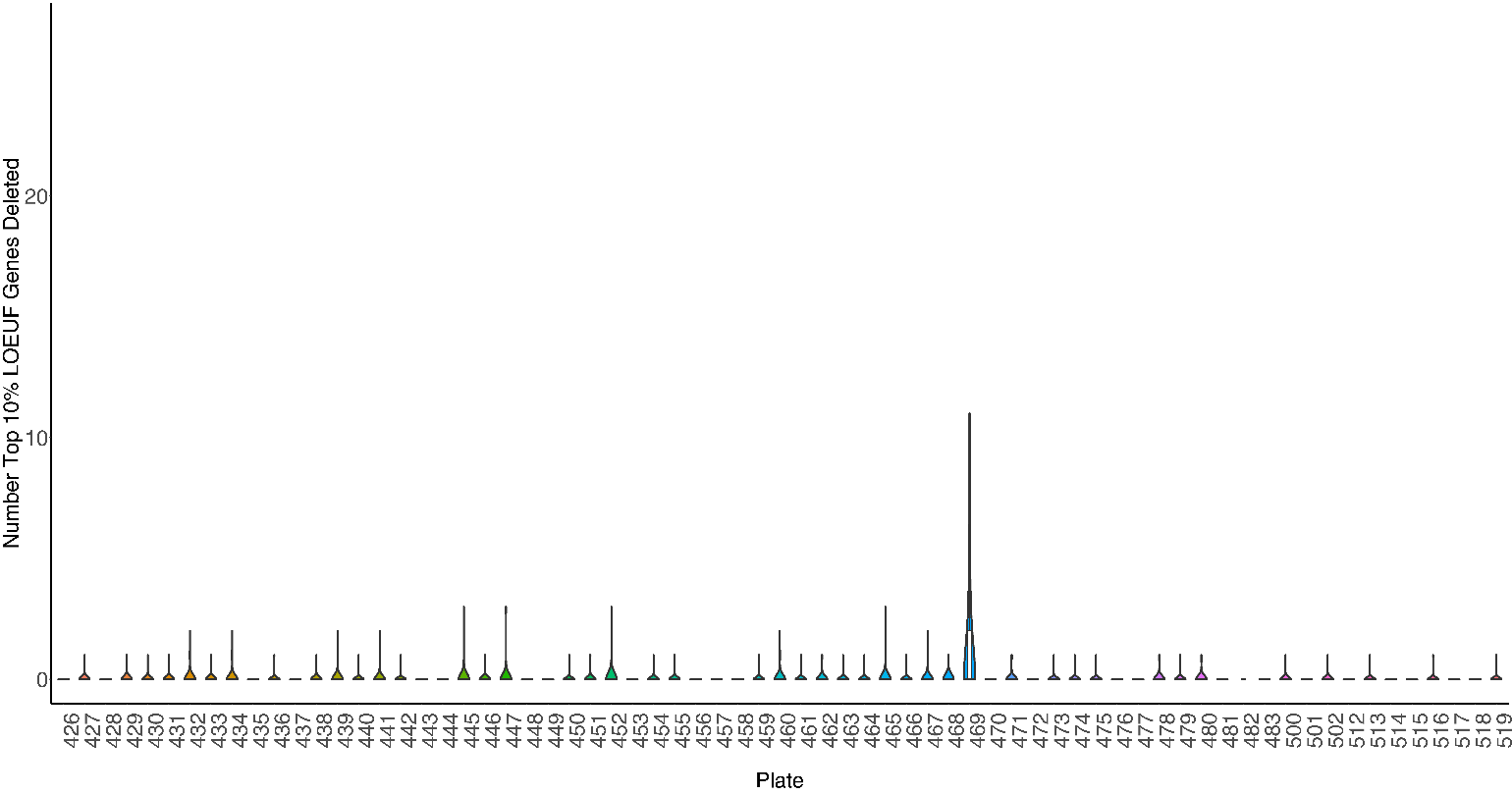

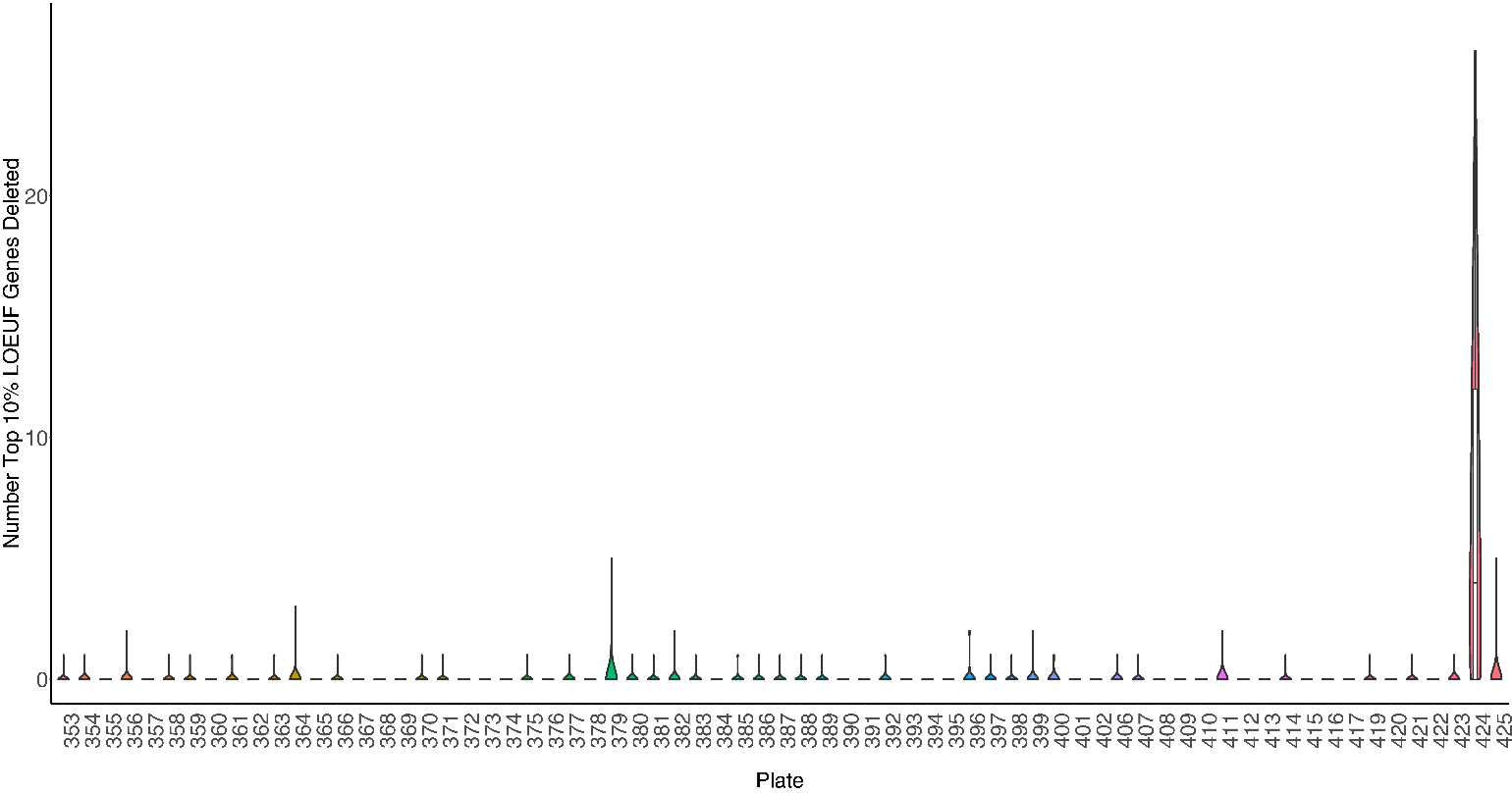

**Supplementary Figure 16**. Distribution of number of top 10% LOEUF genes (i.e., genes that are highly intolerant to LOF) deleted across subjects/samples per genotyping plate in the ABCD study. Boxplots mark the median number of top 10% LOEUF genes deleted per sample per plate, with lower and upper hinges corresponding to the 25^th^ and 75^th^ quartiles
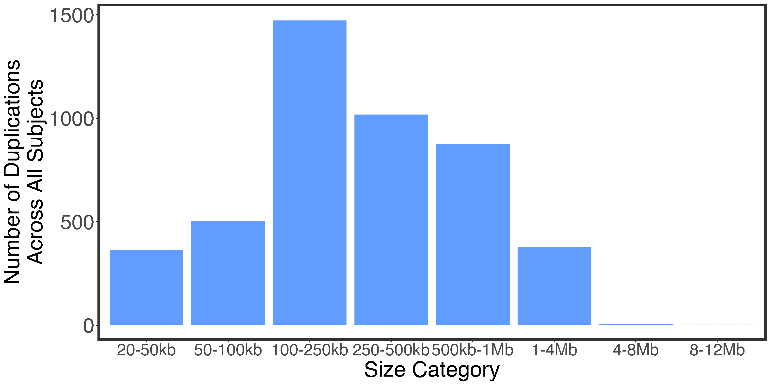

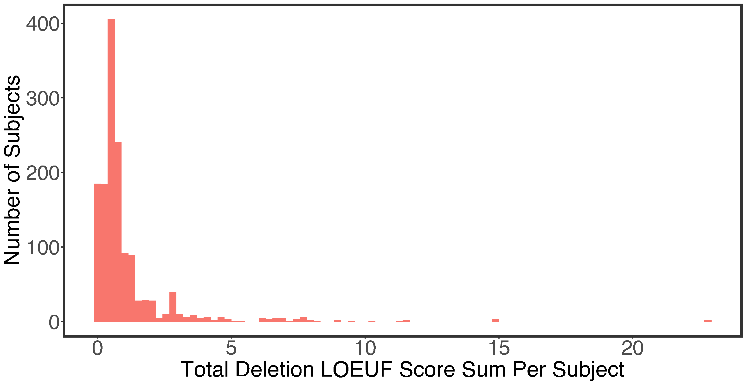

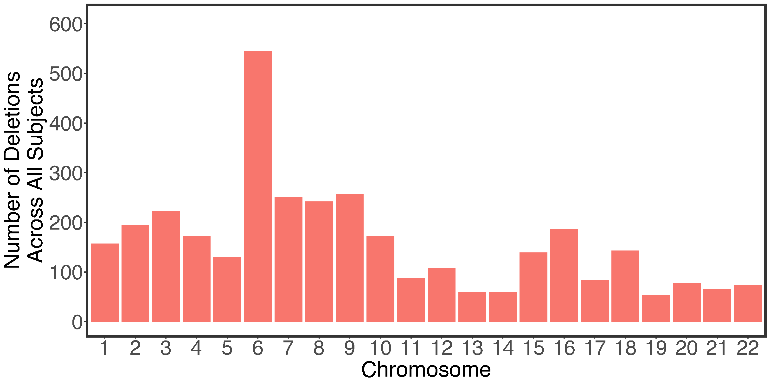

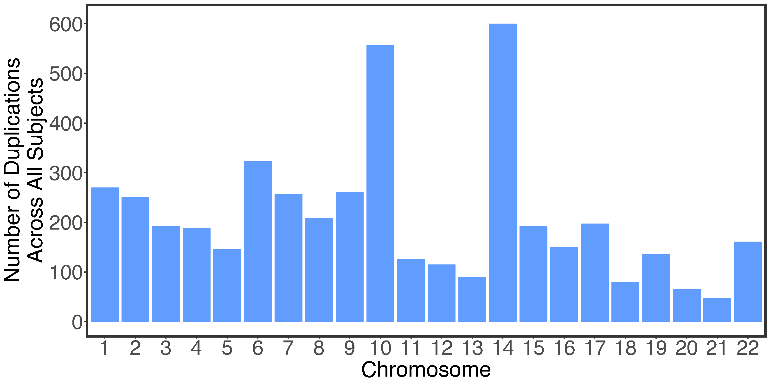

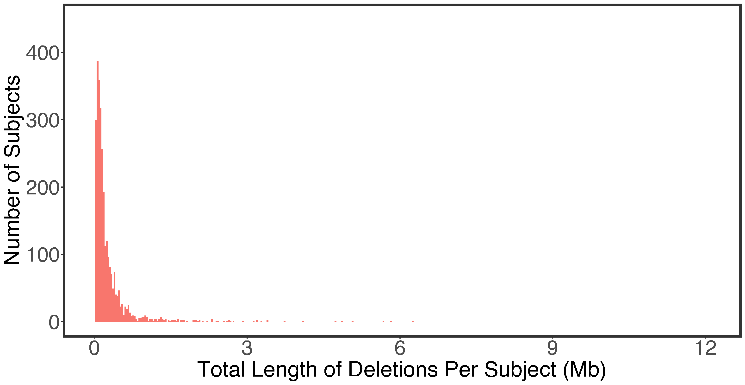

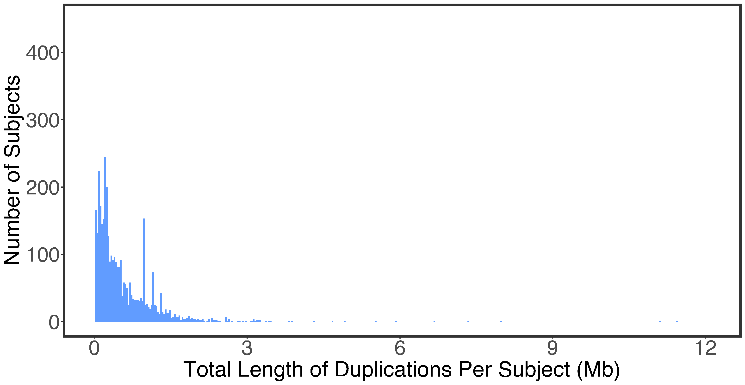

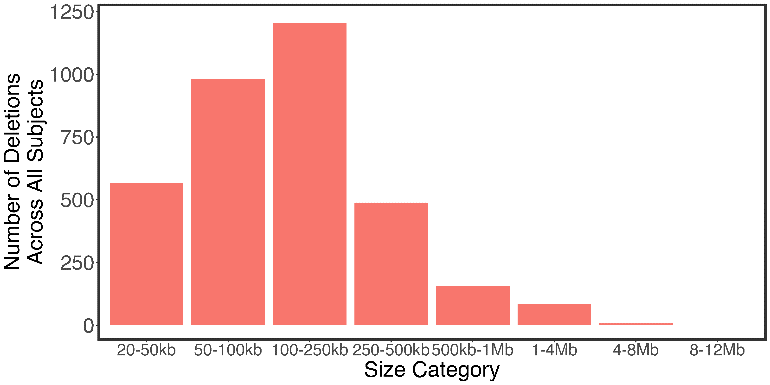

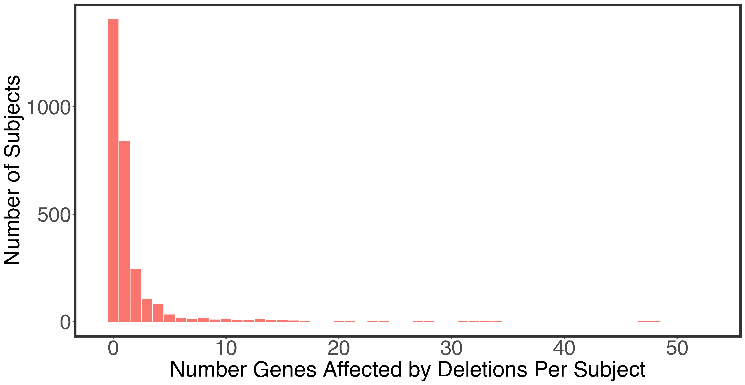

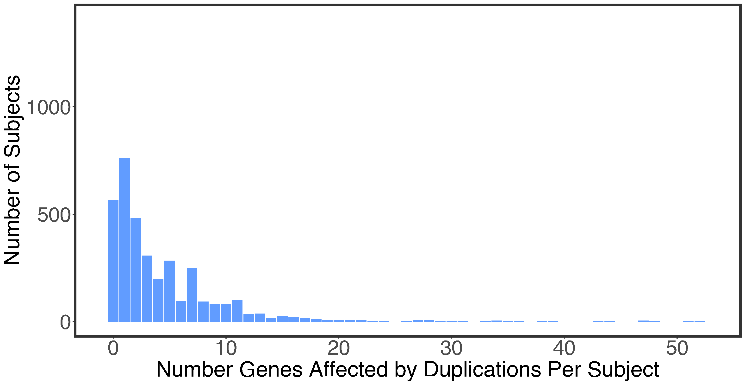

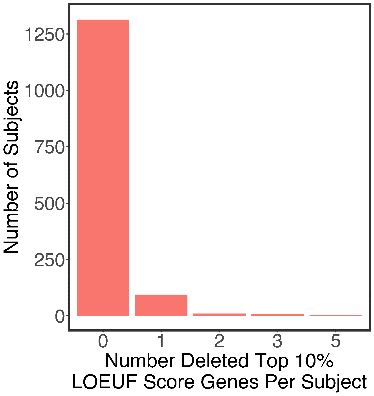

**Supplementary Figure 17.** CNV characteristics in ABCD cohort using main CNV calls.
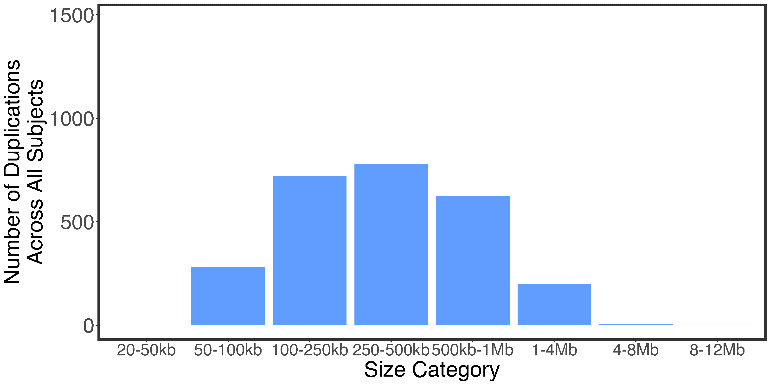

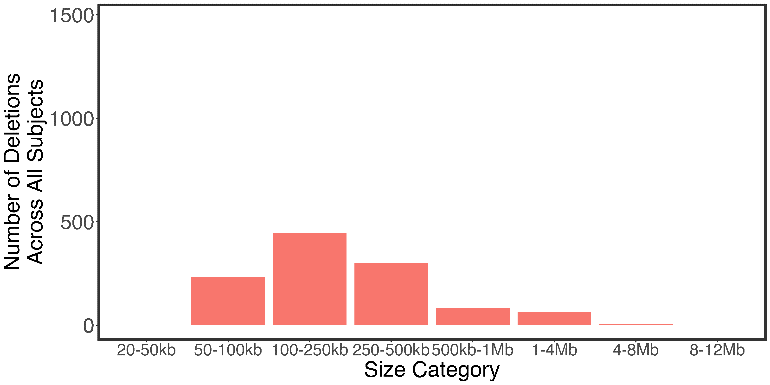

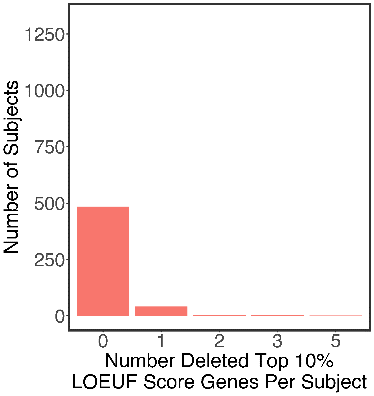

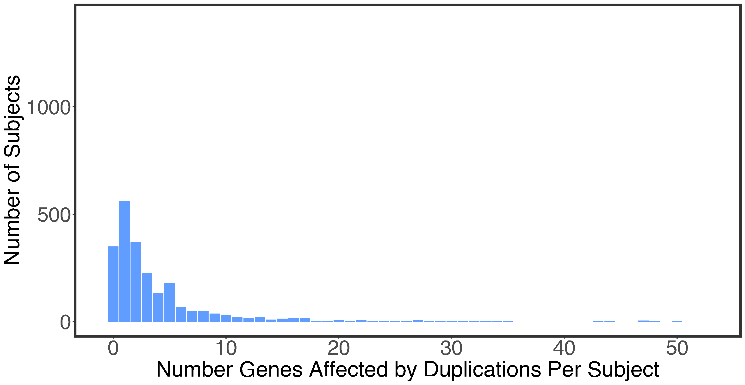

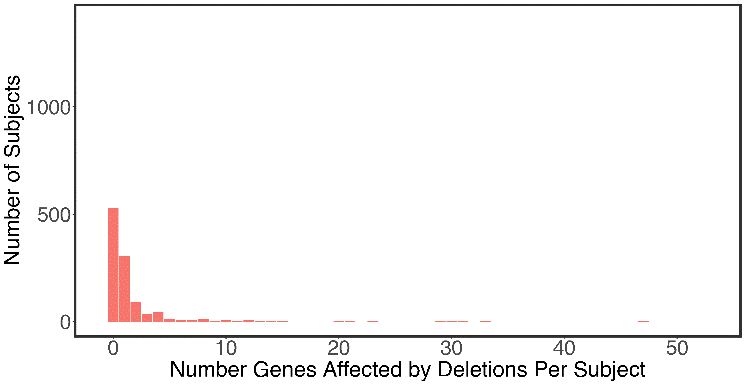

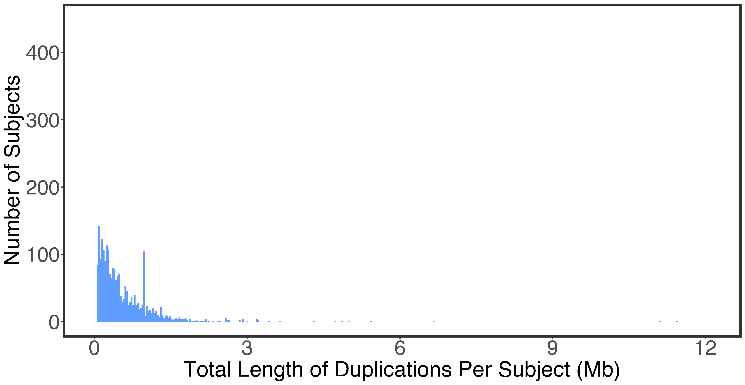

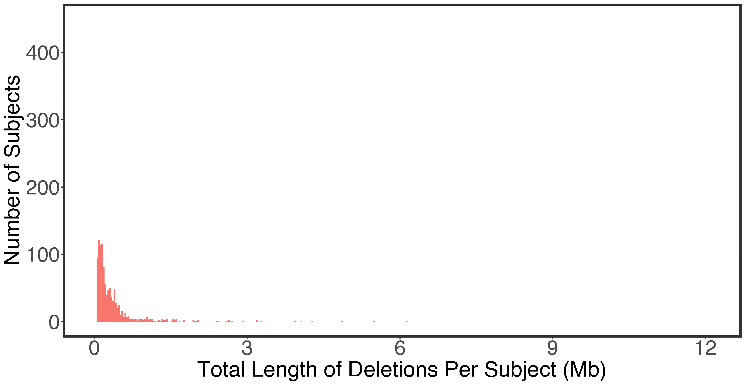

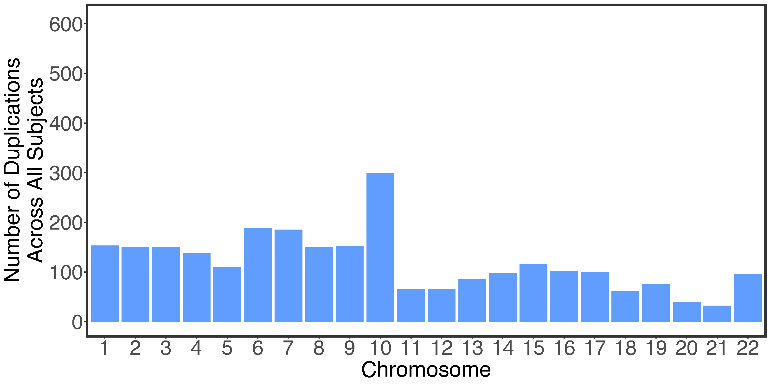

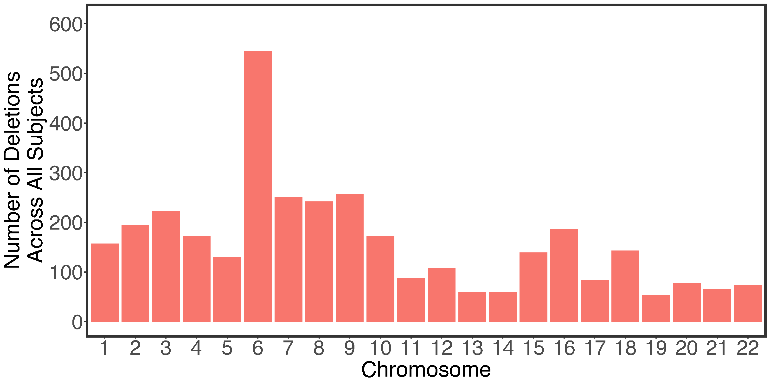

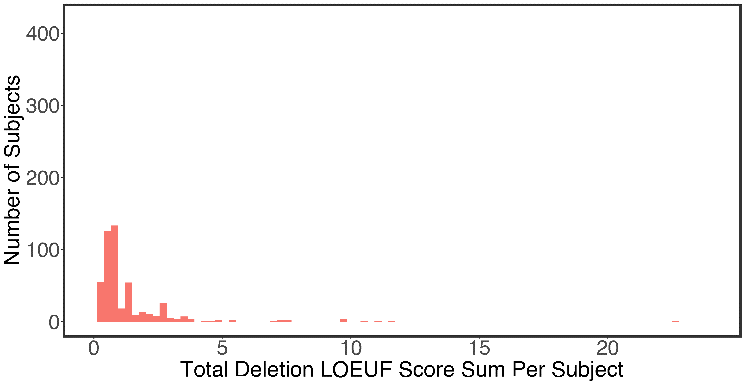

**Supplementary Figure 18.** CNV characteristics in ABCD cohort using Sha et al., 2025 (39) calls.

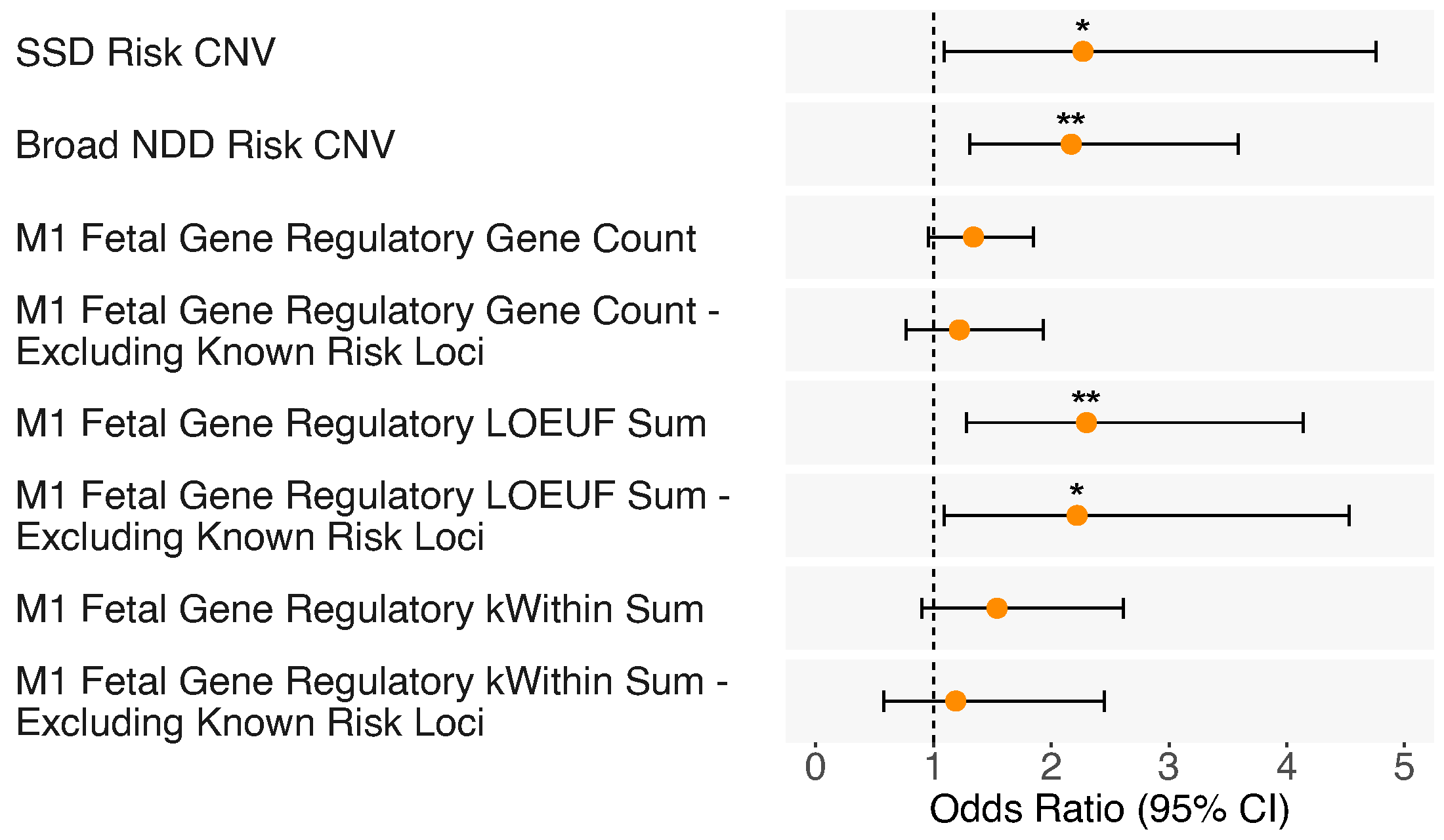

**Supplementary Figure 19.** Using conservative CNV calls from Sha et al., (2025), odds ratio (OR) for borderline intellectual functioning in ABCD study subjects with SSD- and broad NDD-risk CNVs, number of M1 genes deleted, M1 deletion burden weighted by LOEUF score, and M1 deletion burden weighted by kWithin score, including (*n* = 9,930) or excluding subjects with known risk CNVs (*n* = 9,869). **p*<0.05; ***q*<0.05, corrected for number of scores tested.

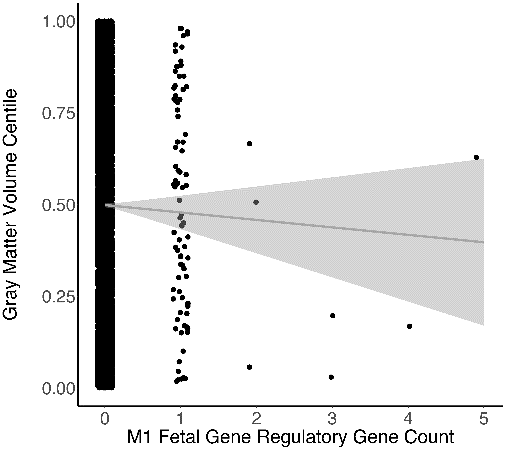

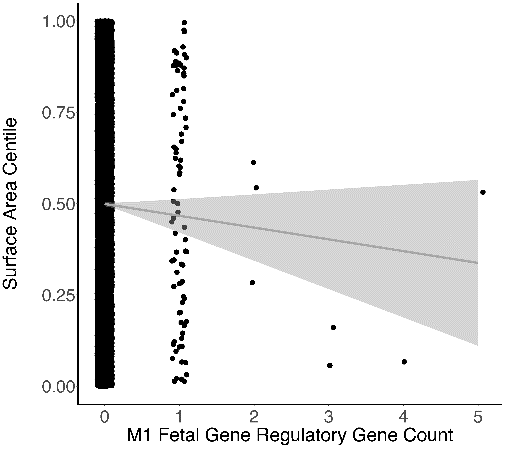

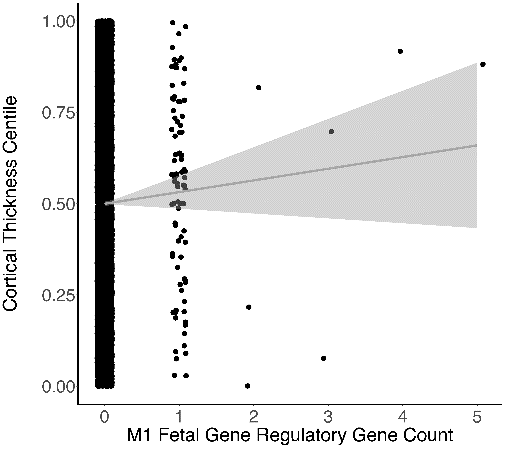

*b* = -0.03, *p* = .20

*b* = 0.02, *p* = .39

*b* = -0.03, *p* = .15

**A**

**C**

**B**

**G**

**I**

**H**

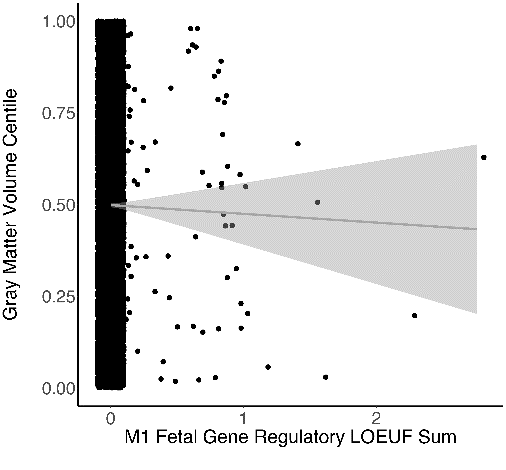

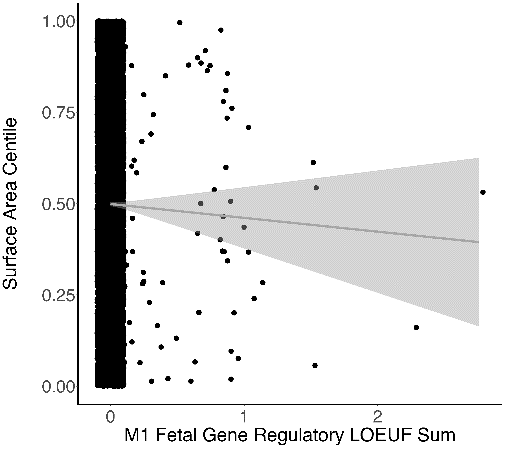

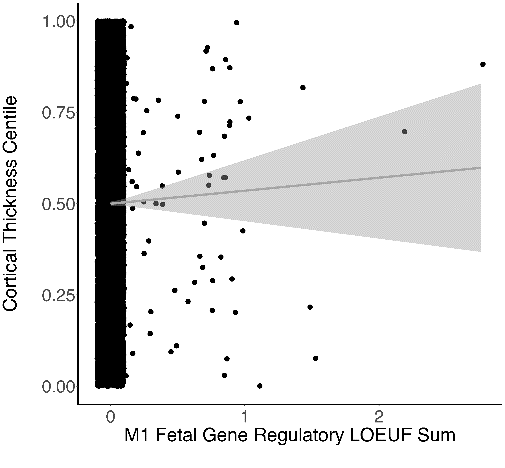

*b* = -0.05, *p* = .24

*b* = 0.01, *p* = .72

*b* = -0.05, *p* = .21

**L**

**K**

**J**

**F**

**E**

**D**

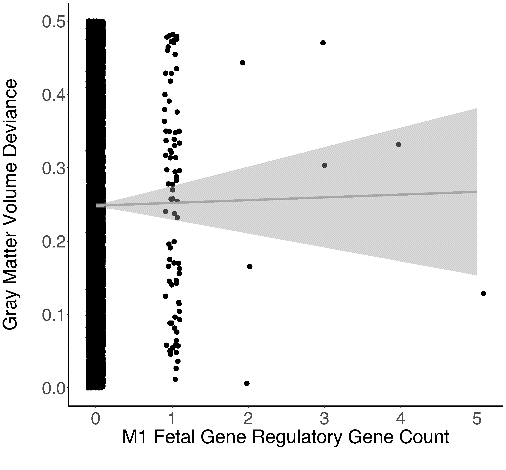

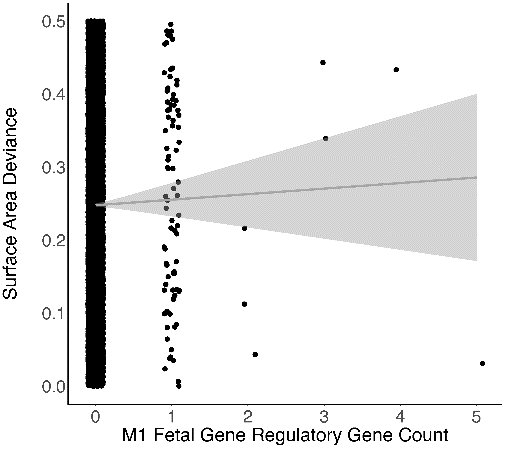

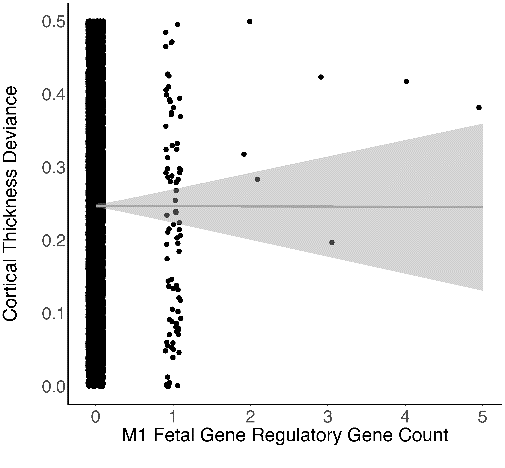

*b* = 0.00, *p* = .78

*b* = 0.00, *p* = .94

*b* = 0.00, *p* = .70

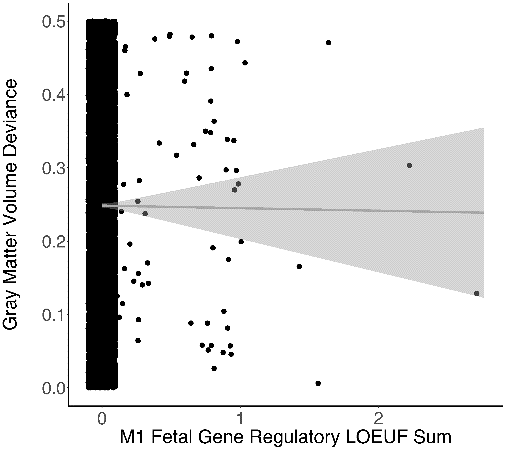

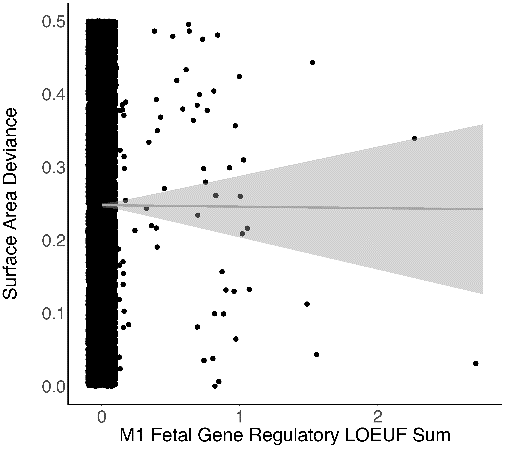

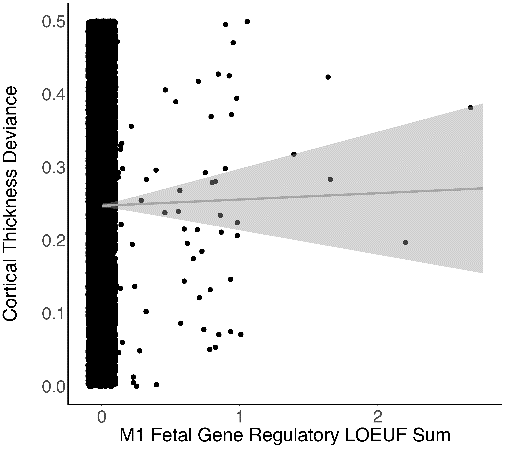

*b* = 0.00, *p* = .92

*b* = 0.01, *p* = .64

*b* = 0.00, *p* = .93

**R**

**Q**

**P**

**M**

**O**

**N**

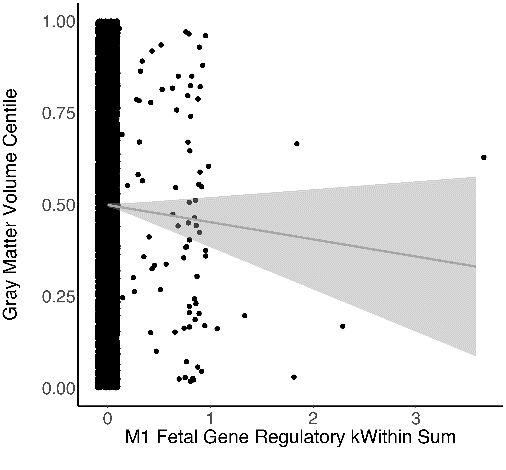

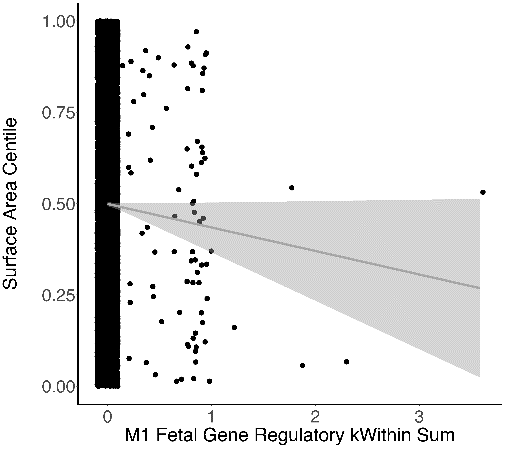

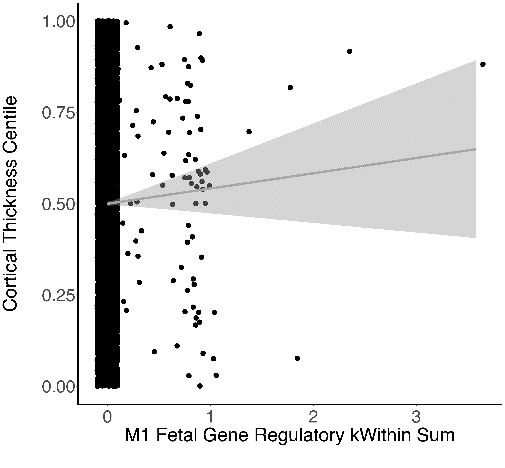

*b* = -0.04, *p* = .18

*b* = 0.03, *p* = .41

*b* = -0.05, *p* = .13

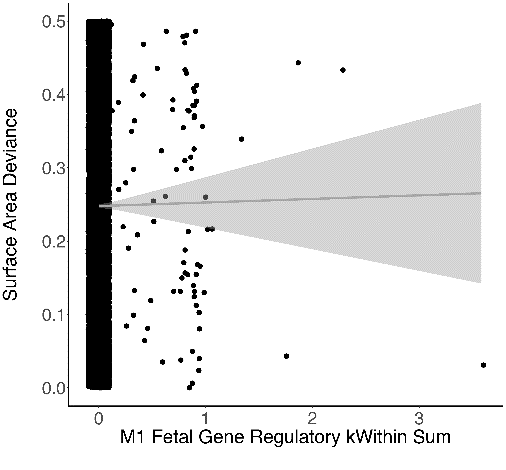

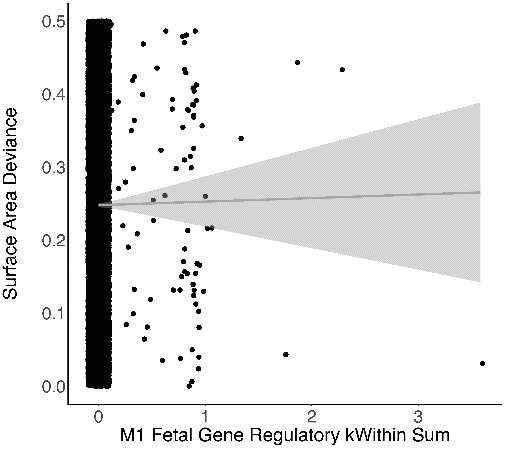

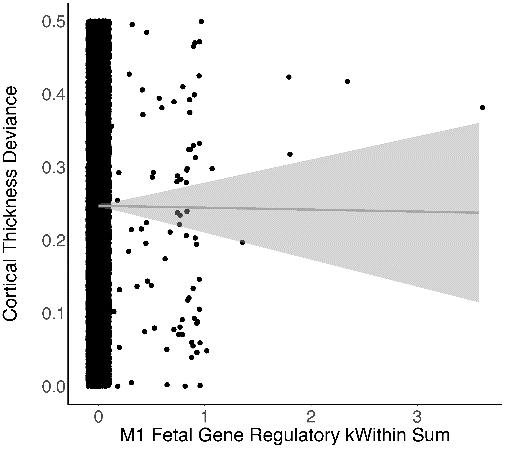

*b* = 0.00, *p* = .81

*b* = -0.00, *p* = .97

*b* = -0.00, *p* = .95

**Supplementary Figure 20.** Associations between M1 deletion gene count based on conservative CNV calls from Sha et al., (2025) and (A) gray matter volume, (B) cortical thickness, or (C) surface area centile or deviance in these centile scores (D-F) in ABCD study subjects without known risk CNVs (*n* = 9,434). Parallel associations between M1 LOEUF score and (G) gray matter volume, (H) cortical thickness, or (I) surface area centile or deviance in these centile scores (J-L). Parallel associations between M1 kWithin score and (M) gray matter volume, (N) cortical thickness, or (O) surface area centile or deviance in these centile scores (P-R). Relationships are plotted using raw centile and M1 scores; reported effect sizes and p-values are from the mixed models. **p* < 0.05.

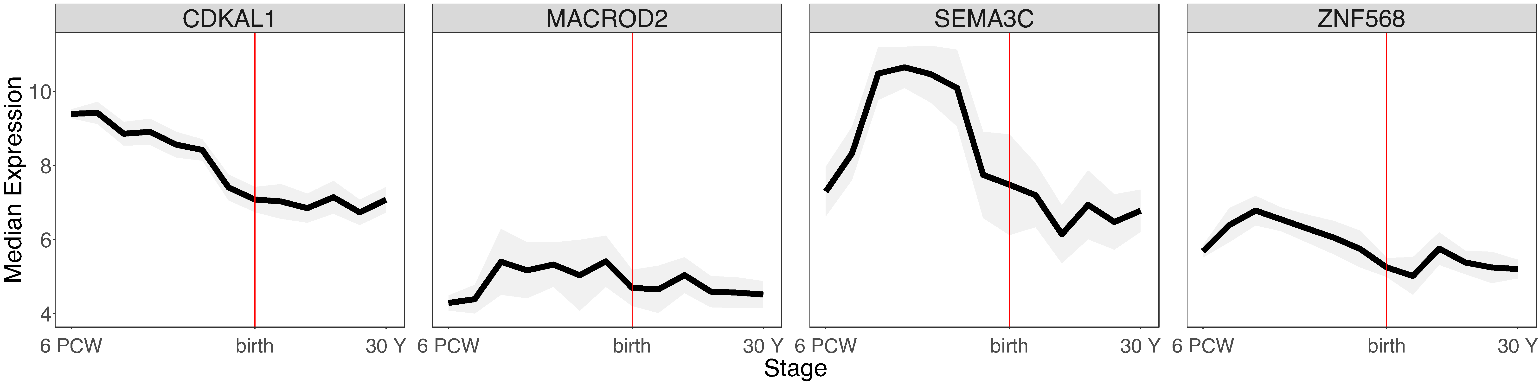

**Supplementary Figure 21.** Expression trajectories of M1 fetal gene regulatory genes of interest from 6 post-conception weeks (PCW) to 30 years of age, found in the SSD-focused sample. M1 genes of interest shown are those outside of NDD-risk loci in SSD cases with borderline IQ: *CDKAL1* (*n* case with deletion = 1, *n* non-case with deletion = 1, 2/2 with borderline IQ; LOEUF percentile score = 0.74; kWithin percentile score = 0.73), *MACROD2* (*n* case with deletion = 1; *n* non-case with deletion = 0, 1/1 with borderline IQ; LOEUF percentile score = 0.79; kWithin percentile score = 0.02), *SEMA3C* (*n* case with deletion = 1, *n* non-case with deletion = 0, 1/1 with borderline IQ; LOEUF percentile score = 0.74; kWithin percentile score = 0.27), *ZNF568* (*n* case with deletion = 1, *n* non-case with deletion = 0, 1/1 with borderline IQ; LOEUF percentile score = 0.47; kWithin percentile score = 0.25).

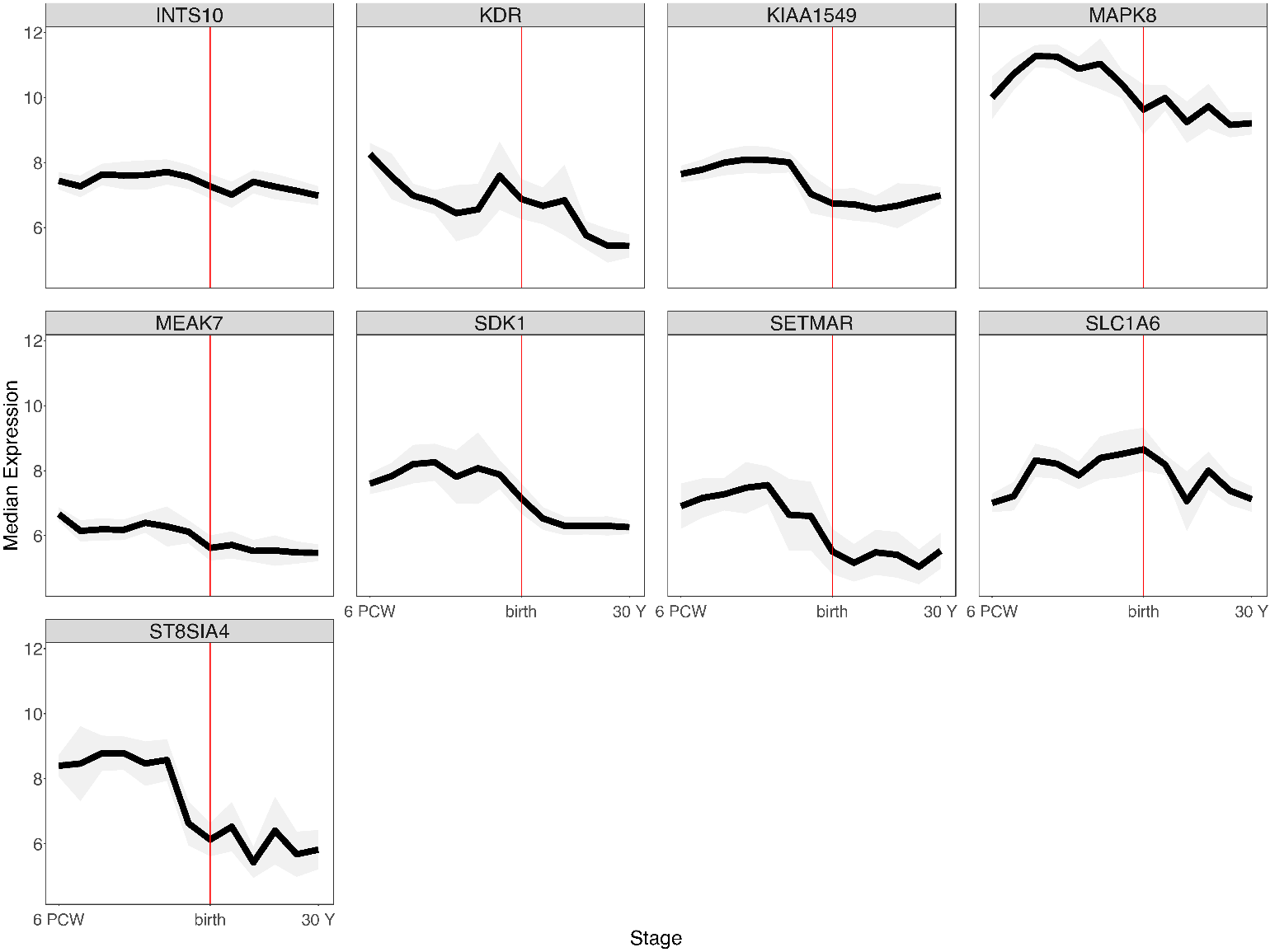

**Supplementary Figure 22.** Expression trajectories from 6 post-conception weeks (PCW) to 30 years of age of M1 fetal gene regulatory genes of interest found in the ABCD Study sample. M1 genes of interest shown are those outside of NDD-risk loci and in which more than 50% of individuals with deletions have borderline IQ: *INTS10* (*n* with deletion = 1, 1/1 with borderline IQ; LOEUF percentile score = 0.97; kWithin percentile score = 0.05), *KDR* (*n* with deletion = 1, 1/1 with borderline IQ; LOEUF percentile score = 0.91; kWithin percentile score = 0.02), *KIAA1549* (*n* with deletion = 2, 2/2 with borderline IQ = 100%; LOEUF percentile score = 0.88; kWithin percentile score = 0.41), *MAPK8* (*n* with deletion = 3, 2/3 with borderline IQ; LOEUF percentile score = 0.94; kWithin percentile score = 0.46), *MEAK7* (*n* with deletion = 5, 3/5 with borderline IQ; LOEUF percentile score = 0.09; kWithin percentile score = 0.10), *SDK1* (*n* with deletion = 1, 1/1 with borderline IQ, LOEUF percentile score = 0.82; kWithin percentile score = 0.27), *SETMAR* (*n* with deletion = 1, 1/1 with borderline IQ; LOEUF percentile score = 0.37; kWithin percentile score = 0.25), *SLC1A6* (*n* with deletion = 1, 1/1 with borderline IQ; LOEUF percentile score = 0.79; kWithin percentile score = 0.01), *ST8SIA4* (*n* with deletion = 1, 1/1 with borderline IQ, LOEUF percentile score = 0.51; kWithin percentile score = 0.44).

Table S1. Summary of recruitment streams and clinical and cognitive measures by contributing research programs.

| Research Program & Location | SSD Patient Recruitment Strategy | SSD *n*  (% Female) | SSD Relative Recruitment Strategy | SSD Relative *n*  (% Female) | Control Recruitment Strategy | Control *n*  (% Female) | Diagnostic Measure | Cognitive Measure* | DNA Sample Tissue Type |
| --- | --- | --- | --- | --- | --- | --- | --- | --- | --- |
| Adolescent Brain-Behavior Research Clinic (ABBRC) - University of California, Los Angeles | Adolescent onset SSD patients (12-17 y/o) | 42 (33.3%) | NA | NA | Demographically similar controls | 15 (40%) | SCID-IV or SCID-5 | WASI 2 Subtest - Verbal + Performance | blood (*n*=46); saliva (*n*=11) |
| Aftercare Program - University of California, Los Angeles | Adult recent-onset SSD patients (18-45 y/o) | 275 (25.4%) | First to third degree relatives of adult SSD patients | 94 (63.8%) | Demographically similar controls | 102 (52.9%) | SCID DSM-III-R, SCID-IV, or SCID-5 | WASI 2 or 4 Subtest - Verbal + Performance (*n* = 242);  WASI Performance Subtest Only (*n* = 11); WAIS-R, WAIS-III, or WAIS-IV Vocabulary Subtest (n = 97); WAIS-R or WAIS-III Vocabulary + Block Design (*n* = 65), MCCB (*n* = 28), WAIS 3-Verbal Subtests (*n* = 7); WTAR (*n* = 6) | blood (*n*=420); saliva (*n*=50); unknown (*n*=1) |
| Child-Onset Schizophrenia Family Study - University of California, Los Angeles | Youth with early-onset SSD | 44 (27.2%) | First to third degree relatives of early-onset SSD patients | 68 (58.8%) | Demographically similar controls | 78 (51.2%) | KSADS or SCID-IV | WASI 2 Subtest - Verbal + Performance (*n* = 171);  WASI Performance Subtest Only (*n* = 11); | blood (*n*=135); saliva (*n*=54); buccal (*n*=1) |
| Multimodal Evaluation of Neural Disorders (MEND) Study - Feinstein Institute for Medical Research | Adolescent recent-onset SSD patients (12-17 y/o) | 69 (26.1%) | NA | NA | Demographically similar controls | 75 (60.0%) | SCID-IV | WASI 2 Subtest - Verbal + Performance | blood (*n*=11); saliva (*n*=133) |
| North American Prodrome Longitudinal Study (NAPLS) 2 & 3 - Multi-Site: Emory University; Harvard Medical School; University of Calgary; University of California Los Angeles; University of California San Diego; University of California San Francisco, University of North Carolina, Chapel Hill, Yale University, Zucker Hillside Hospital | Youth (12-30 y/o) at clinical high risk for psychosis based on Structured Interview for Prodromal States (SIPS) who developed a SSD | 140 (40.7%) | Youth (12-30 y/o) with a first degree SSD relative per Family Interview for Genetic Studies (FIGS) and functional decline who did not develop a SSD within 2 years | 71 (42.3%) | Demographically similar controls | 311 (48.2%) | SCID-IV or SCID-5 | WASI 2 Subtest - Verbal + Performance (*n* = 515); WASI Performance Subtest Only (*n* = 1) | blood (*n*=522) |
| Social Cognition and Functioning in Schizophrenia (SCAF) Study - University of California, Los Angeles | Adult SSD patients (18-60 y/o) | 47 (31.9%) | NA | NA | NA | NA | SCID-IV | MCCB | blood (*n*=47) |
| *WASI = Wechsler Abbreviated Scale of Intelligence, WAIS = Wechsler Adult Intelligence Scale, MATRICS = Measurement and Treatment Research to Improve Cognition in Schizophrenia Consensus Cognitive Battery, WTAR = Wechsler Test of Adult Reading | | | | | | | | | |

Table S2. IQ estimates and age of psychosis onset by primary schizophrenia spectrum diagnosis (SSD) group.

| Schizophrenia Spectrum Disorder (SSD) Group | Mean IQ Estimate (SD) | n with IQ Estimate Information | Mean Age Psychosis Onset (SD) | n with Age Psychosis Onset Information |
| --- | --- | --- | --- | --- |
| Schizophrenia | 97.76 (15.45) | 338 | 20.3 (6.03) | 341 |
| Schizophreniform | 98.94 (16.62) | 86 | 20.76 (3.33) | 93 |
| Schizoaffective disorder | 100.7 (17.06) | 49 | 19.9 (5.41) | 49 |
| Bipolar disorder with psychotic features | 103.7 (15.07) | 20 | 19.78 (5.94) | 18 |
| Brief psychotic disorder | 122 (NA) | 1 | 15 (0) | 1 |
| Delusional disorder | 102.83 (18.71) | 6 | 23 (8) | 6 |
| Major depressive disorder with psychotic features | 99 (19.47) | 7 | 17.71 (3.09) | 7 |
| Psychotic disorder not otherwise specified | 104.31 (15.98)* | 72 | 17.63 (5.09)*^ | 75 |
| Unspecified SSD | 92.58 (16.52) | 19 | 18.53 (4.29) | 17 |

**Pairwise difference from schizophrenia p* < .05*.*

*^Pairwise difference from schizophreniform p* < .05*.*

Table S3. MRI Scanning Protocols and *N*s Included Per Protocol in Analyses Following Quality Control.

| Study/Protocol | Scanner Vendor and Type | Field Strength | Slice Thickness (mm) | TR (ms) | TE (ms) | TI (ms) | Flip Angle | N Controls Used for Scanner/Protocol Normalization | N SSD Subjects in CNV Analysis | N SSD Relatives in CNV Analysis | N Controls in CNV Analysis |
| --- | --- | --- | --- | --- | --- | --- | --- | --- | --- | --- | --- |
| Aftercare_1 | Siemens Avanto | 1.5T | 1 | 2000 | 2.49 | 900 | 8 | 22 | 56 | 0 | 0 |
| Aftercare_2 | Siemens TrioTim | 3T | 1.20 | 2300 | 2.91 | 900 | 9 | 38 | 75 | 0 | 0 |
| Aftercare_3 | Siemens Prisma Fit | 3T | 1 | 2500 | 1.81 | 1000 | 8 | 15 | 24 | 0 | 0 |
| Aftercare_4 | Siemens Sonata | 1.5T | 1 | 1900 | 4.38 | 1100 | 15 | 89 | 43 | 65 | 81 |
| NAPLS2_Site01_1 | Siemens TrioTim | 3T | 1.20 | 2300 | 2.91 | 900 | 9 | 24 | 9 | 2 | 24 |
| NAPLS2_Site02_1 | Siemens TrioTim | 3T | 1.20 | 2300 | 2.91 | 900 | 9 | 25 | 4 | 3 | 25 |
| NAPLS2_Site03_1 | Siemens TrioTim | 3T | 1.20 | 2300 | 2.91 | 900 | 9 | 27 | 3 | 3 | 27 |
| NAPLS2_Site04_1 | GE TwinSpeed | 3T | 1.20 | 7.00 | minimum full | 400 | 8 | 25 | 3 | 1 | 25 |
| NAPLS2_Site05_1 | Siemens TrioTim | 3T | 1.20 | 2300 | 2.91 | 900 | 9 | 27 | 11 | 2 | 27 |
| NAPLS2_Site07_1 | GE 3T/94 | 3T | 1.20 | 7.00 | minimum full | 400 | 8 | 24 | 18 | 15 | 24 |
| NAPLS2_Site08_1 | Siemens TrioTim | 3T | 1.20 | 2300 | 2.91 | 900 | 9 | 33 | 7 | 3 | 31 |
| NAPLS3_Site02_1 | Siemens TrioTim | 3T | 1 | 2400 | 1.90 | 1000 | 8 | 11 | 7 | 2 | 11 |
| NAPLS3_Site06_1 | GE Discovery MR750 | 3T | 1 | 4.00 | 1.33 | 650 | 11 | 11 | 6 | 2 | 11 |
| NAPLS3_Site07_1 | GE Discovery MR750 | 3T | 1 | 4.64 | 2.00 | 650 | 11 | 11 | 4 | 1 | 11 |
| NAPLS3_Site09_1 | Siemens Skyra | 3T | 1 | 2400 | 1.96 | 1000 | 8 | 11 | 11 | 1 | 11 |

Table S4. Baseline demographic, clinical, and CNV burden characteristics for included ABCD subjects.

| ABCD Cohort After Quality Control | |
| --- | --- |
| *N* Subjects (% Female) | 9930 (47.17%) |
| Self-Identified Race |  |
| White *n* (%) | 6494 (65.4%) |
| Black/African American *n* (%) | 1653 (16.6%) |
| Asian *n* (%) | 180 (1.8%) |
| American Indian / Alaska Native / Native Hawaiian *n* (%) | 52 (0.5%) |
| >1 *n* (%) | 1024 (10.3%) |
| Other/Unknown *n* (%) | 527 (5.3%) |
| Hispanic/Latino *n* (%) | 1882 (19.0%) |
| *N* with Included Deletions (%) | 2834 (28.54%) |
| *N* with Included Duplications (%) | 3527 (35.52%) |
| Mean Size of Included Deletions (SD) | 261,492 (426,228) |
| Mean Size of Included Duplications (SD) | 546,983 (626,293) |
| *N* with Included Genic Deletions (%) | 1426 (14.36%) |
| *N* with Included Genic Duplications (%) | 2962 (29.83%) |
| Mean Genes per Genic Deletion (SD) | 2.58 (3.91) |
| Mean Genes per Genic Duplication (SD) | 5.04 (5.43) |
| *N* with SSD / NDD Risk CNV Hits (% Subjects) | 80 (0.81%) / 154 (1.55%) |
| Mean NIH-TB Total Composite IQ ± SD (% Borderline IQ) | 100.09 ± 17.87 (19.74%) |
| Mean Age of NIH-TB Testing (SD) | 9.92 (0.62) |

Table S5. Genotyping array signal intensity quality metrics for included subjects by study cohort.

| Study | Array | Mean Log R Ratio SD (SD) | Mean B Allele Frequency SD (SD) | Mean Absolute Waviness Factor (SD) |
| --- | --- | --- | --- | --- |
| SSD-Focused Cohort | Illumina Global Screening Array | 0.094 (0.026) | 0.030 (0.004) | 0.006 (0.002) |
| ABCD Study Cohort | Affymetrix Smokescreen Array | 0.255 (0.038) | 0.072 (0.009) | 0.019 (0.004) |

Table S6. CNV burden characteristics for included ABCD subjects using Sha et al., CNV calls.

| ABCD Cohort After Quality Control | |
| --- | --- |
| *N* Subjects (% Female) | 9930 (47.17%) |
| *N* with Included Deletions (%) | 1062 (10.69%) |
| *N* with Included Duplications (%) | 2217 (22.33%) |
| Mean Size of Included Deletions (SD) | 354,432 (519,257) |
| Mean Size of Included Duplications (SD) | 559,770 (621,965) |
| *N* with Included Genic Deletions (%) | 533 (5.37%) |
| *N* with Included Genic Duplications (%) | 1868 (18.81%) |
| Mean Genes per Genic Deletion (SD) | 2.68 (4.14) |
| Mean Genes per Genic Duplication (SD) | 4.46 (5.67) |
| *N* with SSD / NDD Risk CNV Hits (% Subjects) | 45 (0.45%) / 97 (0.98%) |
| *N* with M1 Gene Deletion | 102 (1.02%) |
| Mean NIH-TB Total Composite IQ ± SD (% Borderline IQ) | 100.09 ± 17.87 (19.74%) |
| Mean Age of NIH-TB Testing (SD) | 9.92 (0.62) |

Table S7. Model statistics for MRI metrics in the ABCD study cohort, including covariates for tissue type, batch, or non-M1 deletion score.

|  |  | Tissue Covariate | | Batch Covariate | | Non-M1 Score Covariate | |
| --- | --- | --- | --- | --- | --- | --- | --- |
| M1 Model | MRI Metric | Beta (95% CI) | P Value | Beta (95% CI) | P Value | Beta (95% CI) | P Value |
| M1 Gene Count | GMV Centile | -0.02 [-0.05,0.01] | 0.221 | -0.02 [-0.05,0.01] | 0.229 | -0.01 [-0.04,0.02] | 0.642 |
| M1 Gene Count | CT Centile | 0.02 [-0.01,0.05] | 0.204 | 0.02 [-0.01,0.05] | 0.212 | 0.02 [-0.01,0.05] | 0.194 |
| M1 Gene Count | SA Centile | -0.02 [-0.05,0.01] | 0.129 | -0.02 [-0.05,0.01] | 0.139 | -0.01 [-0.04,0.02] | 0.45 |
| M1 Gene Count | GMV Centile Deviance | 0.01 [0,0.03] | 0.07 | 0.01 [0,0.03] | 0.078 | 0.02 [0,0.03] | **0.033** |
| M1 Gene Count | CT Centile Deviance | 0.01 [0,0.03] | 0.064 | 0.01 [0,0.03] | 0.068 | 0.02 [0,0.03] | 0.051 |
| M1 Gene Count | SA Centile Deviance | 0.02 [0,0.03] | **0.041** | 0.02 [0,0.03] | **0.043** | 0.02 [0,0.04] | **0.017** |
| M1 LOEUF Sum | GMV Centile | -0.06 [-0.11,0] | **0.038** | -0.06 [-0.11,0] | **0.039** | -0.04 [-0.09,0.02] | 0.237 |
| M1 LOEUF Sum | CT Centile | 0.03 [-0.02,0.09] | 0.256 | 0.03 [-0.02,0.09] | 0.265 | 0.02 [-0.04,0.09] | 0.466 |
| M1 LOEUF Sum | SA Centile | -0.06 [-0.11,-0.01] | **0.028** | -0.06 [-0.11,-0.01] | **0.031** | -0.03 [-0.09,0.03] | 0.282 |
| M1 LOEUF Sum | GMV Centile Deviance | 0.02 [-0.01,0.05] | 0.163 | 0.02 [-0.01,0.05] | 0.173 | 0.03 [-0.01,0.06] | 0.116 |
| M1 LOEUF Sum | CT Centile Deviance | 0.04 [0.01,0.07] | **0.005** | 0.04 [0.01,0.07] | **0.006** | 0.04 [0.01,0.07] | **0.009** |
| M1 LOEUF Sum | SA Centile Deviance | 0.02 [-0.01,0.05] | 0.13 | 0.02 [-0.01,0.05] | 0.131 | 0.03 [-0.01,0.06] | 0.114 |
| M1 kWithin Sum | GMV Centile | -0.03 [-0.07,0.02] | 0.25 | -0.03 [-0.07,0.02] | 0.257 | -0.01 [-0.06,0.04] | 0.656 |
| M1 kWithin Sum | CT Centile | 0.03 [-0.02,0.08] | 0.176 | 0.03 [-0.02,0.08] | 0.182 | 0.03 [-0.02,0.08] | 0.256 |
| M1 kWithin Sum | SA Centile | -0.04 [-0.08,0.01] | 0.12 | -0.04 [-0.08,0.01] | 0.128 | -0.02 [-0.07,0.03] | 0.468 |
| M1 kWithin Sum | GMV Centile Deviance | 0.02 [0,0.05] | 0.067 | 0.02 [0,0.05] | 0.073 | 0.02 [0,0.05] | 0.068 |
| M1 kWithin Sum | CT Centile Deviance | 0.03 [0,0.05] | **0.04** | 0.03 [0,0.05] | **0.042** | 0.02 [0,0.05] | 0.104 |
| M1 kWithin Sum | SA Centile Deviance | 0.02 [0,0.05] | 0.08 | 0.02 [0,0.05] | 0.081 | 0.02 [0,0.05] | 0.089 |

**SUPPLEMENTARY REFERENCES**

1.	First, Michael B., Spitzer, Robert L, Gibbon Miriam, and Williams, Janet B.W: Structured Clinical Interview for DSM-IV-TR Axis I Disorders, Research Version, Patient Edition. (SCID-I/P). New York, Biometrics Research, New York State Psychiatric Institute, 2002

16.	Wechsler D: WAIS-R Manual: Wechsler Adult Intelligence Scale-revised. Psychological Corporation, 1981

17.	Wechsler D: Wechsler Abbreviated Scale of Intelligence. San Antonio, TX, The Psychological Corporation, 1999

18.	Nuechterlein KH, Green MF, Kern RS, et al.: The MATRICS Consensus Cognitive Battery, part 1: test selection, reliability, and validity. Am J Psychiatry 2008; 165:203–213

19.	Wechsler D: Wechsler Test of Adult Reading: WTAR. Psychological Corporation, 2001
